# COVID-19 spread and Weather in U.S. states: a cross-correlative study on summer-autumn 2020

**DOI:** 10.1101/2021.01.29.21250793

**Authors:** Emmanuel de Margerie

## Abstract

An effect of weather on sars-cov-2 transmission is regularly proposed as a putative cause of unexplained fluctuations of covid-19 new cases, but clear data supporting this hypothesis remains to be presented. Here I measured longitudinal time-series correlations between outdoor temperature, humidity and covid-19 reproduction number (Rt) in the 50 U.S. states (+DC). In order to mitigate the confounding influence of varying social restriction measures, the analysis spans a 5-month period during summer and autumn 2020 when restrictions were comparatively lower and more stable. I used a cross-covariance approach to account for a variable delay between infection and case report. For a delay near 11 days, most U.S. states exhibited a negative correlation between outdoor temperature and Rt, as well as between absolute humidity and Rt (mean r = −0.35). In 21 states, the correlation was strong (r < −0.5). Individual state data are presented, and associations between cold and/or dry weather episodes and short-term new case surges are proposed. After identifying potential confounding factors, I discuss 3 possible causal mechanisms that could explain a correlation between outdoor weather and indoor disease transmission: behavioral adaptations to cold weather, respiratory tract temperature, and the importing of outdoor absolute humidity to indoor spaces.

## Introduction

A full year after the start of the covid-19 epidemic, the fluctuations of contamination numbers through time remain only partially understood and often unpredictable, putting populations and health systems at risk, and challenging social restrictions policies [1].

An influence of weather on covid-19 transmission was repeatedly assessed early in the pandemic (review in [2]), but wider acceptance of such a “weather effect” could still benefit today from stronger evidence. Here l investigated correlations between outdoor temperature, humidity and covid-19 spread, using longitudinal data for the 50 U.S. states (+Columbia district), during summer and autumn 2020.

The present work is mainly exploratory and descriptive, with the objective to stimulate further investigations by expert scientists (e.g. epidemiologists, virologists, atmospheric science and air quality specialists).

## Methods

### Covid-19 data

I used daily new covid-19 cases counts (new cases, NC) collected for the 50 states (+ DC) by the *Covid Tracking Project* [3] (i.e. *positiveIncrease* variable in original dataset).

Raw new case counts were 7-day averaged in order to smooth day-of-week fluctuations (e.g. lower case counts on weekends). Delay-free symmetric averaging (from day-3 to day+3) was applied:

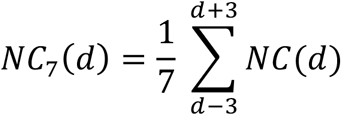

### Delay from infection to case report

New cases reported on a given date have been infected several days before. This delay can be estimated considering the following elements:

∘ the delay from exposure to symptom onset is typically 5 days, but can vary from 2 to 11 days [4].
∘ the optimal time for RT-PCR testing (lowest false-negative) is 3 days after symptom onset [5]
∘ in U.S., average reported delay between testing and result was 3-4 days (June-September 2020 data [6]).
∘ on the other hand, antigen tests should be conducted within 7 days of symptom onset, and can provide same-day results [7].
∘ the new case data in the *Covid Tracking Project* dataset is not reported by date of testing, but rather by date of publication by the state administration, and states have variable reporting delays (see faq in [3]).

Hence the delay from infection to case report (**ICR**) might be as short as ∼5 days for a test with same-day result performed early after infection (e.g. systematic testing of asymptomatic people such as university students). On the opposite, the delay can potentially attain 3 weeks for symptomatic cases tested one week or so after symptom onset, in a state with testing and/or reporting delays. In between these extreme values, I estimated the ICR delay in the *Covid Tracking Project* dataset was most probably within 12 ± 5 days, depending on each state’s testing and reporting swiftness. Conversely, new cases reported on a given date were assumed to reflect contaminations about 12-day old.

### Disease spread

If one is interested in quantifying disease spread (under the putative immediate influence of an environmental factor), a relevant estimator of transmission is not new case count *per se* (NC, which depends on the state’s accumulated transmission history), but rather the effective reproduction number **Rt**. Rt is the average number of new infections caused by a single infected individual. Several precise Rt computation methods exist, based on refined models (see [8] and [9]). Here I simply estimated Rt as the ratio of new infections to the concurrent infectious cases that got infected on the previous generation. Covid-19 generation time being close to one week [9], I used:

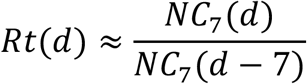

Note that, as Rt is based on reported cases and not current infection numbers, its value, as was the case for NC, reflects transmission with a ∼12 days delay. Rather than time-shifting Rt values to (day-12) upon calculation (as is done elsewhere, e.g. [10]), I voluntarily left Rt estimates in the same time frame as the new cases data, for easier graphical presentation of results.

### Weather data

I used daily average temperature (**T**, outdoor) and relative humidity (**RH**, outdoor), as reported for each US county in the *covid-19-open-data* dataset [11] (variable names in original dataset : *average_temperature* and *relative_humidity*).

According to the dataset information, this weather data was compiled from multiple NOAA stations for each county : *“The reported data corresponds to the average of the nearest 10 stations within a 300km radius”* [11]. Additionally, the dataset contains single daily T and RH values for each state, which are the values I used in the present work. To my knowledge, it is unfortunately not indicated how these aggregated values by state were derived from county values.

As RH levels are not directly comparable when temperature varies, two other daily variables were computed from RH and T:

- Absolute humidity (**AH**, outdoor) is the quantity of water contained per air volume, in g/m^3^. As cold air cannot contain as much water vapor as hot air, AH depends on T and RH and was computed as follows:

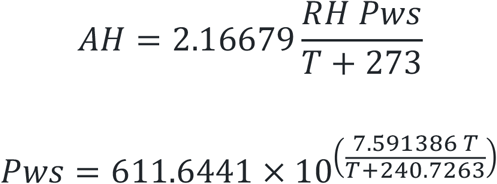

with: AH: absolute humidity (g/m^3^) RH: relative humidity (0 to 1) T: temperature (°C) Pws: water vapor saturation pressure (Pa) Note that here, daily mean AH is directly computed from daily mean T and RH, which is not 100% correct, as hourly T and RH (not available) should first be converted to hourly AH before averaging. I included outdoor AH in the analysis because it was already identified as a factor (along T) of more frequent respiratory diseases [12], and it can be a proxy to indoor RH (see discussion).
- Wet-bulb temperature (**WBT**, outdoor) is the temperature experienced by wet objects: if the surrounding air is not saturated with water vapor (RH < 100%), evaporation of water from a wet object cools down the object below ambient air temperature. Hence WBT is lower than T, and the drier the air is (low RH), the lower is WBT. Here, WBT is used as a candidate proxy to the temperature of the upper respiratory tract for people breathing outdoor, as cold and dry air cools down mucosa [13] and potentially affects biological processes therein [14,15]. Daily WBT was estimated from daily T and RH using the empirical formula in [16].

T, WBT and AH were then 7-day averaged using the same method as for covid-19 cases.

### Study period

Strong social restrictions, such as applied during lockdowns (e.g. business closure, stay-at-home orders) are known to have spectacular effects on disease transmission. These human-policy effects are hypothesized to add-up with (or possibly overcome) any putative weather effect. Hence, for the present work, I excluded periods when the strength of restriction policies are known to have varied significantly. Most US states lifted “first-wave” lockdowns between late April and late May 2020, with a few states lifting later in early June [10]. More recently, “3^rd^ wave” surges forced states to progressively put back strong restrictions, during November or December 2020 depending on states (as reported in the press e.g. [17] and [18]). The interval period chosen here, from June 15th 2020 to November 15th 2020, is supposed to have seen relatively lower and more stable restrictions levels in most U.S. states.

### Statistical approach

For each state, I computed the Pearson product-moment correlation coefficient (noted **r**) between each weather variable (T, WBT, AH) and covid-19 Rt, across the whole 5-month study period (i.e. n = 154 days). Before computing r, state Rt data was shifted by a delay from 1 to 30 days, in order to observe the effect of assumed ICR delay. This approach is similar to normalized cross-covariance (e.g. *xcov()* Matlab function), but here the delayed variable (Rt) is never clipped at the end of the time series, i.e. n = 154 data value pairs is always verified. Note that this implies that for long delays, covid-19 case counts from late November and early December were included.

The resulting **5-month r** values indicate whether disease transmission followed weather variation over the study period as a whole. r can vary form −1 to 1, and the closer r is to −1 or 1, the stronger the correlation (negative or positive correlation, respectively). Correlation strength was evaluated based on Cohen’s guidelines on effect sizes [19]:

∘ strong correlation: 0.5 ≤ | r |
∘ moderate correlation: 0.2 ≤ | r | < 0.5
∘ weak correlation: | r | < 0.2

Note that with n = 154, any | r | ≥ 0.15 is in theory significant to the p < 0.05 level. However, data are smoothed time series containing some auto-correlation that should be removed before assessing correlation significance. For the present exploratory work, I did not go further into significance testing at this point. Effect size as estimated through r value convey more information than significance thresholding for describing the results below.

In order to evaluate short-term correlation between weather variable and Rt, I also computed r in 31-days sliding windows, with again a variable delay applied on Rt series. These **1-month r** values help precise at which time-scale, and on which sub-period, the correlation between weather and Rt is observed.

I used Matlab® for all analyses and graphics.

## Results

Figure 1 presents synthetic correlation results. Thin lines represent individual states. Thick lines show mean of 51 states. Note that as most detected correlations are negative (i.e. r < 0), vertical axes have been flipped upside-down for easier visualization of later results. **5-month correlations** show that Rt was most often negatively correlated with weather variables (T, WBT, AH) over the full study period. Strongest mean correlation were measured for an ICR of 11, 10 and 11 days respectively (r = −0.35, −0.36, −0.35). | r | near 0.3 suggest moderate correlations strength by common standards [19], but some states had stronger or weaker correlations (strongest correlation −0.77, −0.77 and −0.75 for T, WBT and AH respectively).

**Figure 1.**
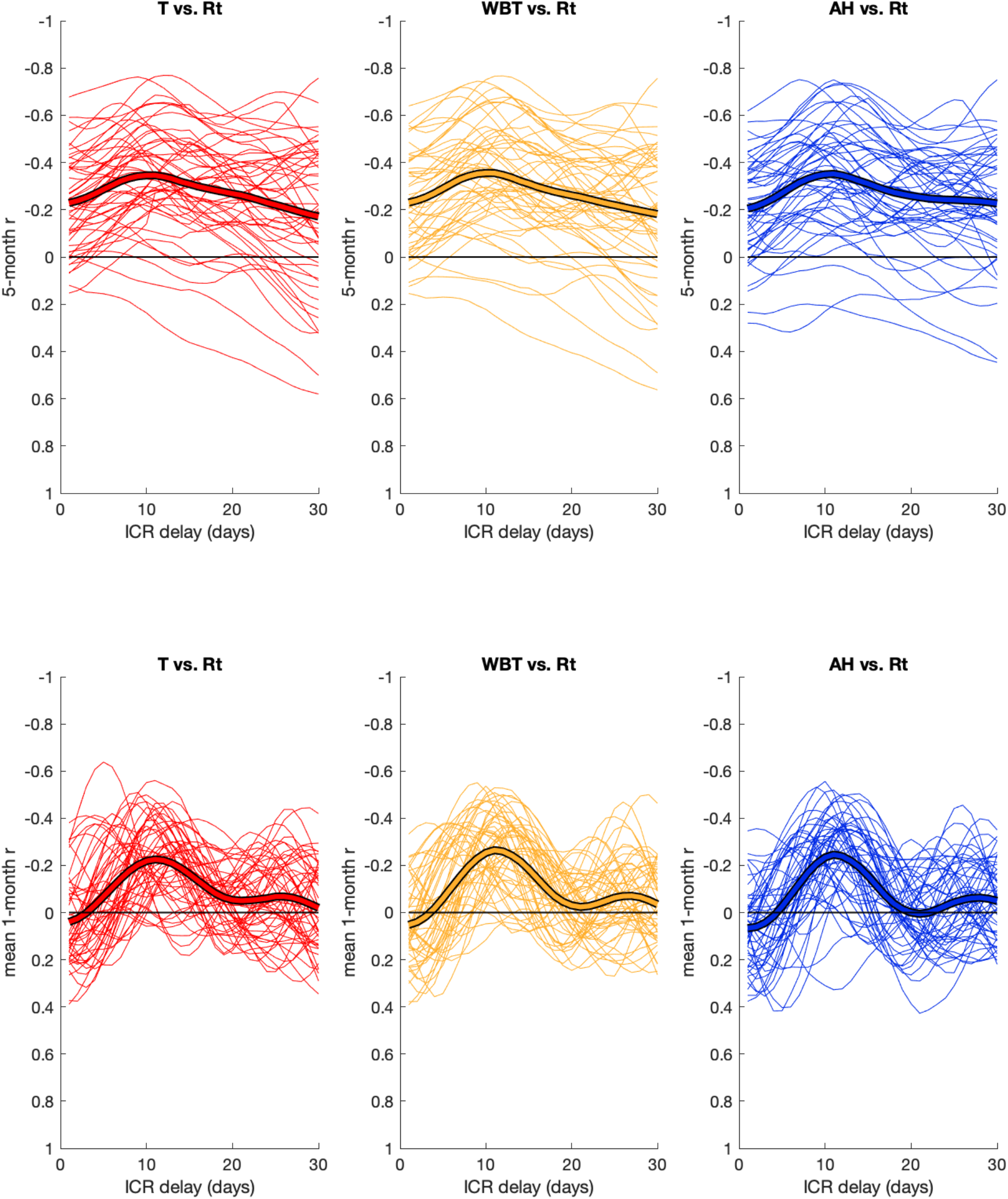
Correlation results for all U.S. states.

**1-month sliding window correlation** provides a time-series of r values for each state, that was averaged in the present figure (mean for 124 windows of 31 days). Thus each thin line is the mean r for a given state, as a function of ICR, and thick line is the mean of means. These short-term correlations show a similar peak in correlation strength, for an ICR delay of 11 days (mean r: −0.23, −0.27 and −0.25 for T, WBT and AH, respectively). For very short or very long delays, 1-month correlations tend to disappear (r ≈ 0), which was not the case for 5-month r.

### 21 states with strongly negative weather-Rt correlations (r ≤ −0.5)

In the following state-specific figures, the state’s time-series for new cases, Rt, T, WBT and AH are plotted on the left. Results of correlations (5-month and mean 1-month) are plotted on the right, as a function of ICR delay. The ICR value within 12±5 days that yielded the most negative r value (5-month or 1-month, T or WBT or AH) was chosen to shift displayed new cases and Rt time-series horizontally. On the bottom left, the state’s 1-month r time-series for the displayed ICR is shown. Note that, as previously, vertical axes are flipped upside-down for weather variables and r plots, for easier comparison between Rt and weather data. States are presented in decreasing correlation strength order. States with weak or no correlation will be described in another section.

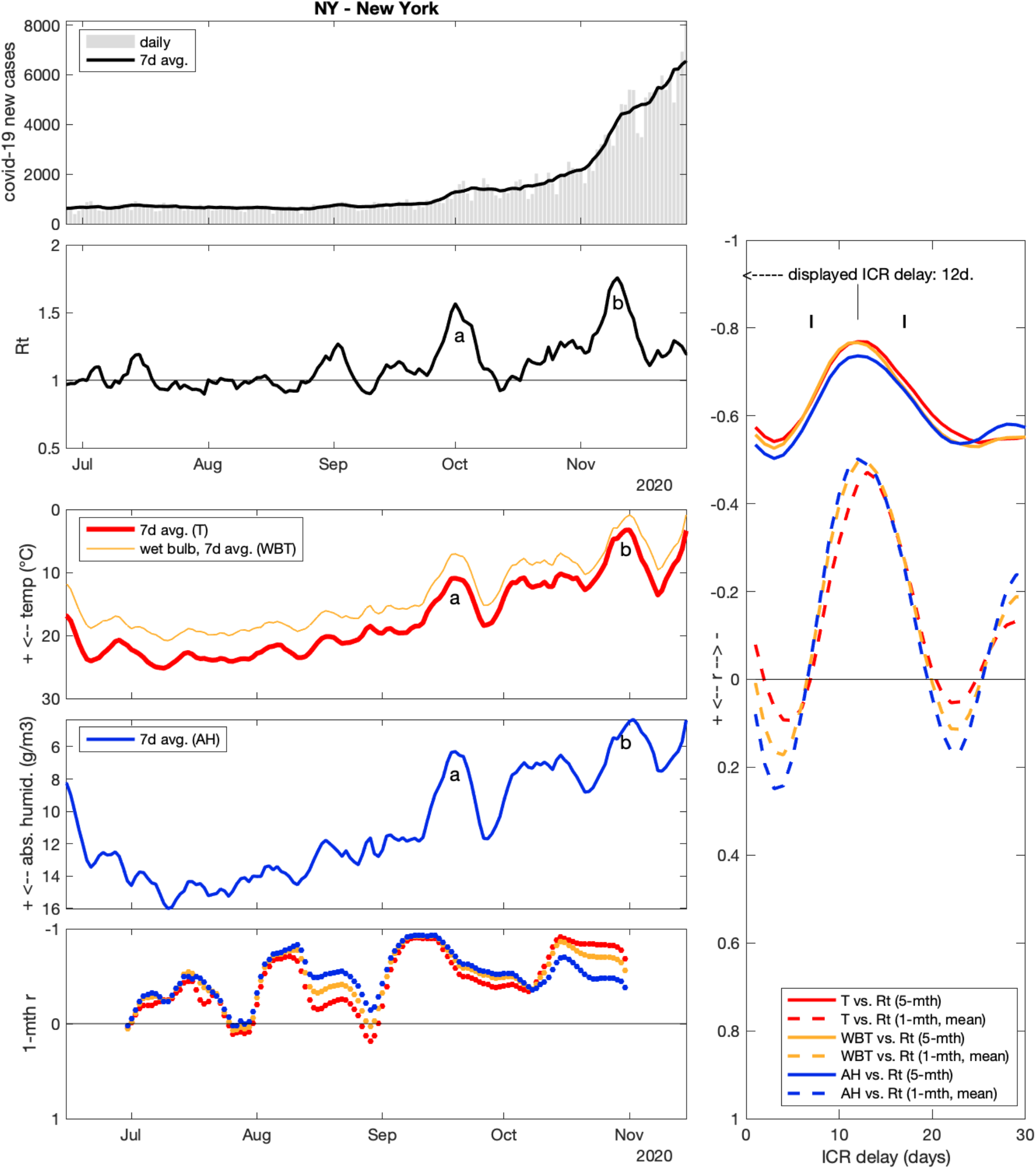

### New York state (NY)

I first describe correlation values (right panel). In NY state, the strongest 5-month correlations with Rt were found for an ICR of 12 days, with r values of −0.77 for T and WBT and −0.74 for AH. 1-month correlations showed a marked peak for similar delays (r = −0.47, −0.50 and −0.50 at 13, 13 and 12 days, for T, WBT and AH, respectively). Looking at time series (left panels) through the study period as a whole, Rt showed an overall increasing trend, while T, WBT and AH decreased over the period (n. b. series are flipped vertically). These general trends explain the negative 5-month r, regardless of the assumed ICR delay. More interesting is the smaller-scale parallelism between Rt and weather data. Such “phase correlation” causes the marked peak in mean 1-month r around 12 days delay, as well as the additional “boost” in 5-month r value.

In concrete terms, NY state experienced two main covid-19 surges during the study period. A first significant surge (a) happened around Oct 1, with new daily cases increasing from ∼800 on Sep 20 to ∼1400 on Oct 10. This translates into a peak in the Rt curve, culminating at Rt = 1.6 on Oct 1. This surge can be related to a cold weather episode around Sep 19, when outdoor temperature temporarily fell from 18°C to 11°C (AH simultaneously fell from 12 to 6 g/m^3^). The second surge (b) happened around Nov 10 (Rt = 1.8, daily new cases increasing from ∼2500 to ∼5000). There was again a corresponding cold episode, around Oct 31 (i.e. 10 days before), with temperature falling from 13°C to 3°C (and AH from 9 to 4 g/m^3^). The parallelism is not perfect, and smaller anomalies do not always coincide between Rt and weather data (Rt inevitably contains some amount of noise, see discussion). The lowest left panel shows the evolution of 1-month r (for ICR = 12 d.), for Rt vs. T, WBT and AH.

In NY the variation of WBT and AH was very similar to T, yielding very similar r values (both long- and short term), regardless of the chosen weather variable. Thus from NY state data, it is unclear whether temperature or humidity should be considered a more likely driver of infections.

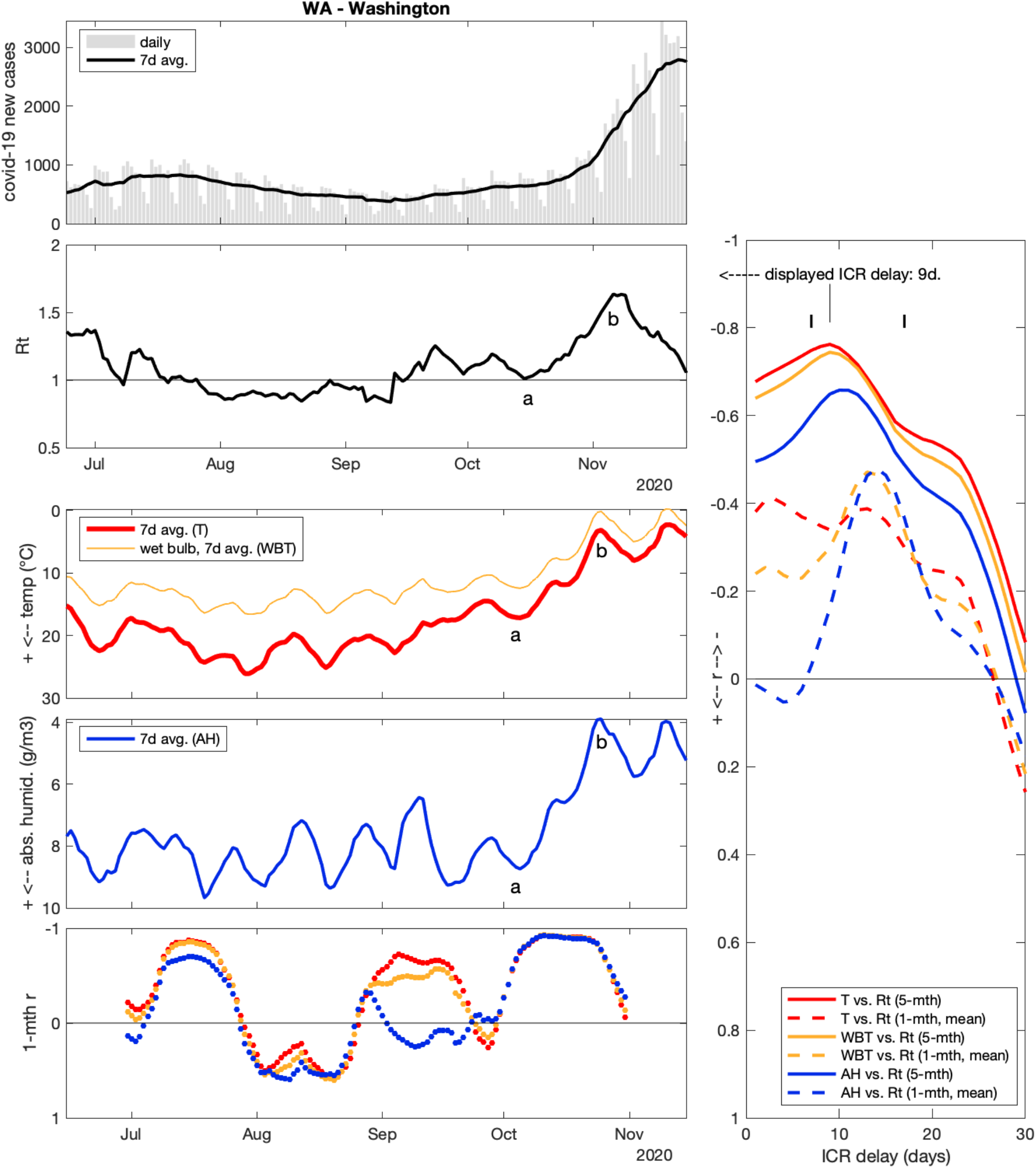

### Washington state (WA)

Strongest 5-month correlation (r = −0.76) was found for T vs Rt, assuming a 9-day ICR (on the lower side of the assumed 12±5 days realistic range for ICR). T had an overall U-shaped evolution (on inverted axes) that was also found in Rt (except for Nov). Results are less clear for WA than for NY, as short-term correlation for WBT and AH were stronger for a 13-14 days ICR. Linking weather and Rt episodes is also less easy, but I still note that a steep decrease in T and AH from Oct 5 to Oct 25 (from 17 to 3°C and 9 to 4 g/m^3^, (a) to (b) on figure) was followed by an Rt increase from 1.0 on Oct 16 to 1.6 on Nov 6, i.e. with a 11-12 days delay. This corresponds to the main case surge observed in WA during the study period, with daily cases increasing form ∼650 mid-Oct to ∼2500 mid-Nov.

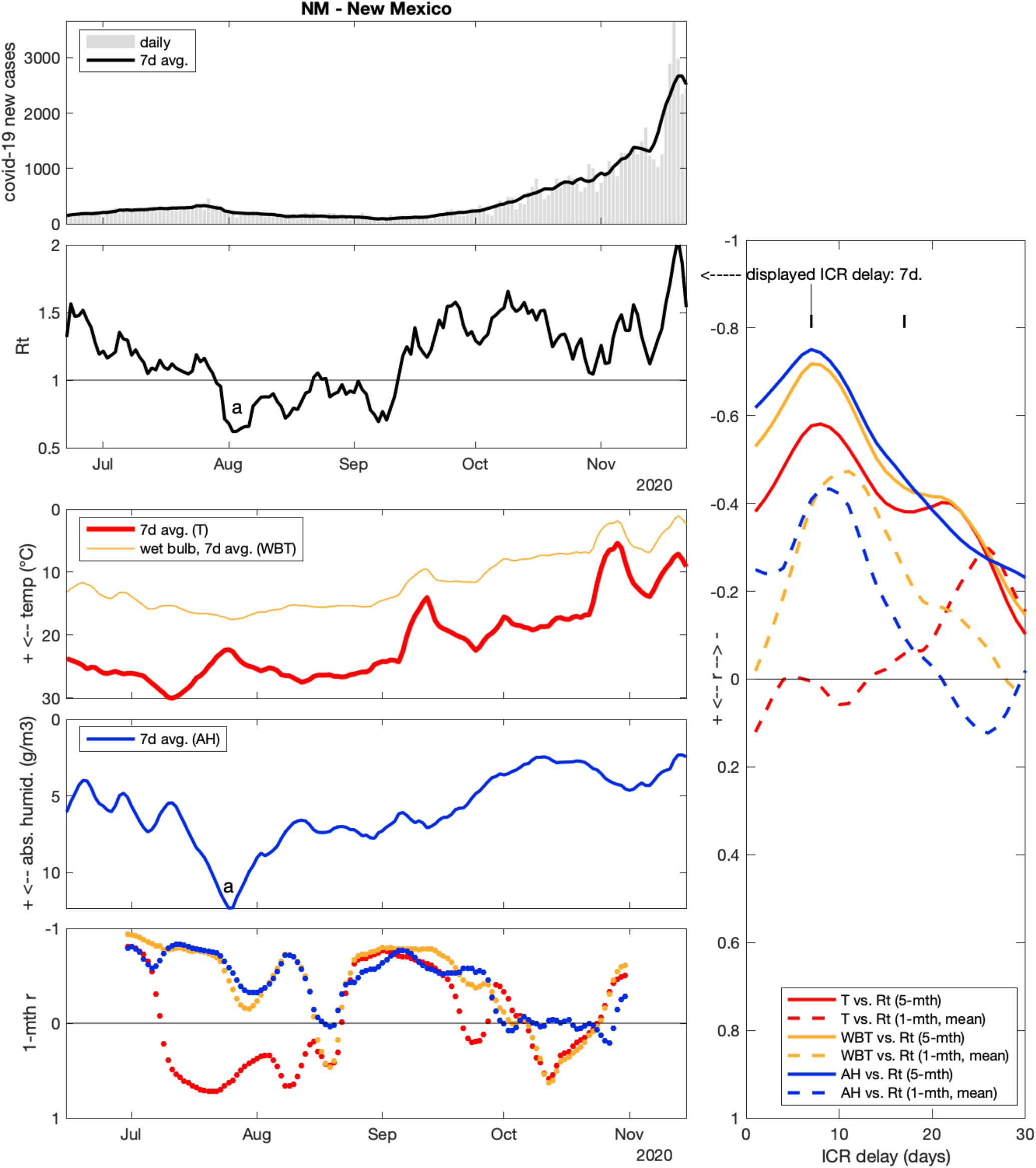

### New Mexico (NM)

Rt was best correlated with AH, for an ICR of 7 days (at the lower limit of realistic range for ICR). As a visual translation, both AH and Rt exhibited an overall V- or W-shaped evolution over the 5-month period, with a marked AH peak (a) on Jul 25 (12 g/m^3^) that corresponds to a minimal Rt of 0.6 on Aug 2 (i.e. 8 days later). Interestingly, during summer, AH or WBT appear as more consistent predictors of Rt than T (see lower left panel). Contrary to NY or WA, finding graphic correspondence between sharp weather episode and Rt peaks (as a visual support for correlation results) was not so easy during the autumn.

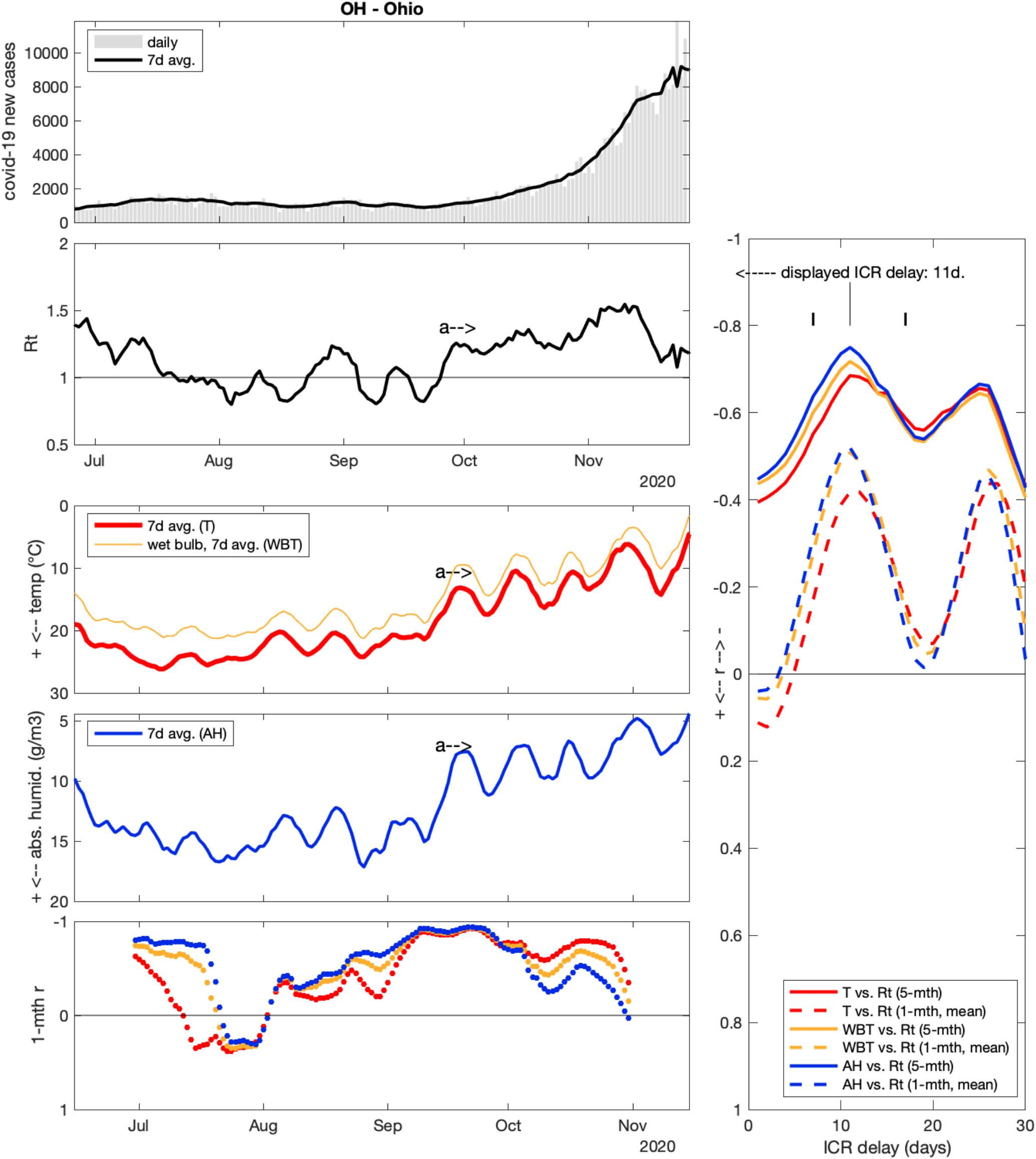

### Ohio (OH)

Long- and short-term correlations congruently suggested ICR = 11 days, for all 3 weather variables (strongest r: −0.75 for AH). r values have another peak when correlating with ∼26-day offset, which goes beyond realistic ICR delays. This “rebound” is presumably caused by periodic phase correlation (both Rt and weather tended to oscillate in OH). The main covid-19 event in OH was a prolonged exponential increase in new cases starting around Sep 26, with Rt remaining within 1.2-1.5 for the following 6 weeks. In the meantime, daily new cases increased exponentially from ∼1000 to ∼7500. This durable increased covid-19 transmission level from late Sep (a) was preceded by an equivalent decrease in T (dropping and remaining below 18°C) and AH (below 12 g/m^3^) on Sep15, i.e. 11 days earlier.

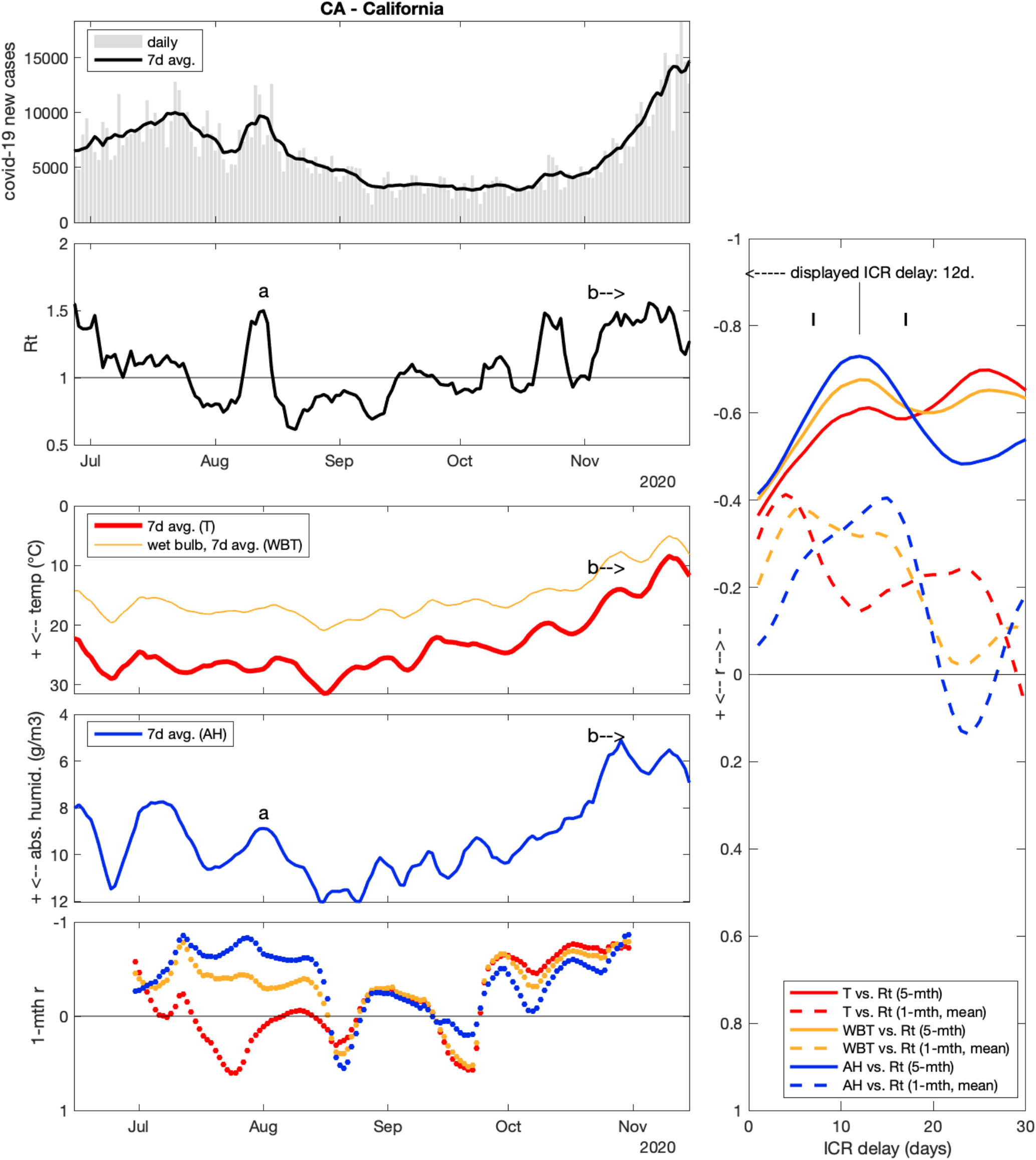

### California (CA)

Rt was most strongly correlated with AH (r = −0.73, for ICR = 12 days). 5-month results for T and WBT, as well as 1-month results, were less clear. As noteworthy episodes in CA that can illustrate correlation results, there was a short surge (a) around Aug 13 (Rt = 1.5) that could correspond to a local dry episode around Aug 1, and (b) the sustained surge starting around Nov 5 (Rt remaining above 1.3 for 3 weeks) coincided with AH dropping below 7 g/m3 (and T below 17°C) near Oct 24, i. e. 12 days earlier.

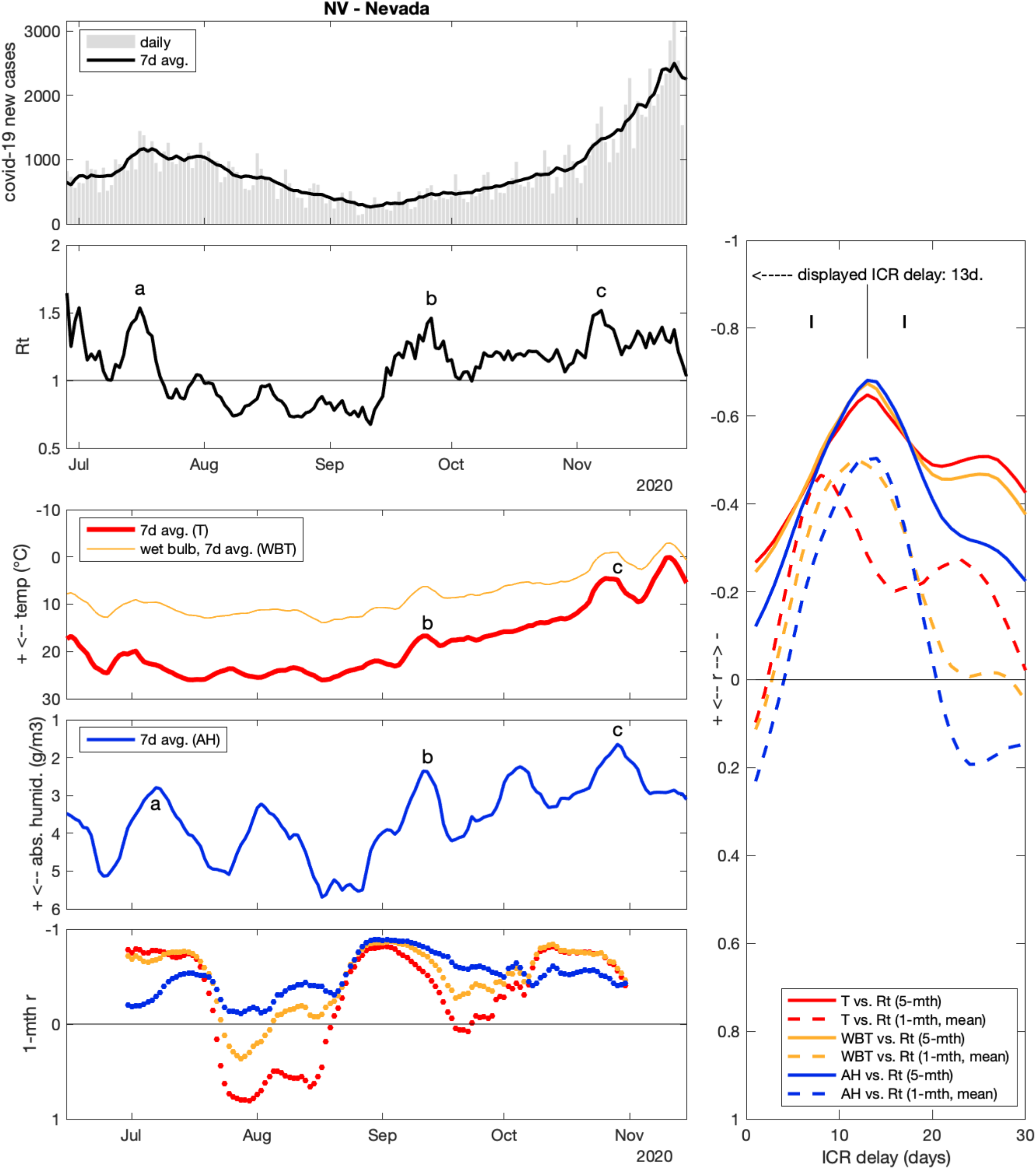

### Nevada (NV)

5-month r indicated strongest correlations for ICR = 13 days, with similar values for T, WBT and AH (r = −0.68, −0.67 and −0.64, respectively). 1-month r were less congruent. 3 short covid-19 surge episodes with Rt ∼ 1.5 happened on Jul 16, Sep 26 and Nov 7 (a, b, c). Each one was preceded by an episode where AH attained very low values: 2.8 g/m^3^ on Jul 7, 2.4 g/m^3^ on Sep12 and 1.6 g/m^3^ on Oct 29, respectively. Note that these events would correspond within realistic, but different ICR delays (9, 14 and 9 days respectively). Also note that there were 2 other dry episodes during the study period (on Aug 2 and Oct 5) that were not followed by comparable Rt peaks.

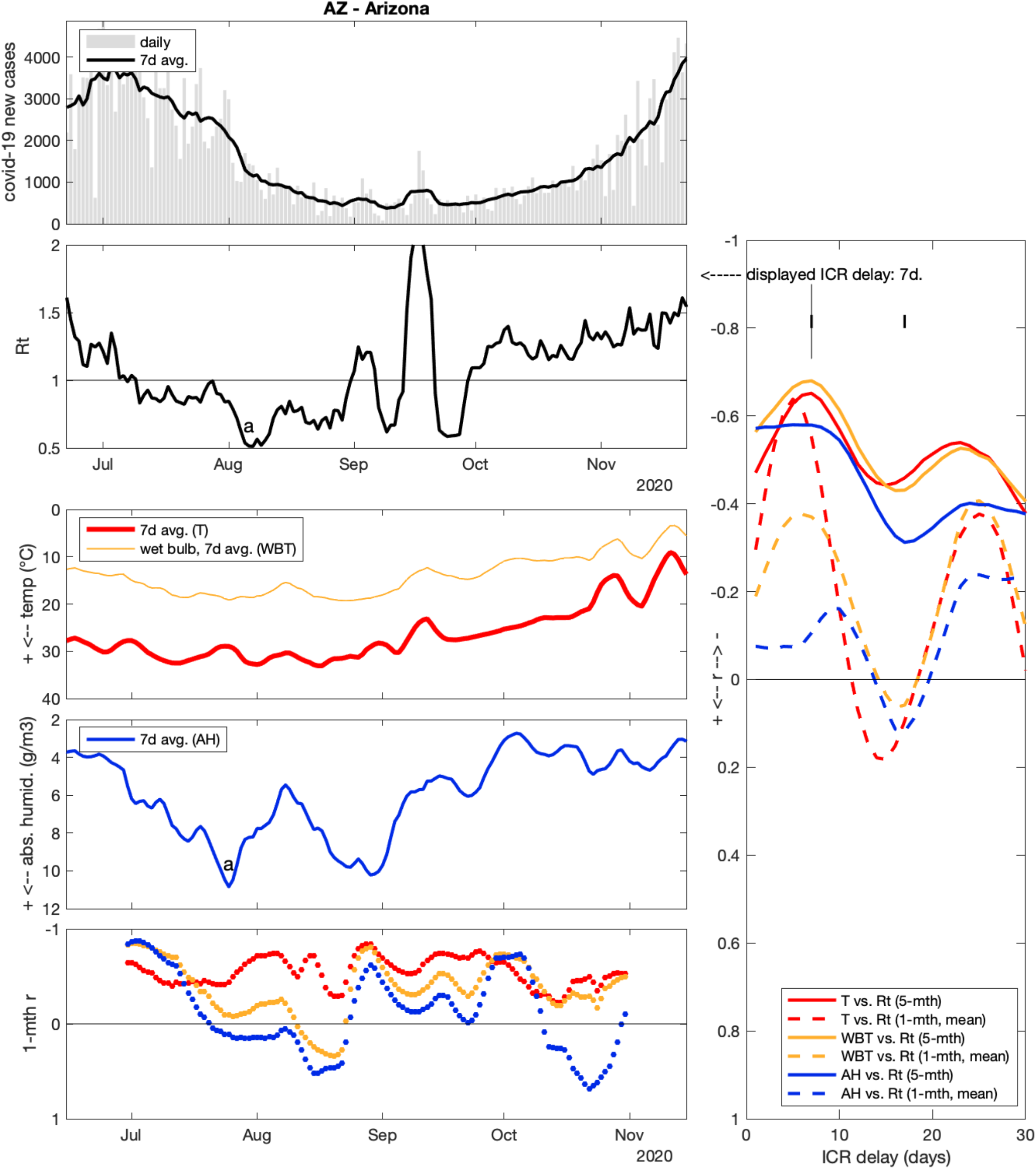

### Arizona (AZ)

Strongest 5-month correlation was observed between Rt and WBT (r = −0.68 for ICR = 7 days, a barely realistic short delay). Short-time correlations were not congruent, suggesting even shorter ICR delays for T and WBT. There was a very short-lived and high Rt = 2.1 peak on Sep 16 that could correspond to a cold peak (WBT = 12°C on Sep 12), but the 4-days implied ICR delay, although not impossible, is considered unrealistic. More convincingly, the highest AH over the whole period in AZ (11 g/m^3^) was reached on Jul 25 (a), followed 12 days later by the lowest Rt (0.5 on Aug 6). I found no other sharp graphic association between weather and case count data in AZ.

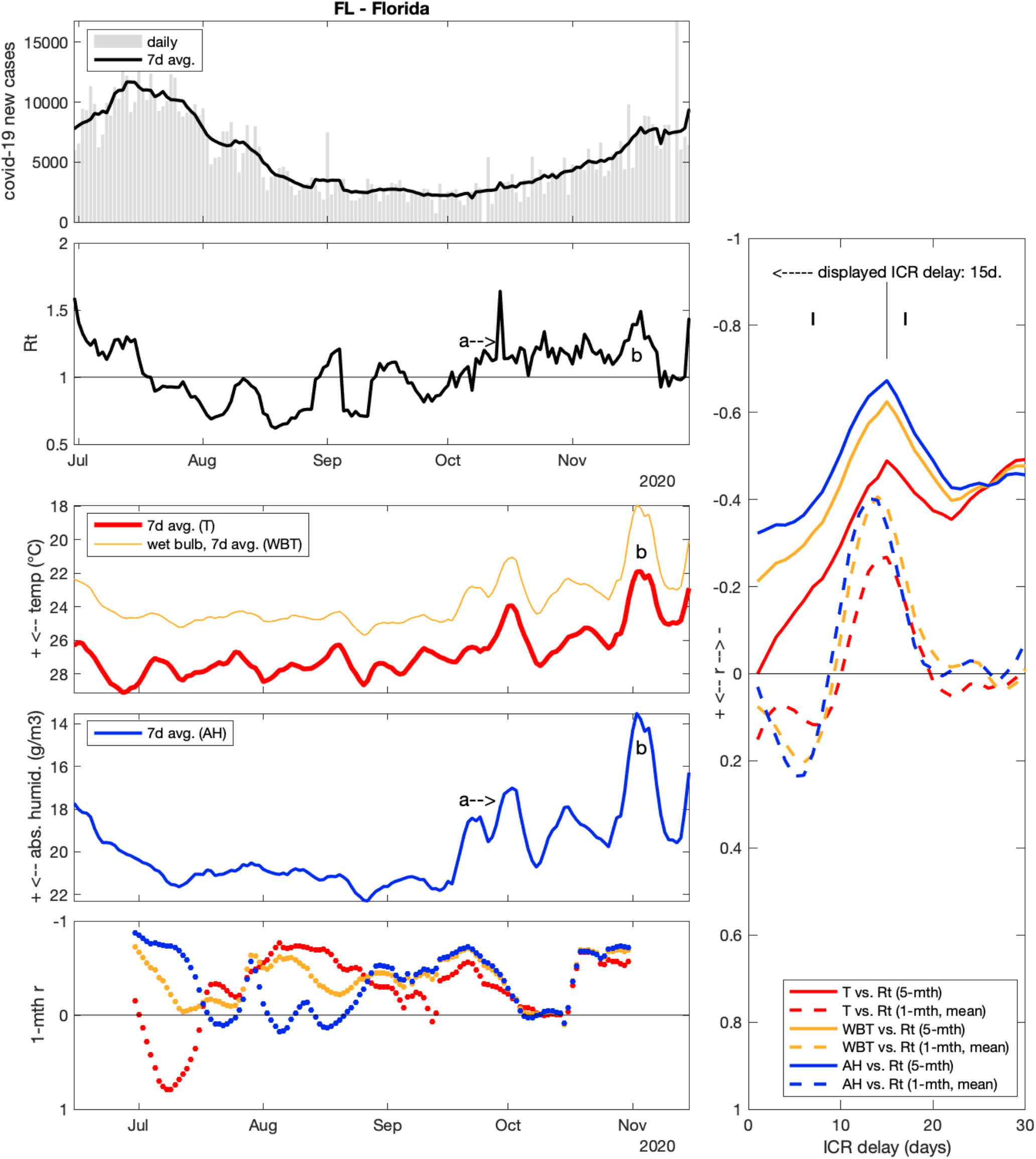

### Florida (FL)

Long- and short-time correlation data were congruent in FL, suggesting a rather long ICR delay of 14-15 days. Stronger 5-mth correlation was observed for AH vs. Rt (r = −0.67). There unfortunately are a few “spikes” in FL new case counts that make Rt series noisier than average. Still, I note the following events: after the summer decrease, case started to increase again from the first week of October (a), with Rt remaining close to 1.2 for the following 7 weeks. This could correspond to AH dropping durably below 20 g/m^3^ around Sep 19 (i.e. about 15 days earlier). Also, a further transient drop (b) of AH down to 14 g/m^3^ (and T down to 22°C) around Nov 2 was followed by a peak of Rt up to 1.5 on Nov 18, i.e. 16 days later.

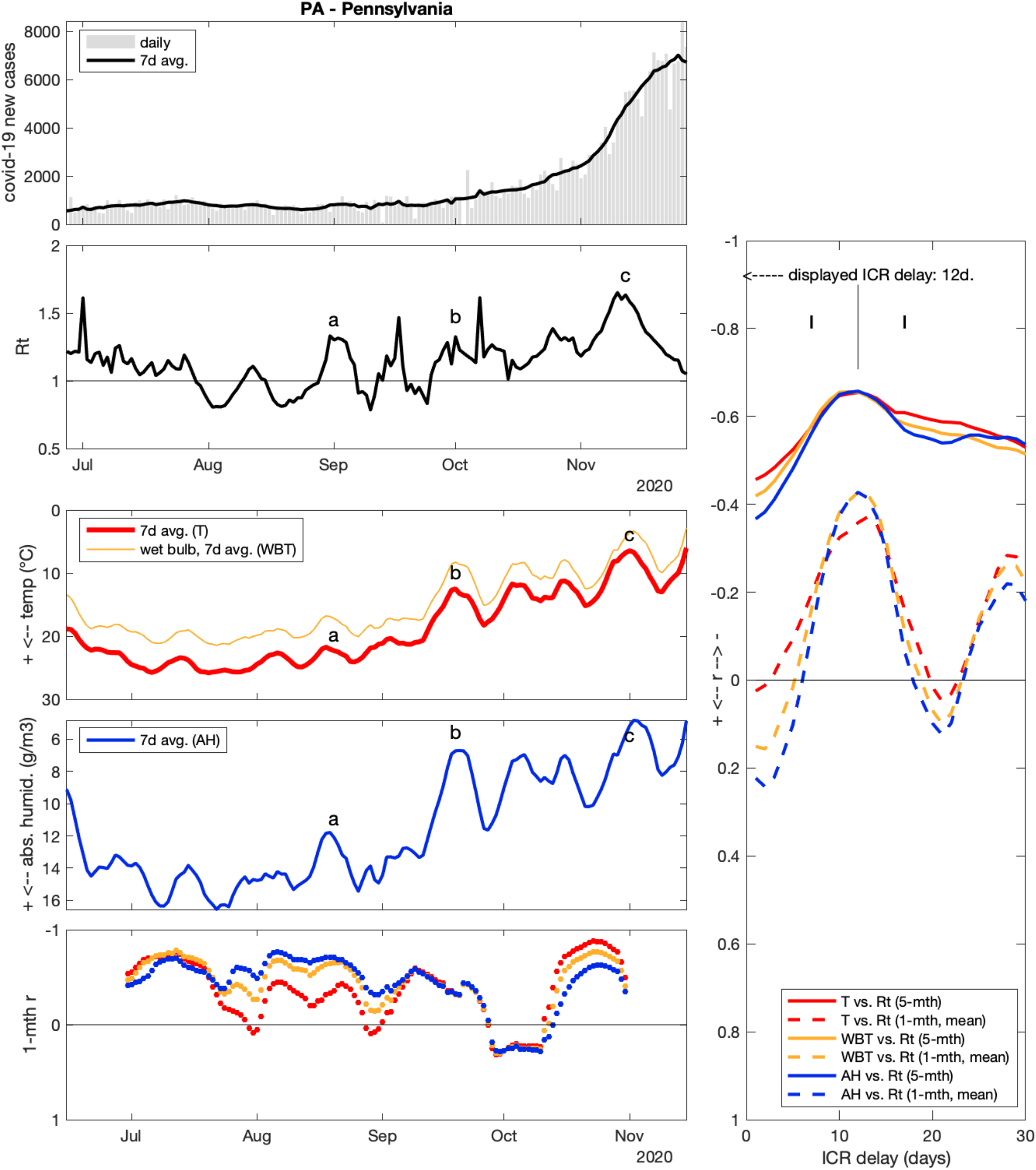

### Pennsylvania (PA)

T, WBT and AH had equal 5-month r = −0.65 for ICR delay = 11-12 days, and the peaks in 1-month r were congruent. As for previous states, to illustrate graphical translation of correlation results, the association between 3 cold-dry events (a, b, c) and Rt peaks is proposed on figure. The last of these Rt peaks is the strong case surge of November in PA, with cases increasing from ∼2200 to ∼7000 (Rt = 1.7 on Nov 10).

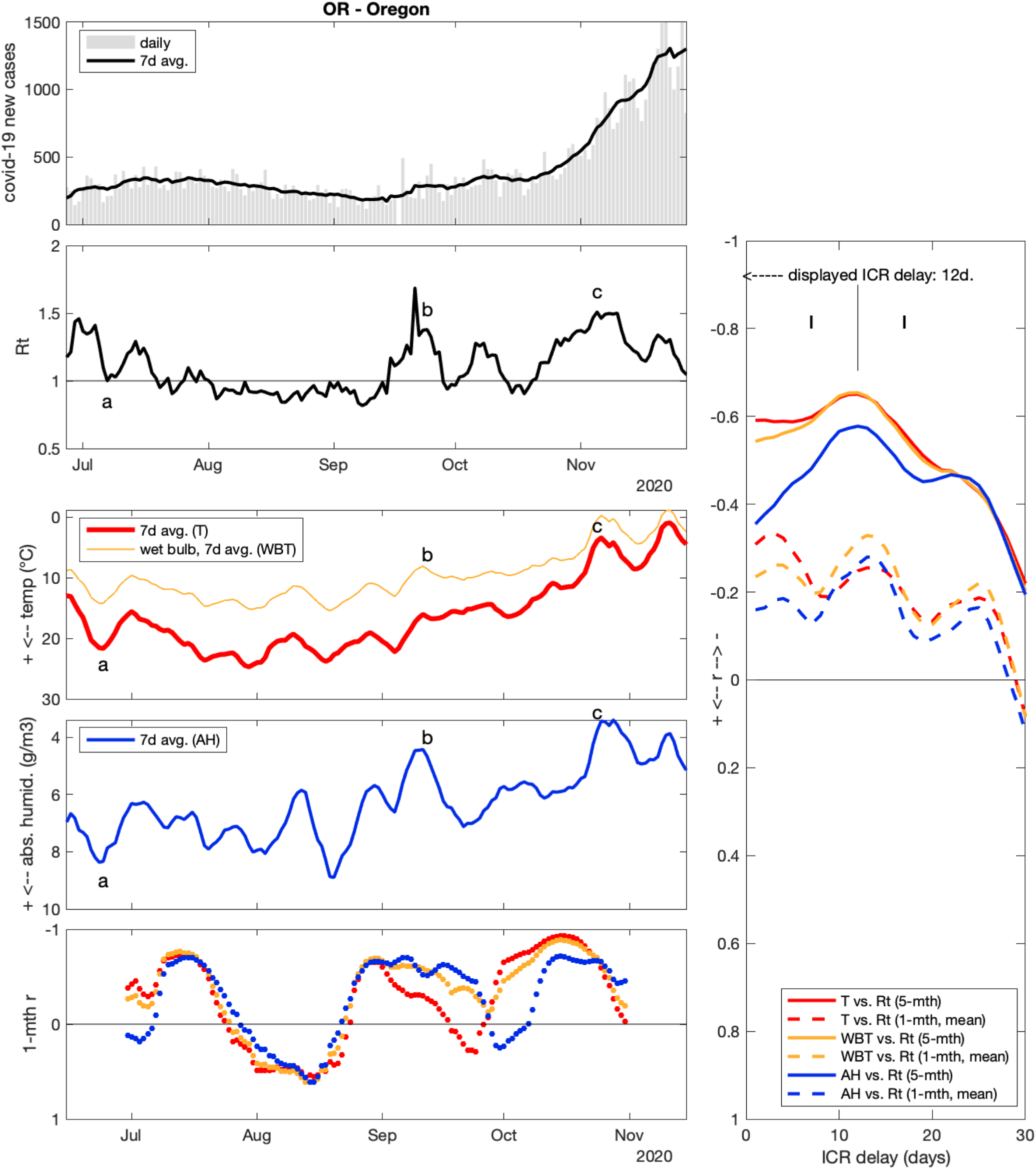

### Oregon (OR)

WBT and T had the strongest 5-month correlation with Rt (r = −0.65), for ICR = 12 days again. AH had a slightly weaker correlation overall (r = −0.58). 1-month correlations results were less clear, with r-value oscillations. I proposed 3 weather-Rt associations on figure (a, b, c), the last one corresponding to a large case surge in November. On the other hand, it is noteworthy that OR saw significant weather variations during August, that do not seem to correspond to any strong Rt variations.

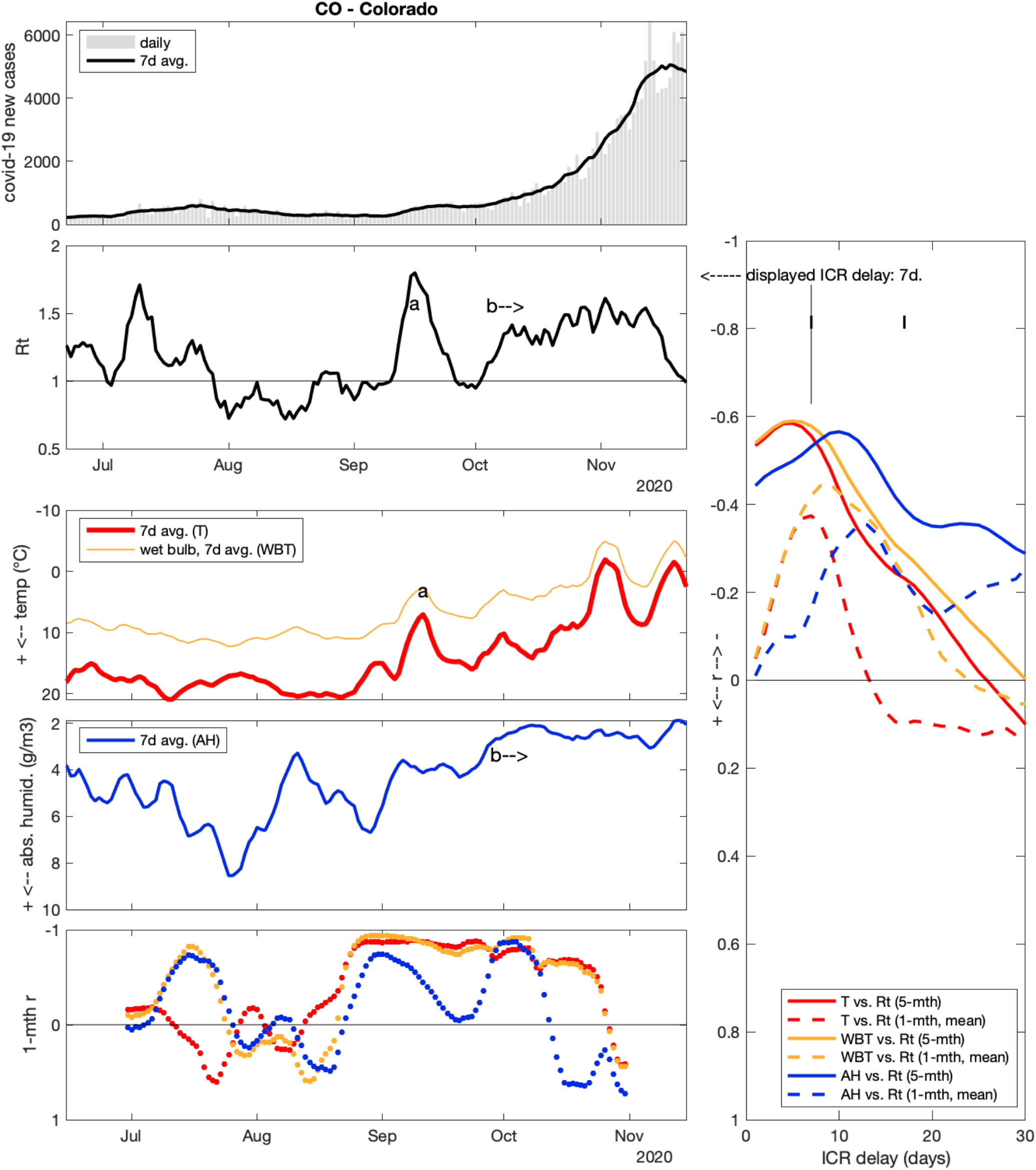

### Colorado (CO)

Rt vs. T or WBT correlations suggested an ICR of 6 days (r = −0.6), which is considered unrealistically short. AH correlation rather suggest ICR = 10 days (r = −0.57). Among noteworthy episodes, there was (a) a surge around mid-September (Rt = 1.8 on Sep16), with cases surging from ∼250 to ∼600. This could correspond to a cold anomaly around Sep 11, but again 5 days is a very short delay. More convincingly, the Oct-Nov prolonged surge from ∼600 to ∼5000 daily cases, with Rt settling near 1.4-1.5 for 6 weeks, could correspond to AH falling below 3 g/m^3^ from Sep 28 (b).

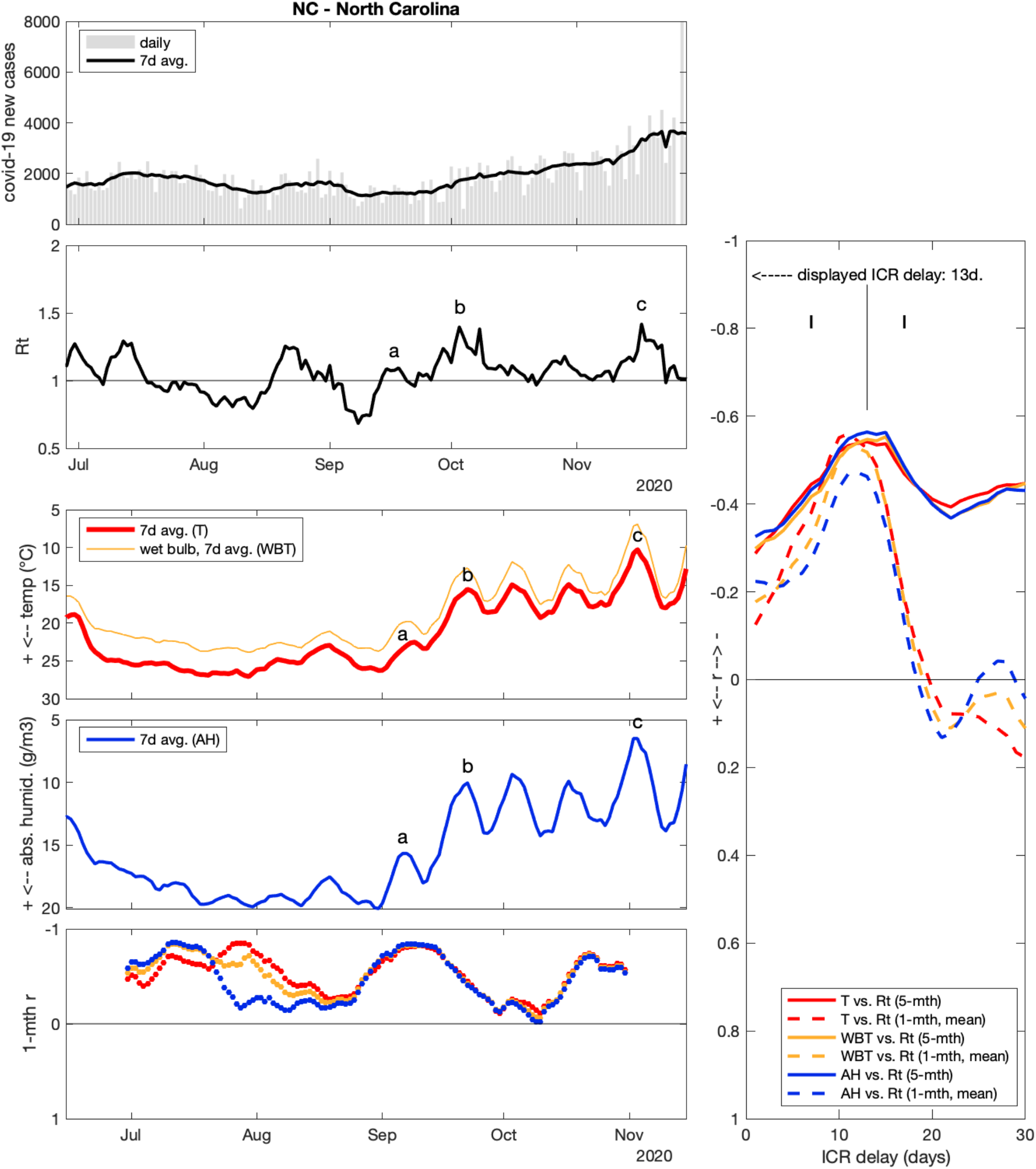

### North Carolina (NC)

Rt vs. AH returned the strongest 5-month correlation (r = −0.56 for ICR = 13 days), while mean 1-month correlation was strongest for T (r = −0.56 for ICR = 11 days).

3 peak correspondences are proposed on figure (a, b, c).

(Note: an isolated spike of 6142 new cases on Sep 25, probably from backlog reporting, was removed from NC analysis because it caused a strong Rt peak which disrupted correlation results).

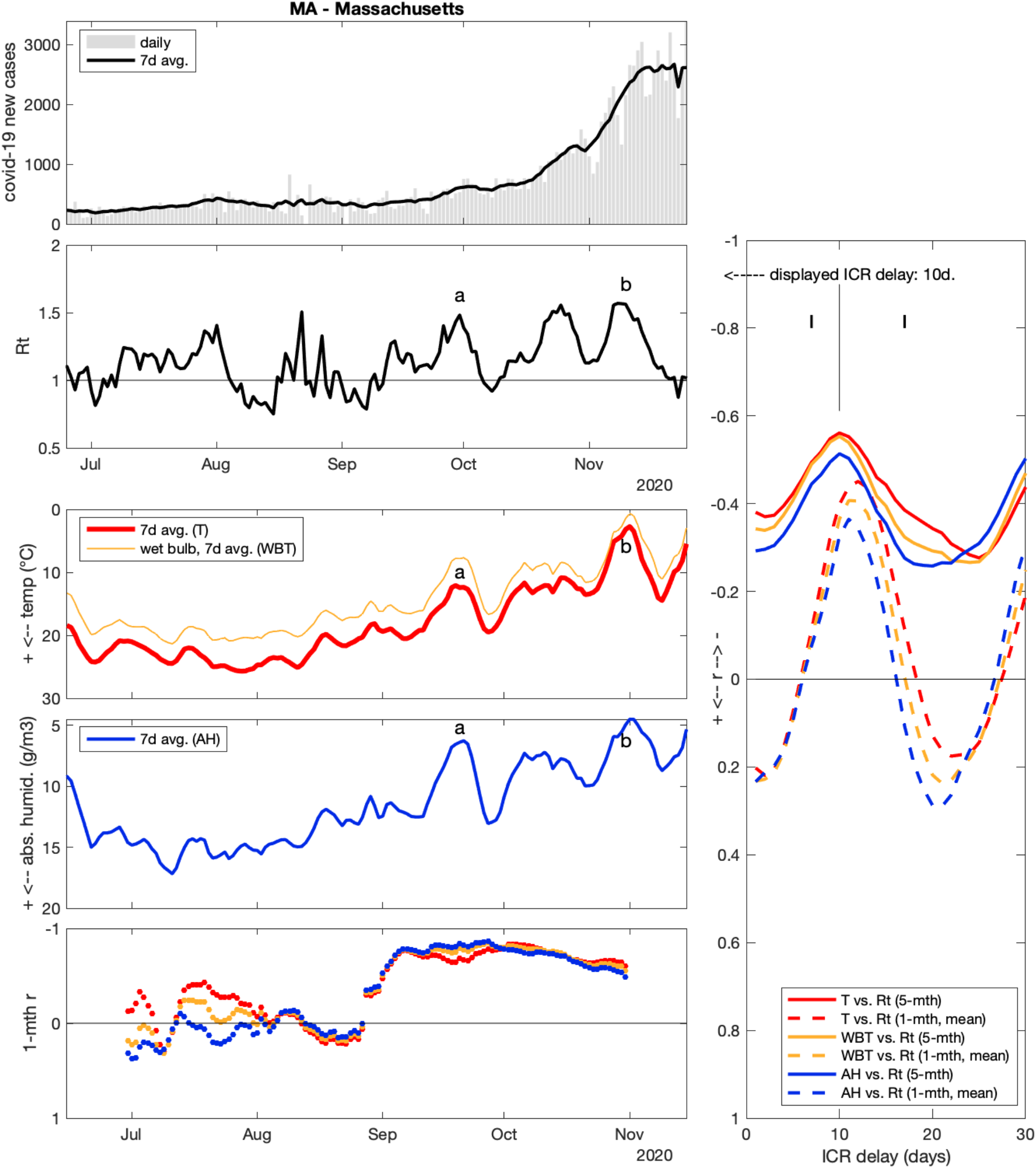

### Massachusetts (MA)

T was the weather variable most strongly correlated with Rt (r = −0.56 for ICR = 10 days). Mean 1-month r rather suggest ICR = 11-12 days, and 1-month r were weak to null during the summer (see lower-left panel). 2 peak associations are proposed on figure (a, b).

(Note: a negative spike of −7757 new cases on Sep 25, most probably caused by state data correction, was removed from MA analysis).

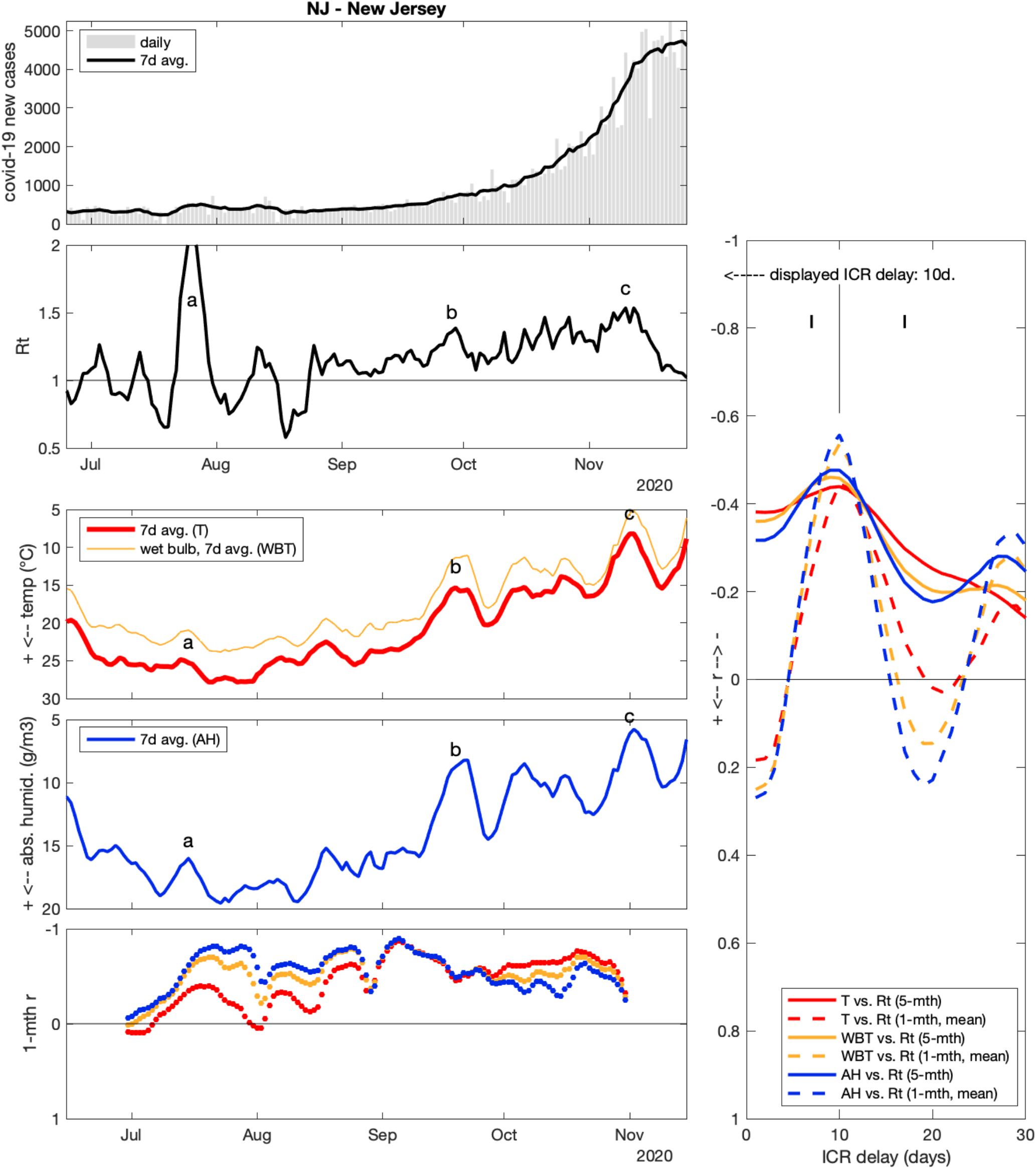

### New Jersey (NJ)

in NJ, peak short-term (1-month) correlations were stronger than peak long-term (5-month) correlations, indicating that cold and/or dry episodes tended to correspond to Rt peaks, but that the weather change over the study period (U-shaped) did not match Rt trend so well. Correlation was strongest for AH vs. Rt (mean 1-month r = −0.56, ICR = 10 days). 3 peak associations are proposed on figure (a, b, c).

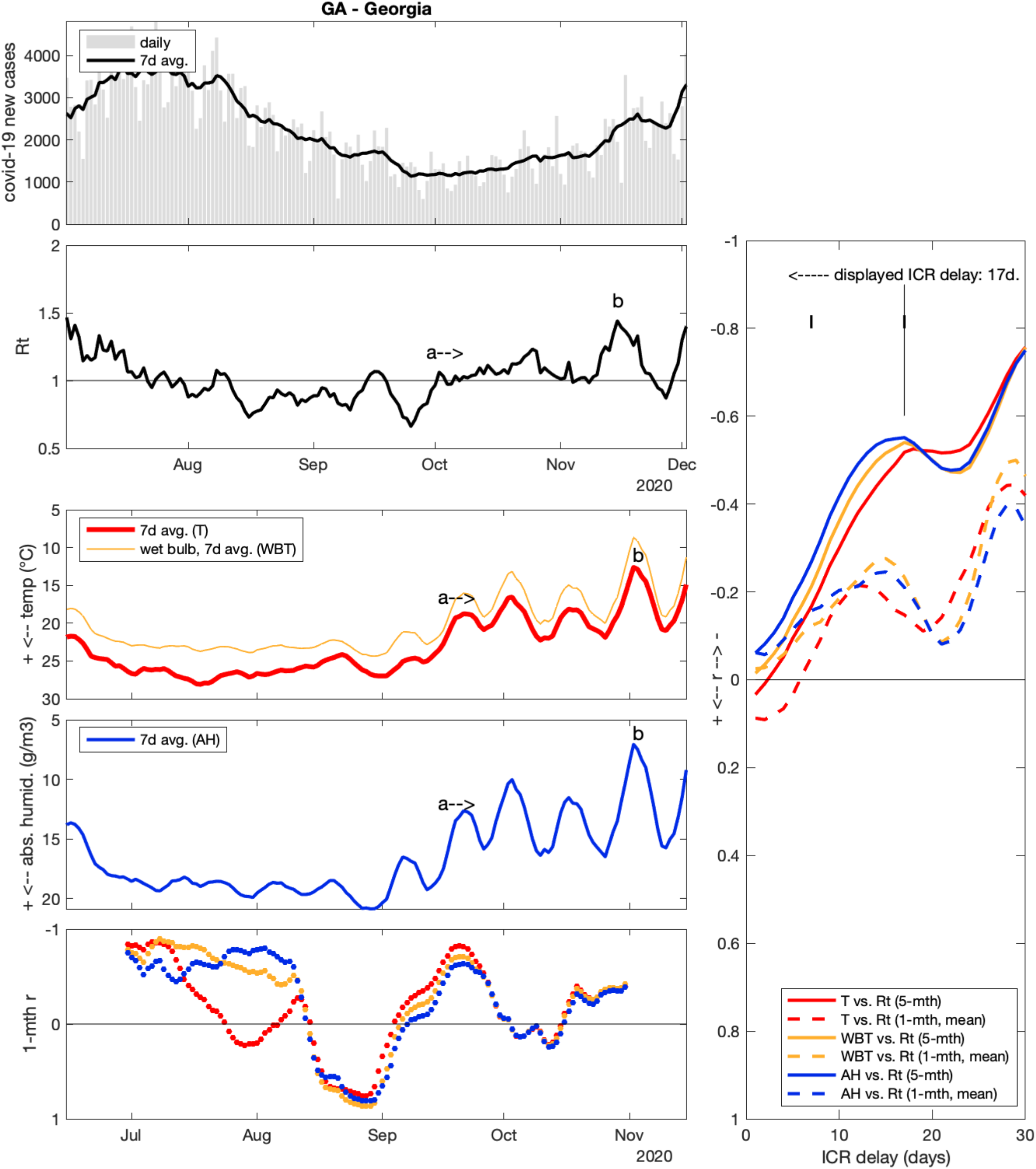

### Georgia (GA)

AH had the strongest correlation with Rt (r = −0.55), with a long ICR of 17 days. Even stronger r could be attained for time-shifts > 25 days, but these are considered unrealistic infection-to-case-report delays. In GA, most of the summer saw a decrease in Rt after a large surge started in June. AH and WBT increased in parallel. The dynamics changed around Oct 2, with Rt re-increasing to 1 and remaining stable for 5 weeks (a). This could correspond to T and AH stepping down from Sep 17 (below 22°C and 16 g/m^3^, respectively). Dynamics changed again with a new surge (b) culminating on Nov 16 (Rt = 1.4), which coincides with a cold-dry episode around Nov 2 (i.e. 14 days earlier).

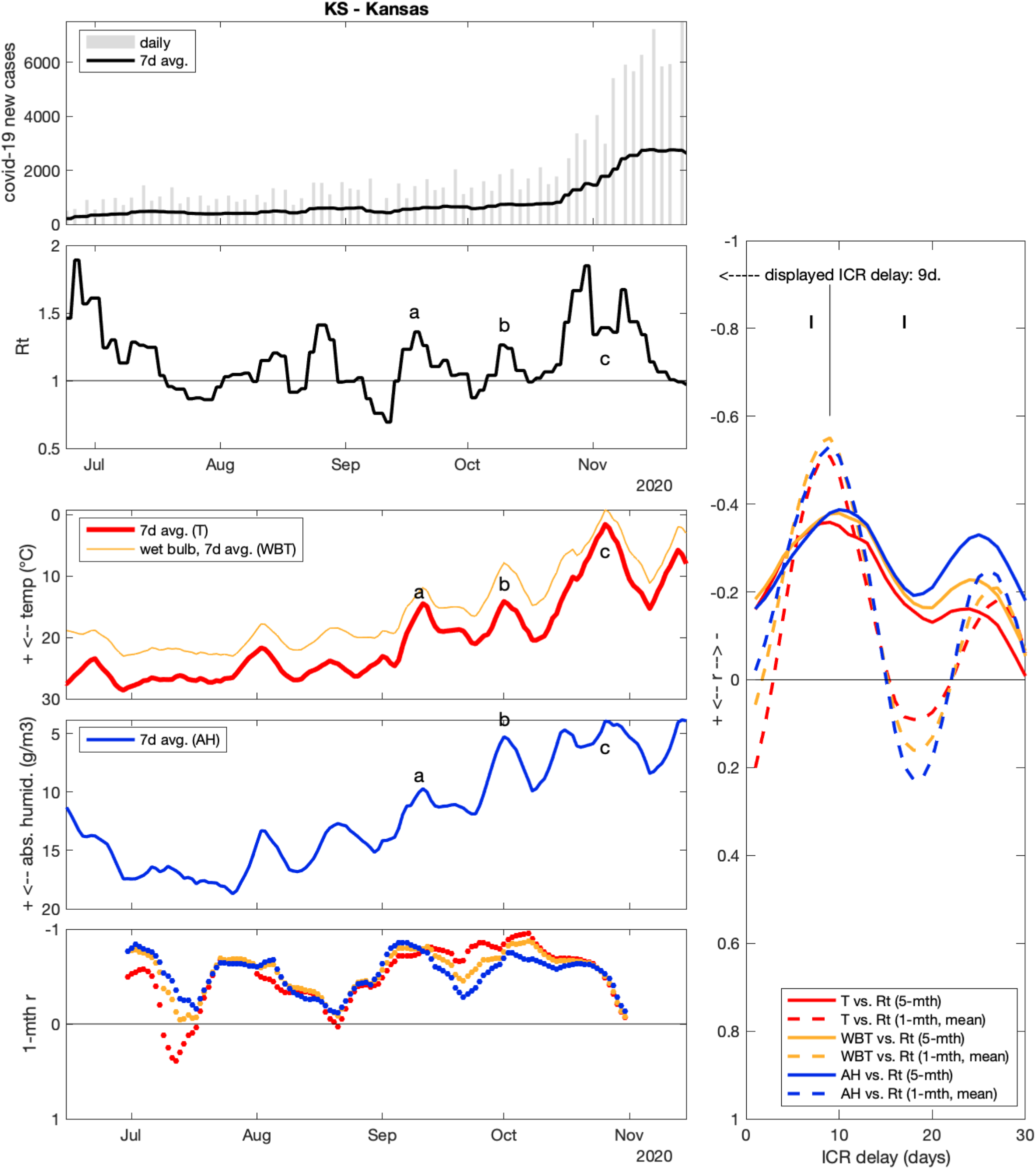

### Kansas (KS)

As is the case for NJ, 1-month r attain more negative values than 5-month r (strongest correlation WBT, mean 1-month r = −0.55 for ICR = 9 days). 3 peak matches are proposed to illustrate correlation results (a, b, c).

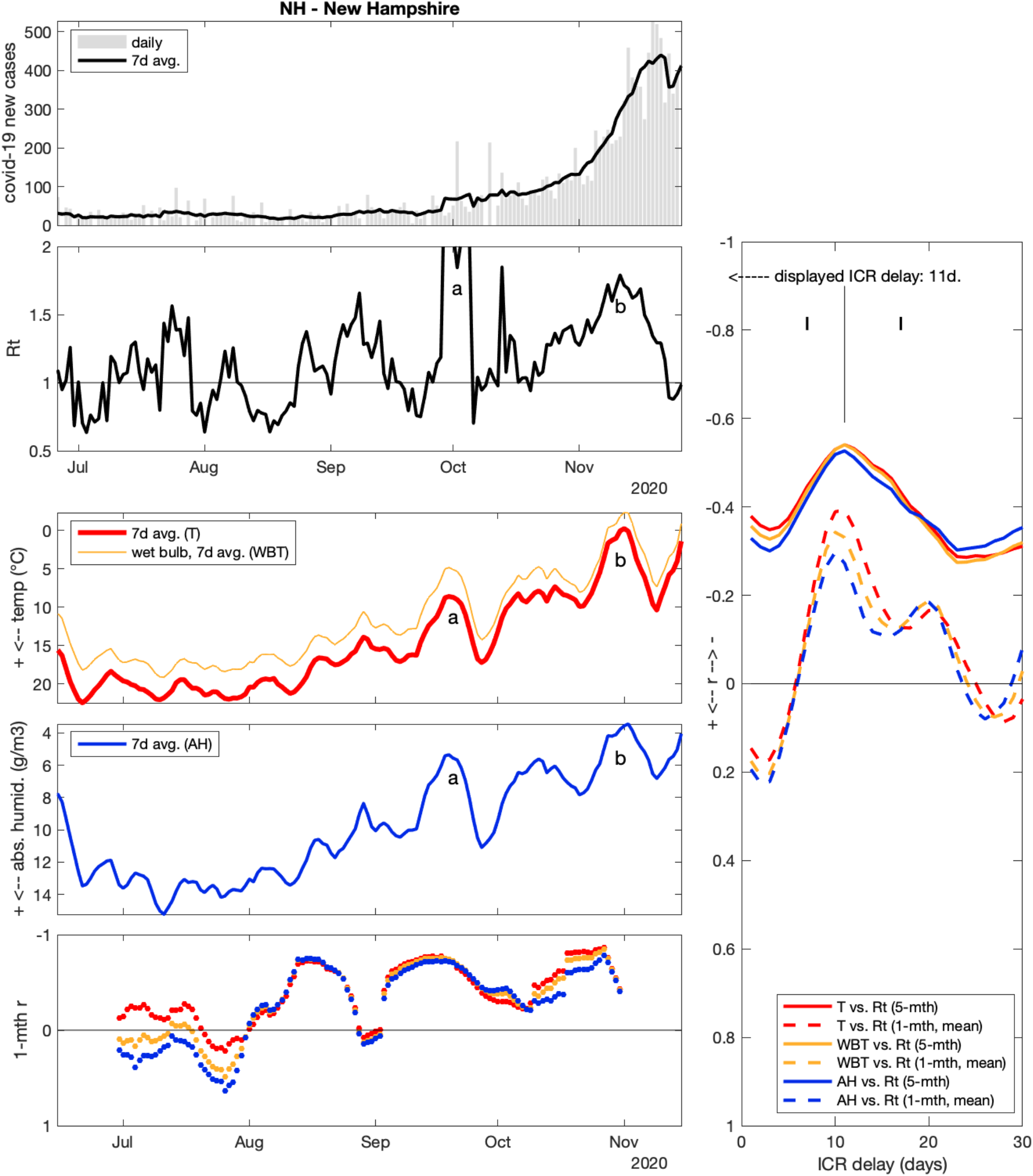

### New Hampshire (NH)

New case counts are very low in NH, resulting in noisier Rt than in other states. However, correlations congruently indicated an ICR delay of 11 days (r = −0.54 for T, with very similar values for WBT and AH). 2 main peak correspondences are proposed (a, b).

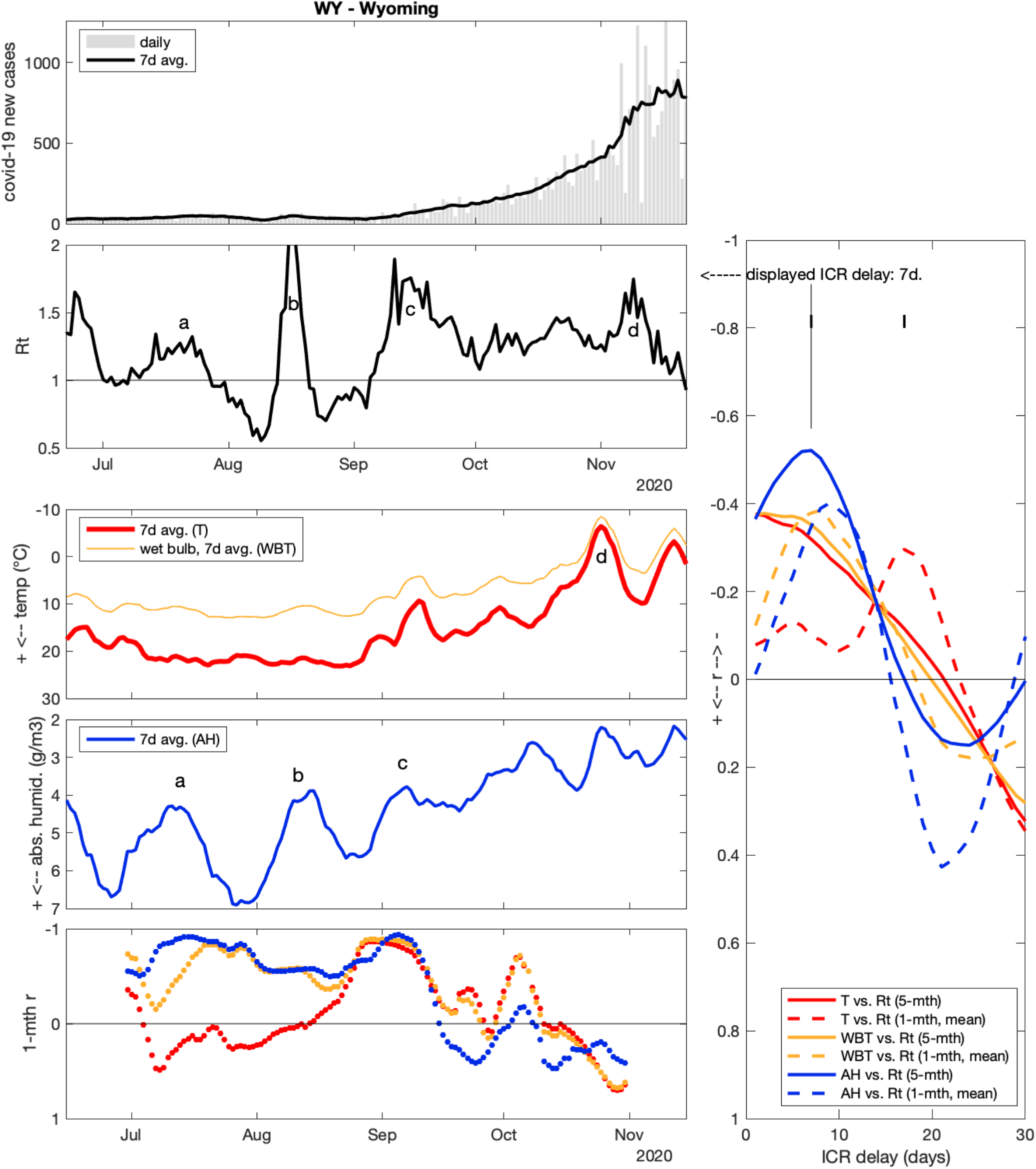

### Wyoming (WY)

Correlations for the 3 weather variables were not congruent in WY. T and WBT did not exhibit any peak for 5-month correlation, and 1-month correlations suggested different ICR depending on the considered variable. A stronger correlation was observed for AH (5-mth r = −0.52 for ICR = 7 days). When plotting data, parallelism can be observed between AH and Rt during the summer (a, b, c), but less so after Sep 15. Later, a peak in Rt around Nov 9 could be linked to a cold episode around Oct 25, but with a longer assumed ICR delay of 15 days (d).

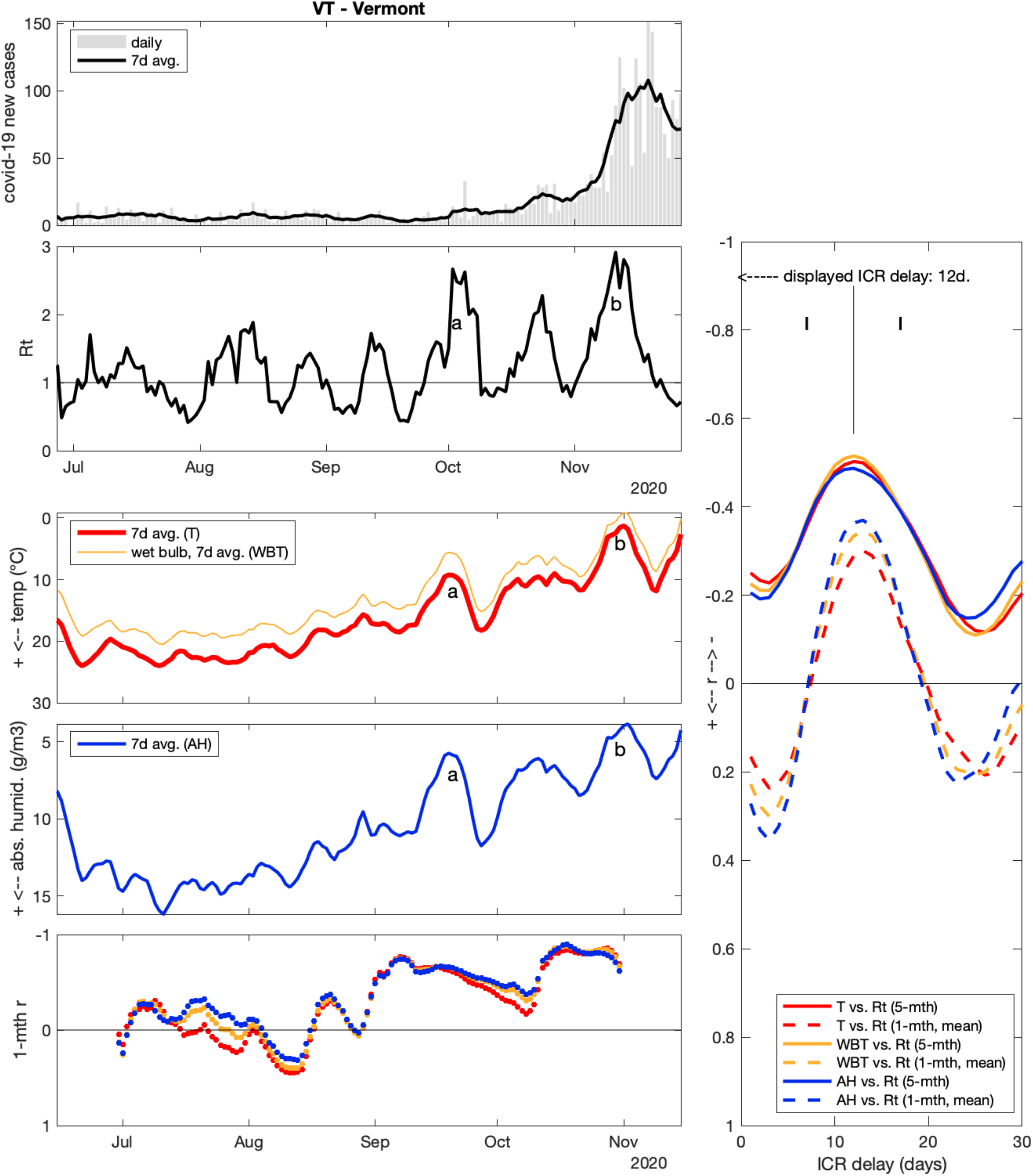

### Vermont (VT)

Cases counts were even lower in VT than in NH, resulting in Rt with larger volatility (hence the enlarged Rt scale for this state). However, there was good congruence between short and long-term correlations when assuming an ICR of 12-13 days. 2 main peak associations are proposed (a, b). These are essentially the same cold-dry events as in neighbor states of NY and NH.

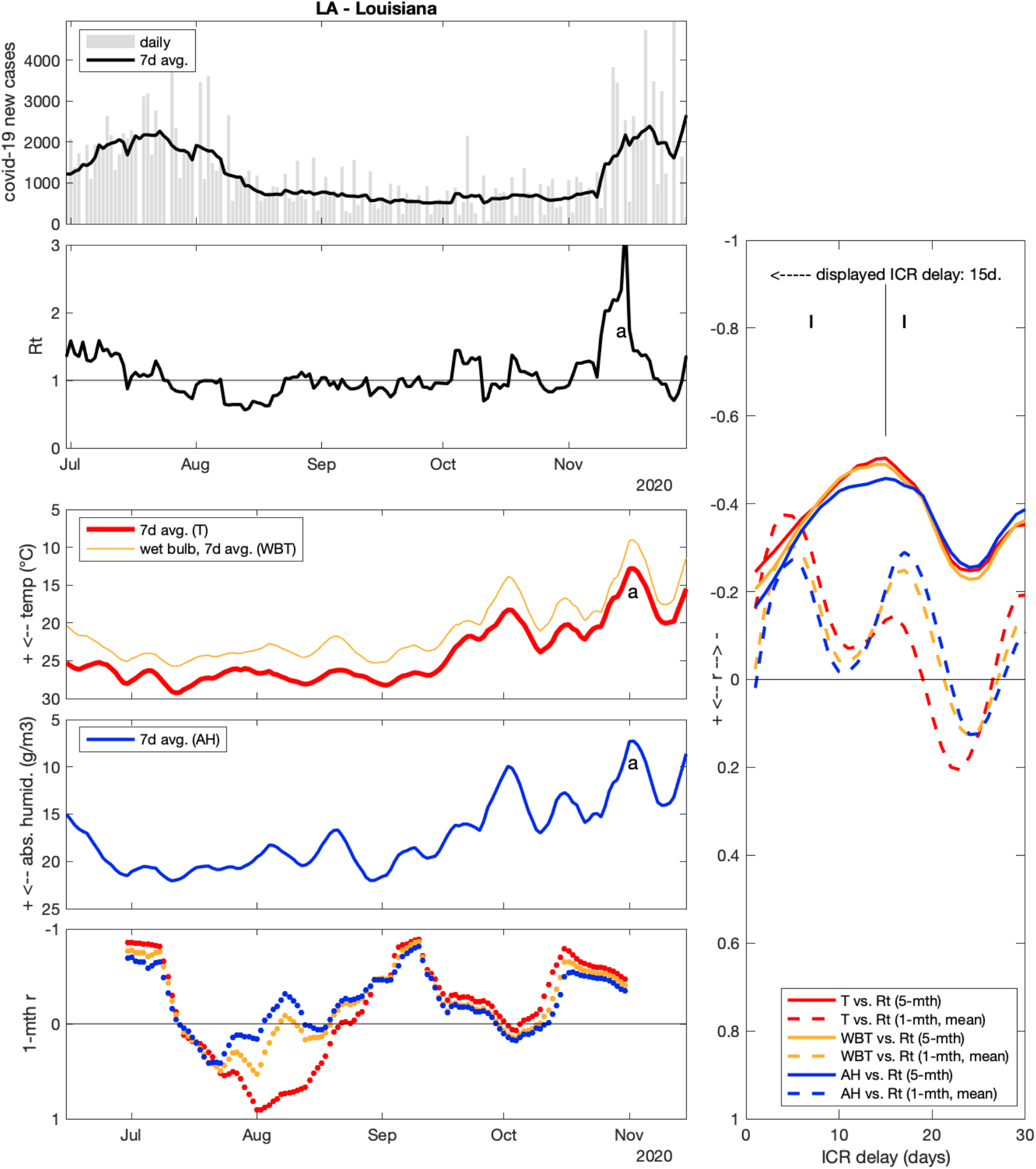

### Louisiana (LA)

5-month r between Rt and T attained −0.5 for ICR = 15 days, indicating good match between overall (U-) shape variation of both variables. The decrease of Rt after the June-July surge was paralleled by T increasing until mid-July (but not after that, whereas Rt decreased further until mid-August). More recently, there was a rapid case surge in LA, culminating at estimated Rt = 3.4 on Nov 15 (daily cases increased from ∼800 to ∼2200 “only” because the Rt peak was short-lived), that can be related to a cold event on Nov 1. However, an older cold episode around Oct 2 did not seem to affect Rt values (Rt scale enlarged for this state).

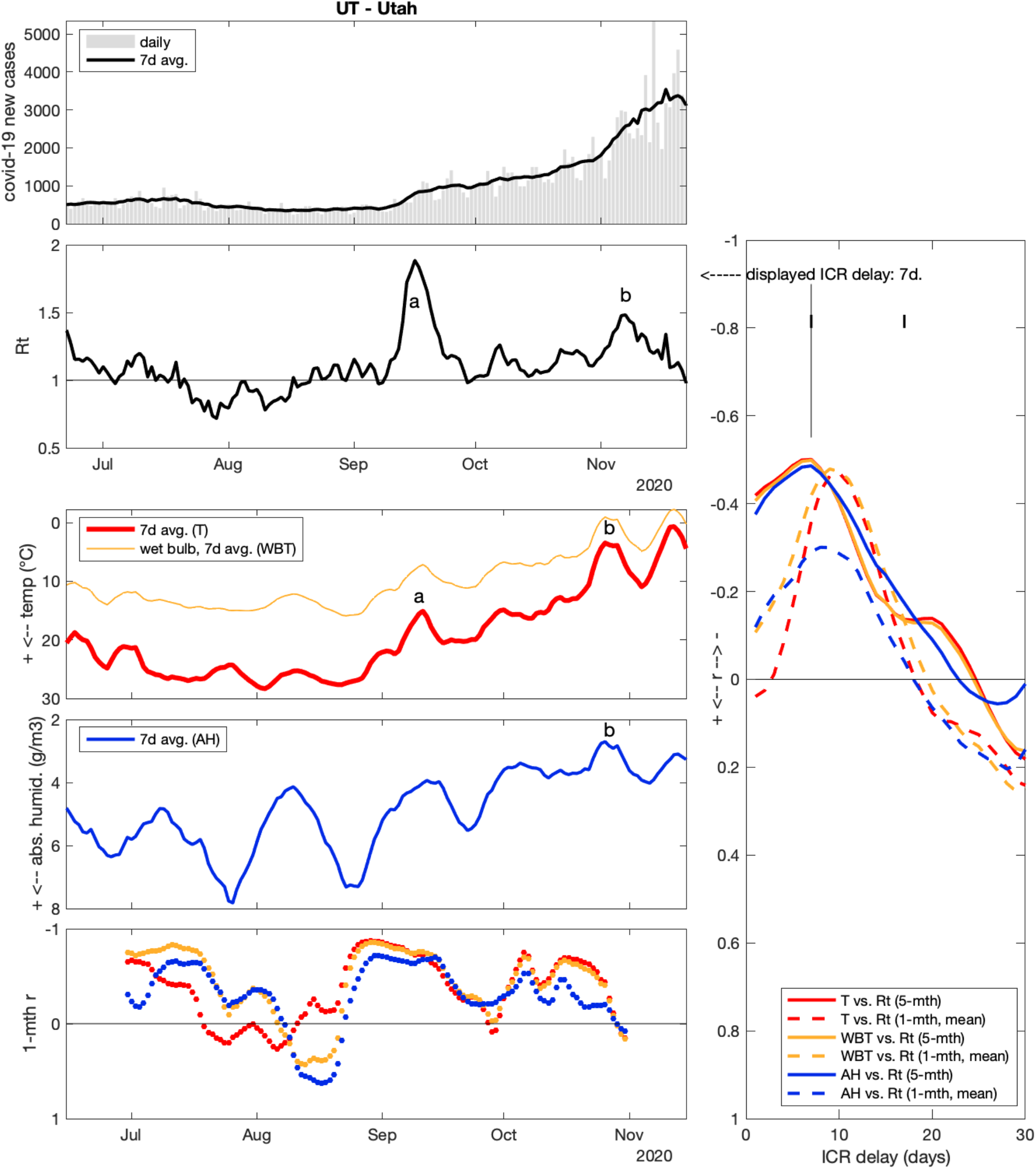

### Utah (UT)

In UT, 5-month r attained −0.5 for correlation of T vs. Rt, assuming an ICR of 7 days. Short-term correlation rather suggested an ICR close to 10 days. 2 main surges happened in UT: the first (a) around Sep 16 (Rt = 1.9), that could be related to a cold event on Sep 11 (but with a short assumed ICR of only 6 days). The second surge (b) happened around Nov 7 (Rt = 1.5), possibly corresponding to another cold episode around Oct 26 (more realistic ICR = 12 days). There were important AH variation during the summer in UT that could not easily be related to corresponding Rt variations.

### 23 states with moderately negative weather-Rt correlations (−0.5 < r ≤ −0.2)

The weather-Rt correlations in these states was less strong than for previous states, but remain mainly congruent with previous results. When associations between weather episodes and Rt peaks were useful to illustrate correlation results, they were labeled in the following figures by letters (a-e). Note that letter alignment are for realistic ICR (12±5 d.), but as previously, were not constrained to respect the single ICR value chosen on the right panel, as effective ICR might vary through time for a given state.

(Supplementary note for TX: two strong spikes of 17820 and 71734 new cases, on Sep 22 and Nov 1, probably caused by backlog reporting, were removed from TX analysis because they caused strong Rt peaks which disrupted correlation results).

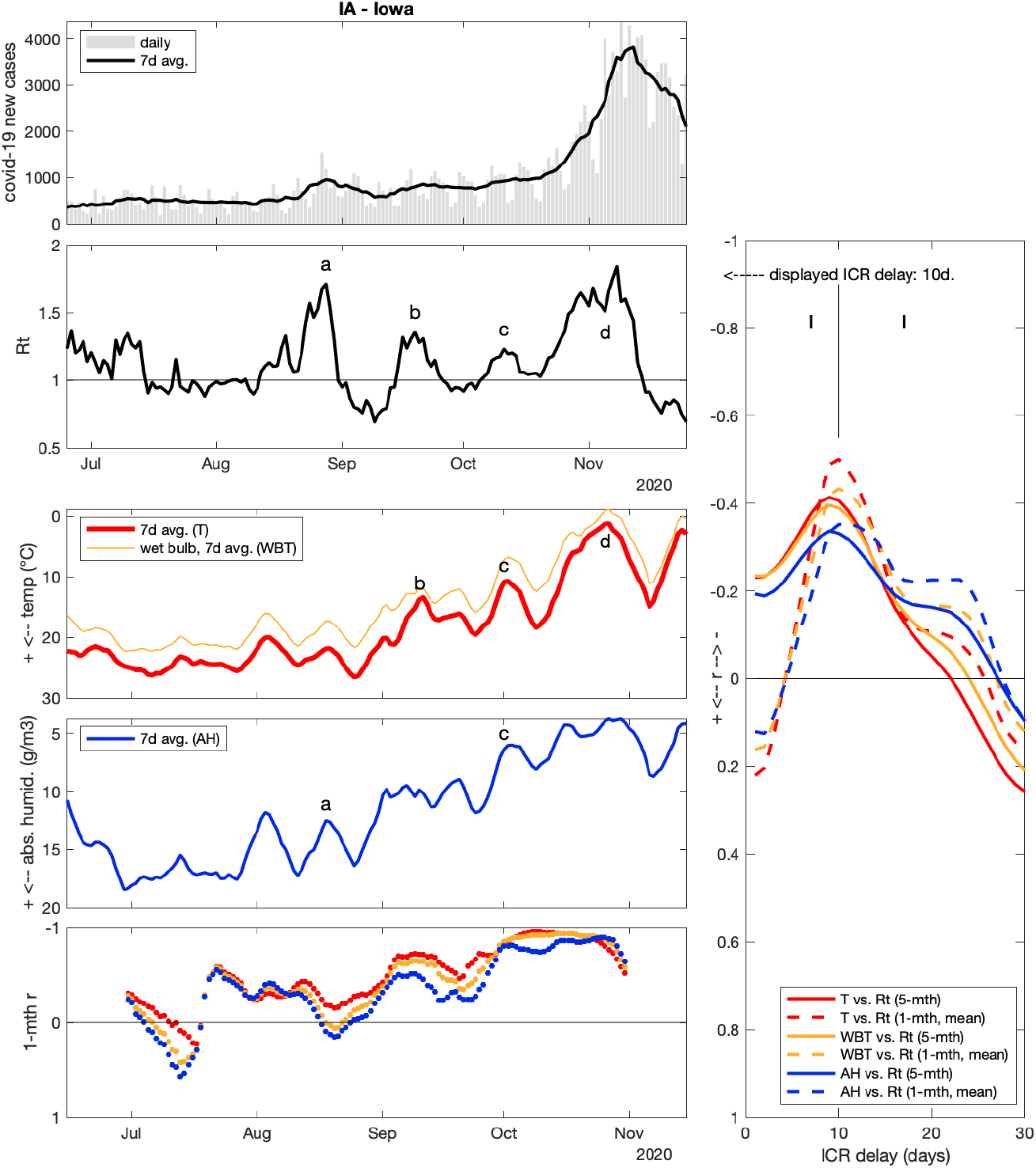

**Iowa (IA)** r > −0.5

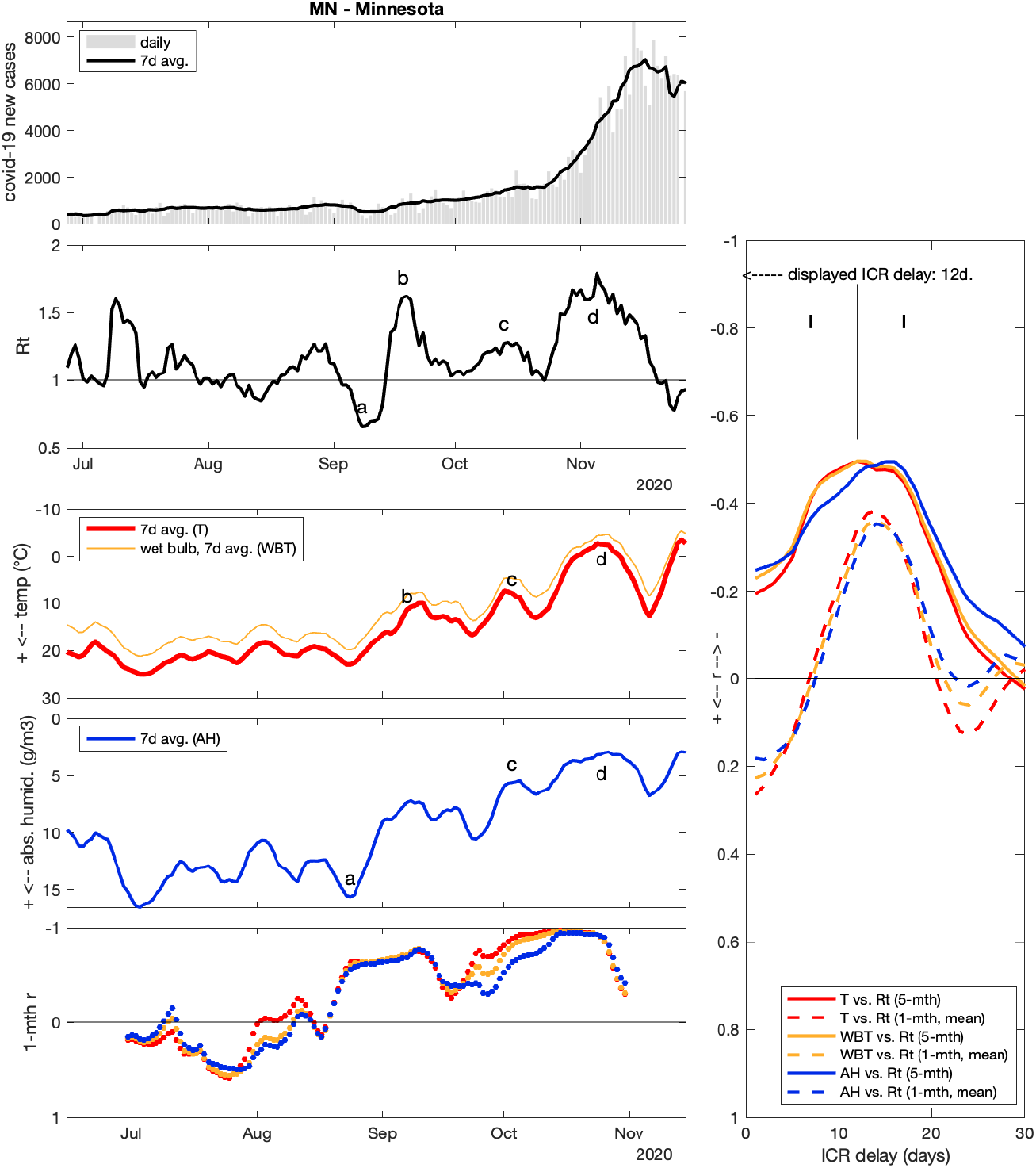

**Minnesota (MN)** r > −0.5

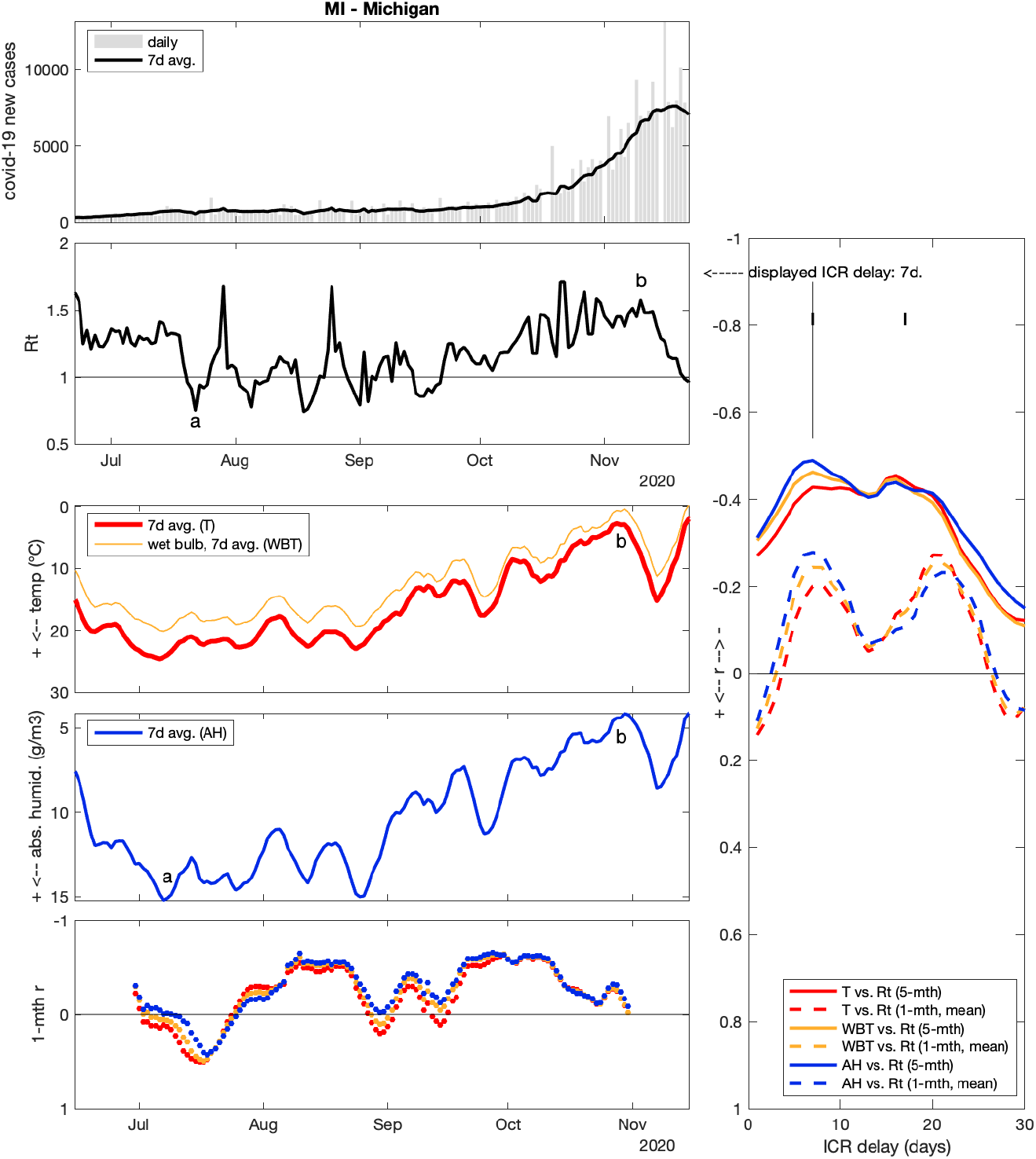

**Michigan (MI)** r > −0.49

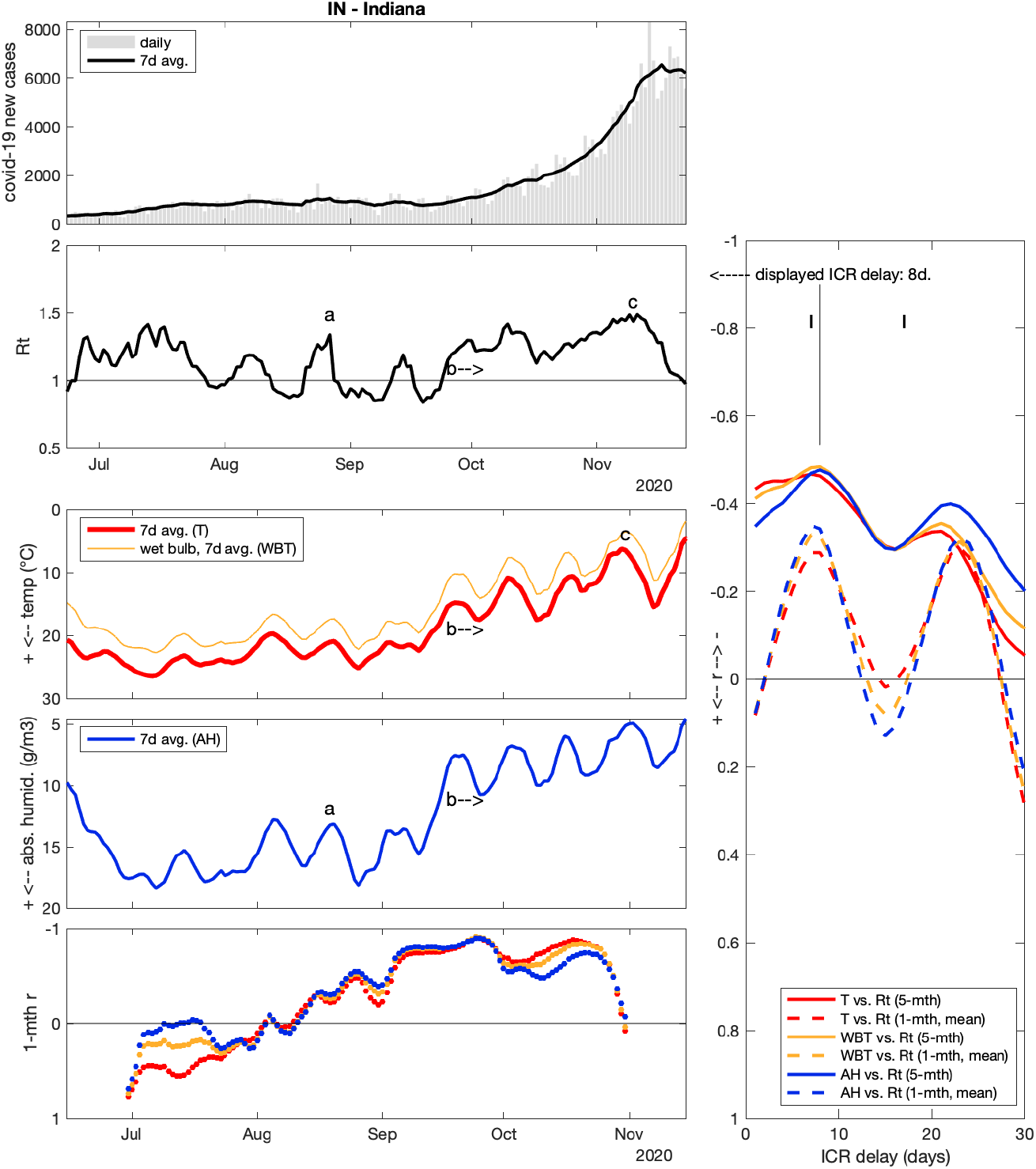

**Indiana (IN)** r > −0.49

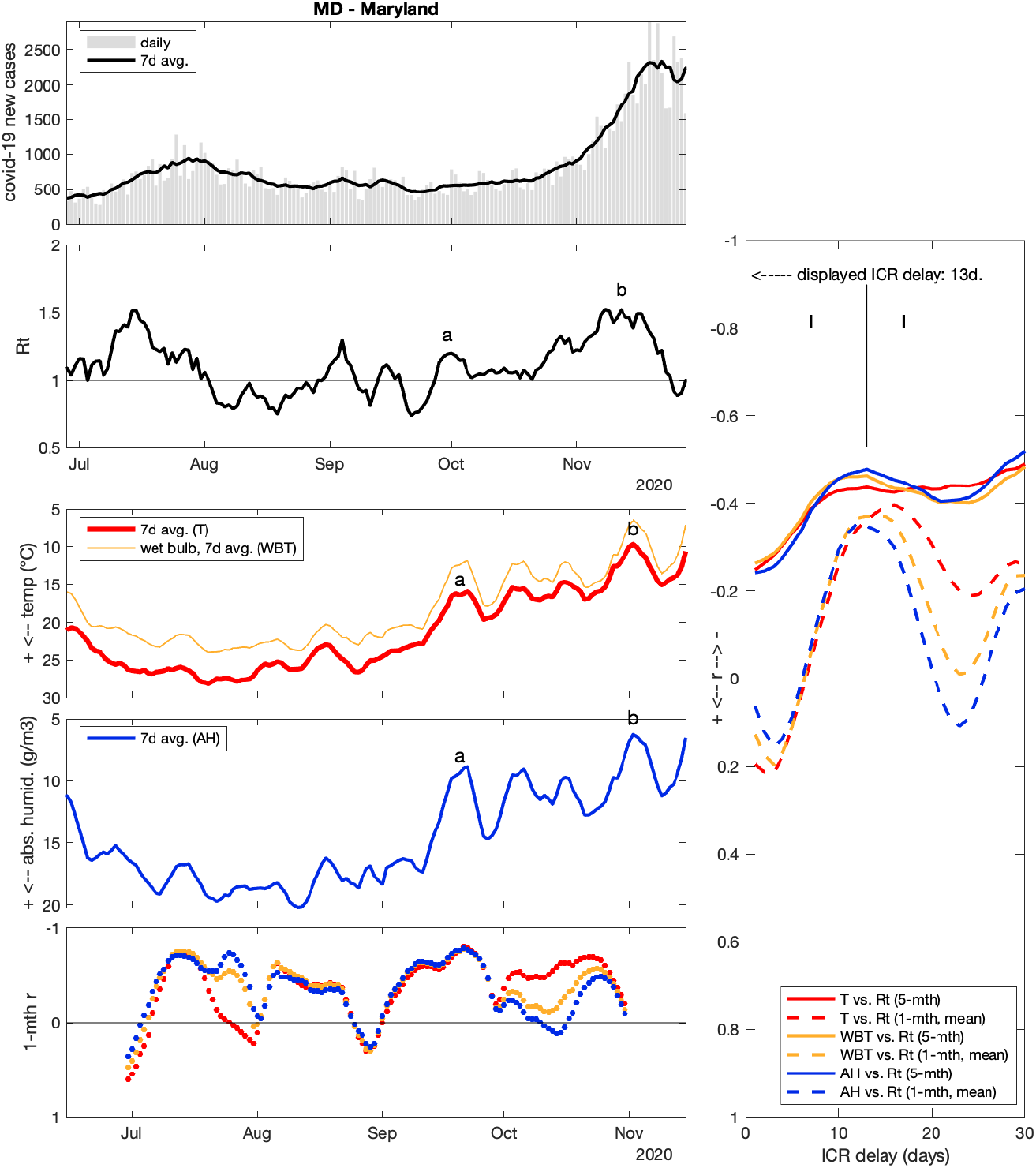

**Maryland (MD)** r > −0.48

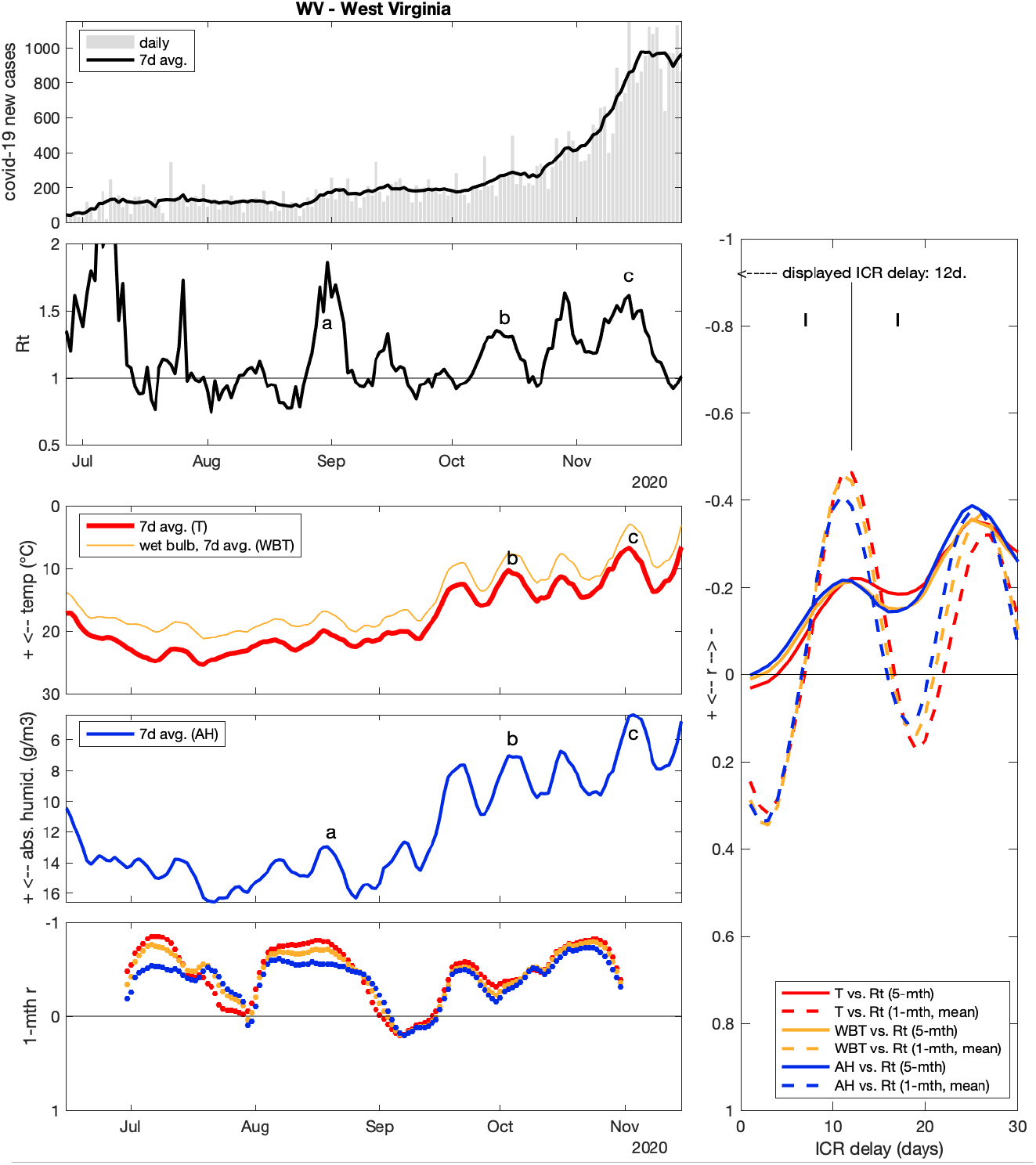

**West Virginia (WV)** r > −0.47

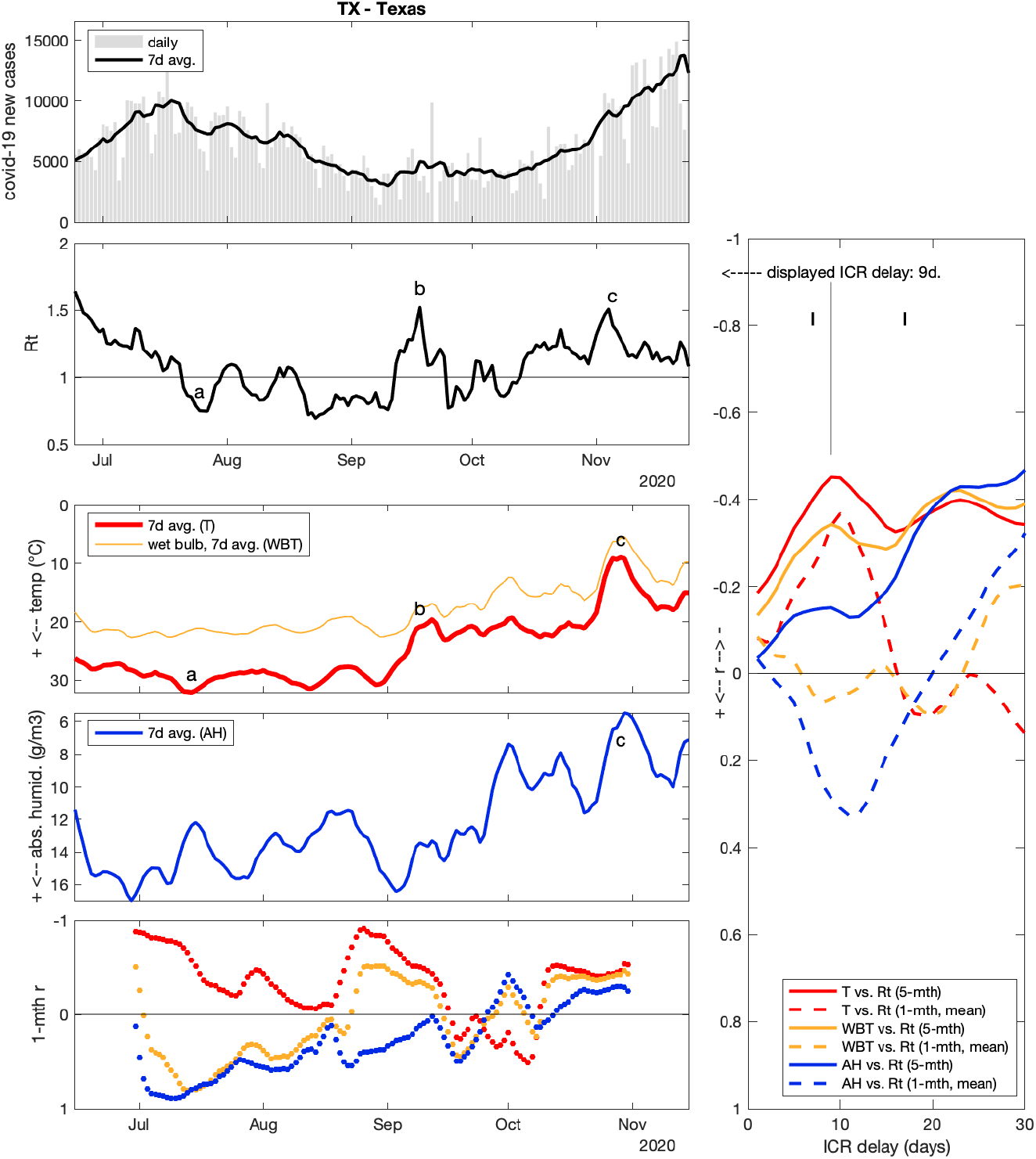

**Texas (TX)** r > −0.46

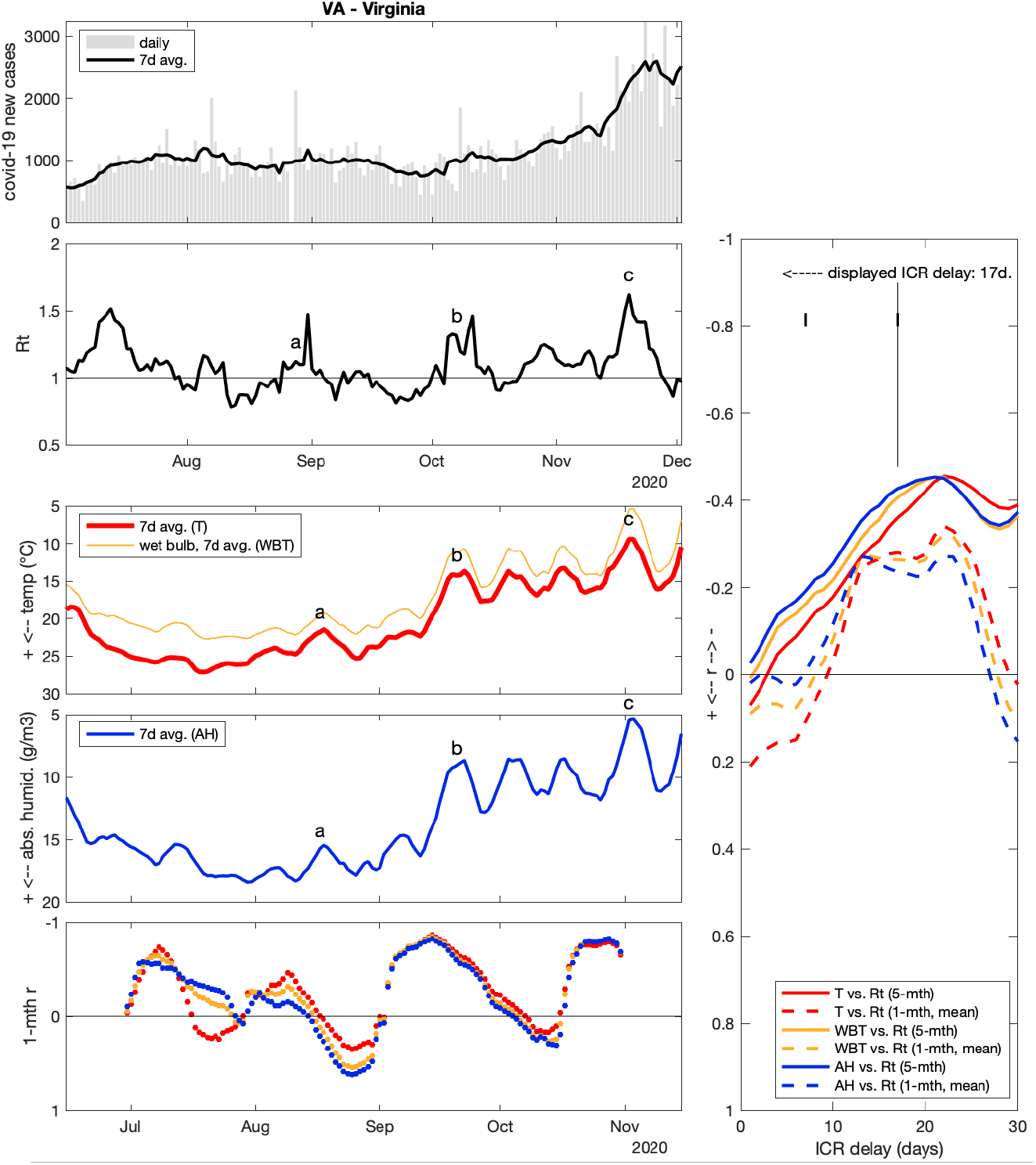

**Virginia (VA)** r > −0.43

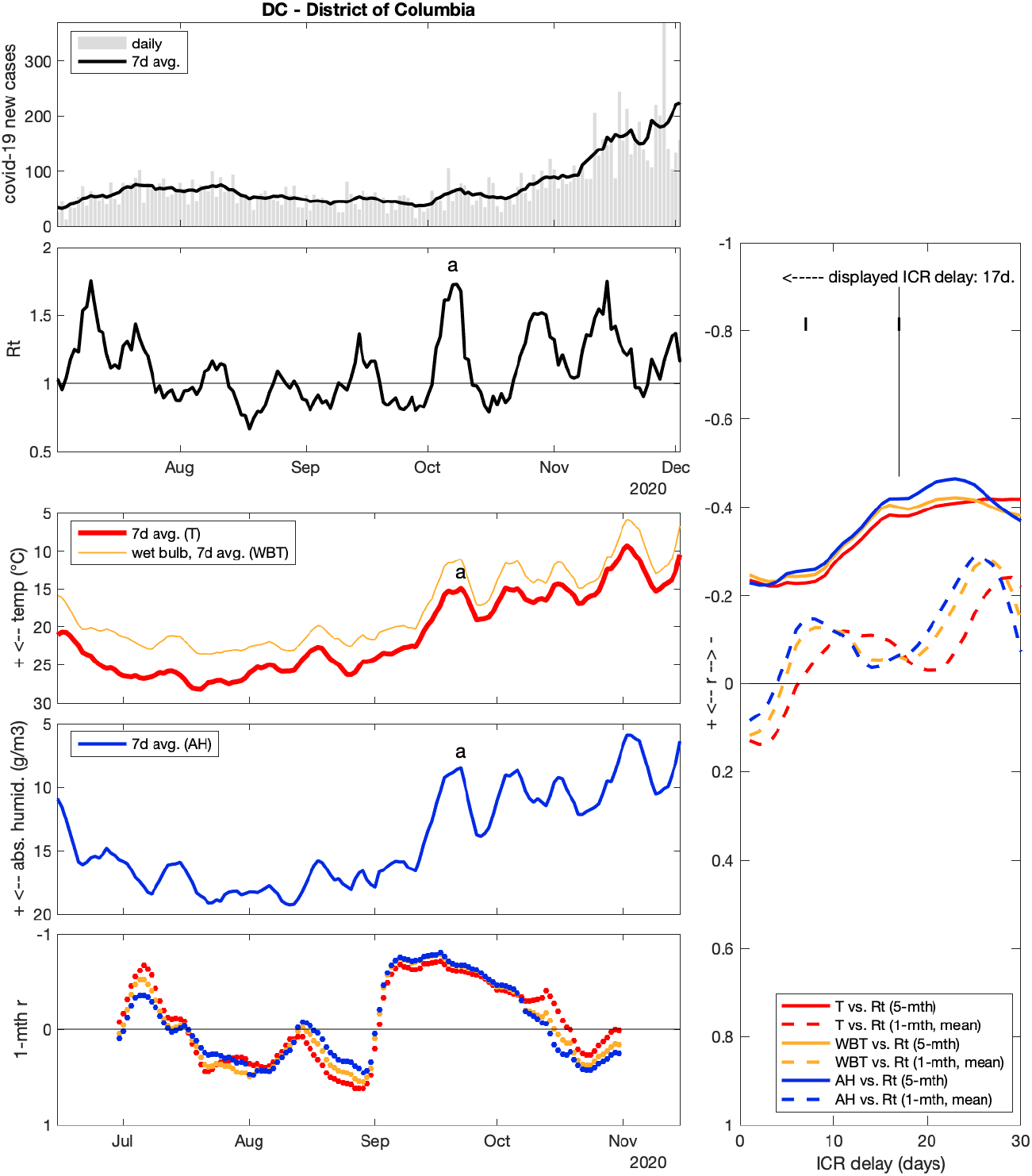

**Columbia district (DC)** r > −0.42

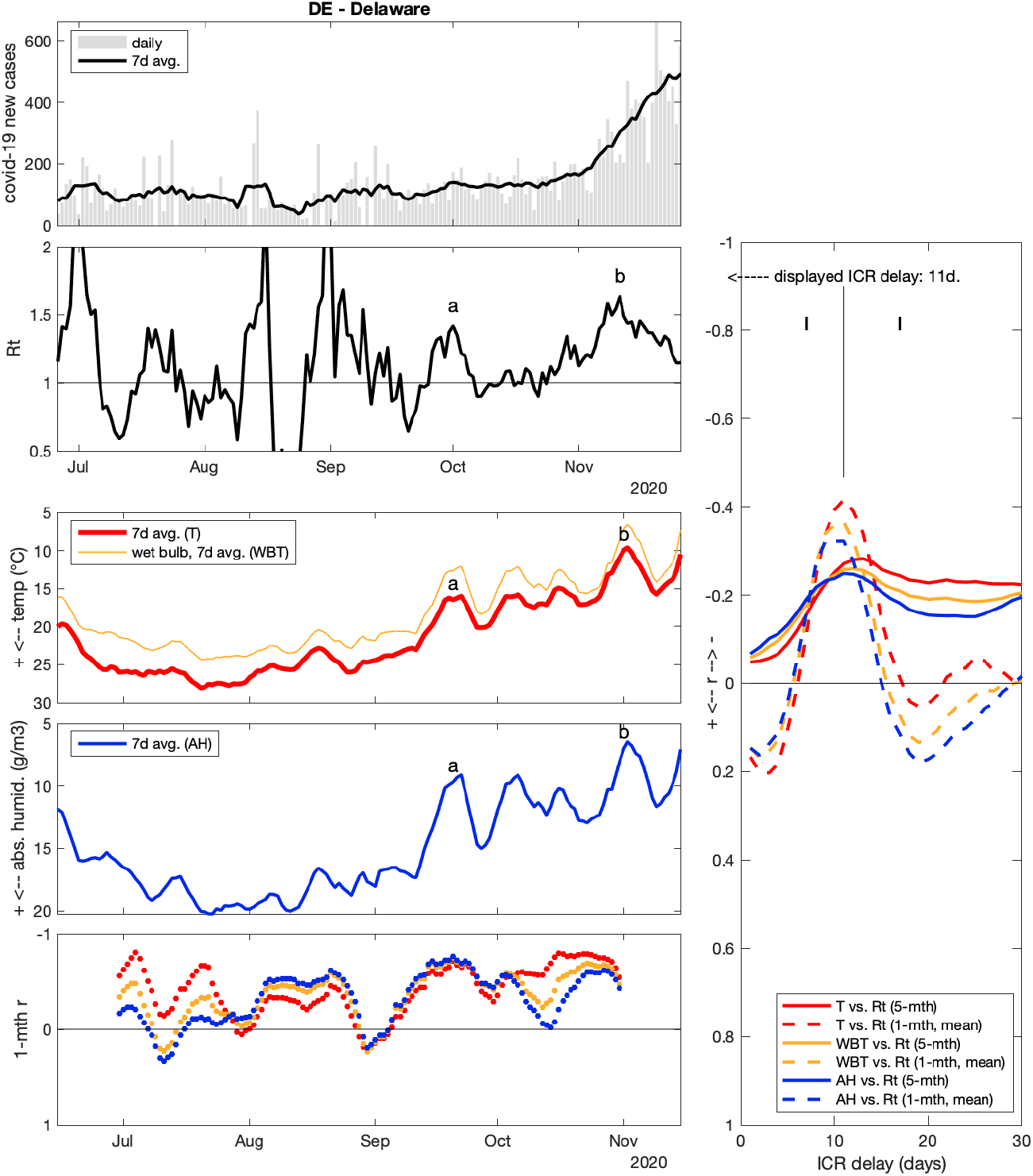

**Delaware (DE)** r > −0.42

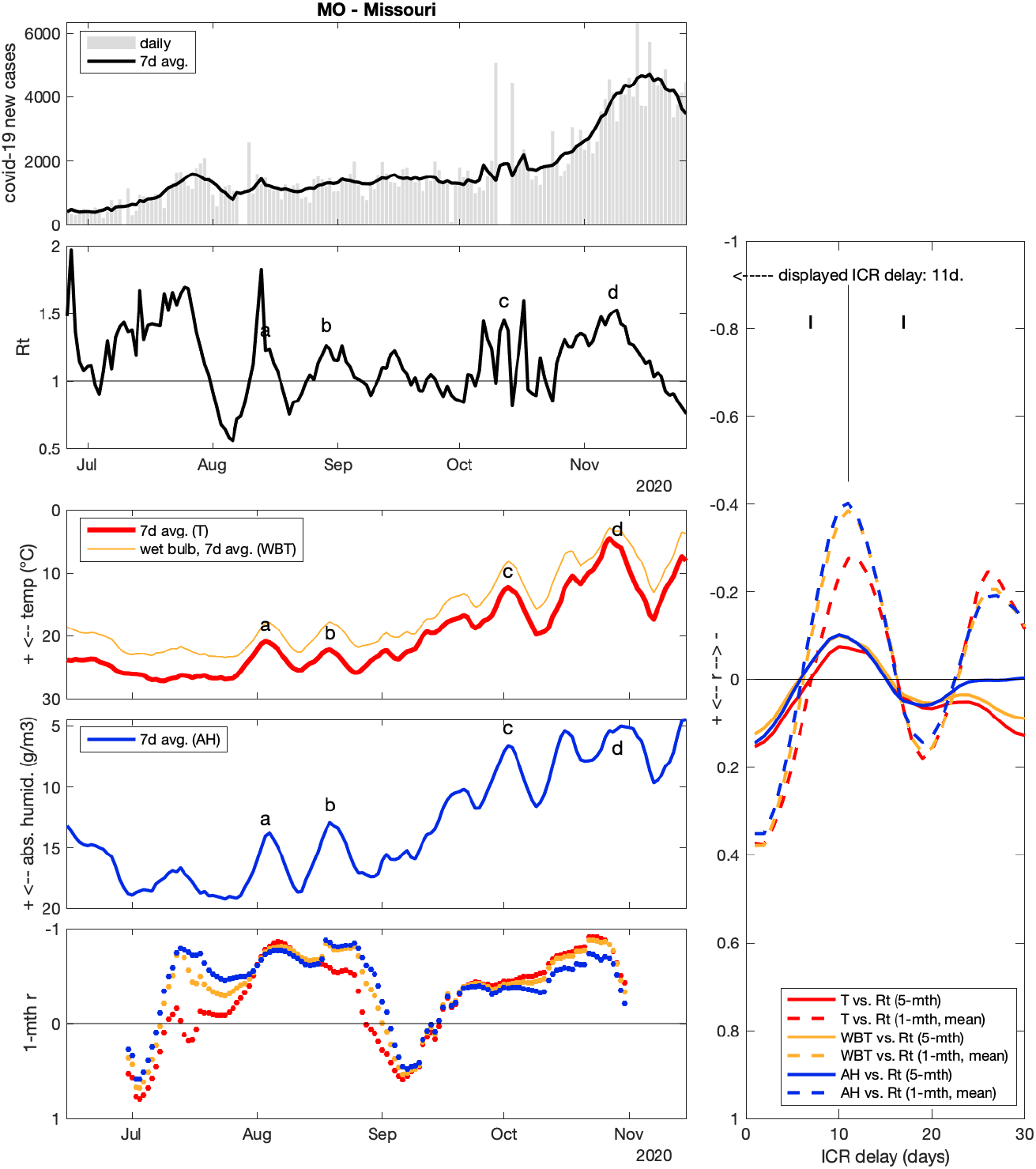

**Missouri (MO)** r > −0.41

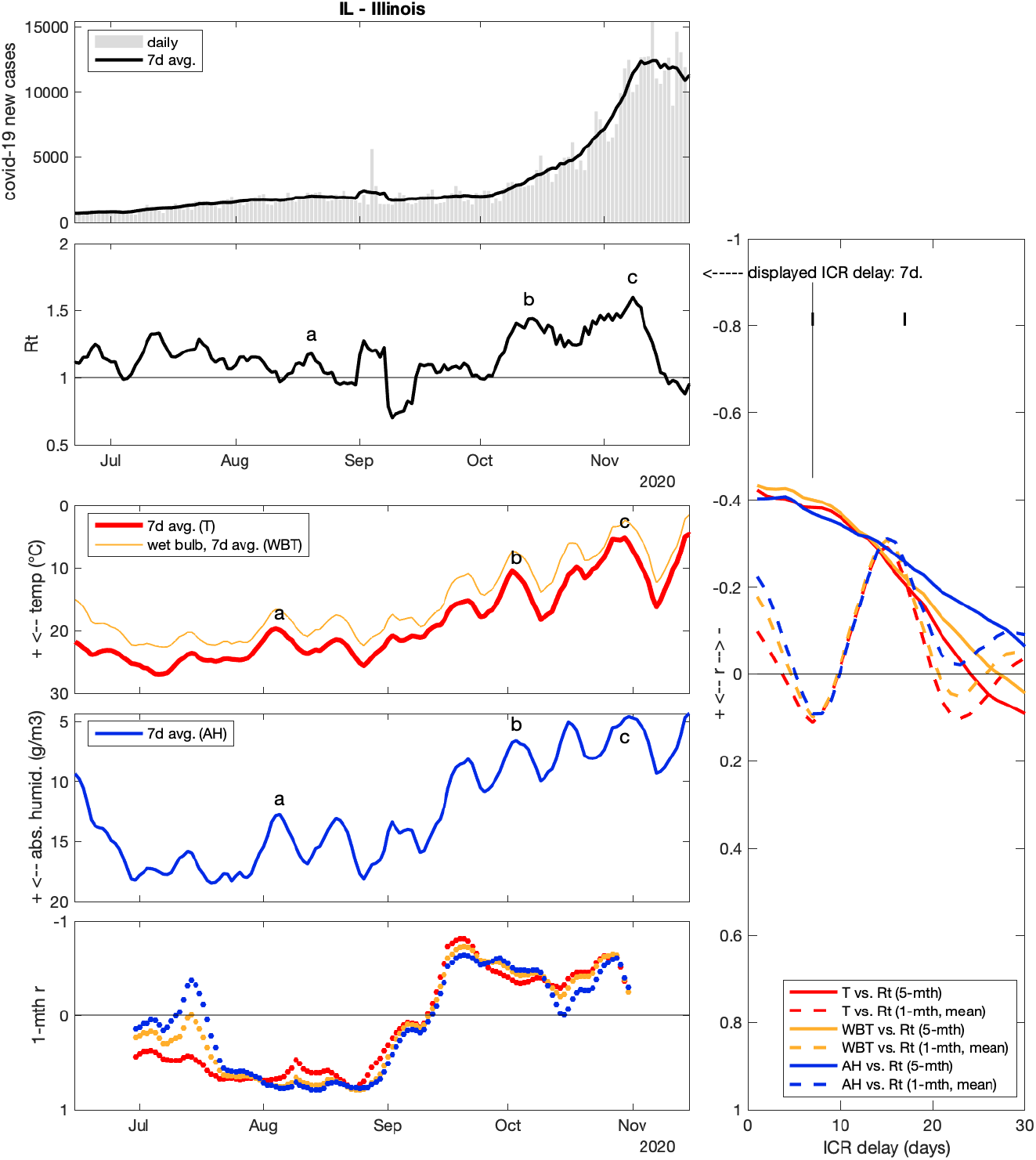

**Illinois (IL)** r > −0.41

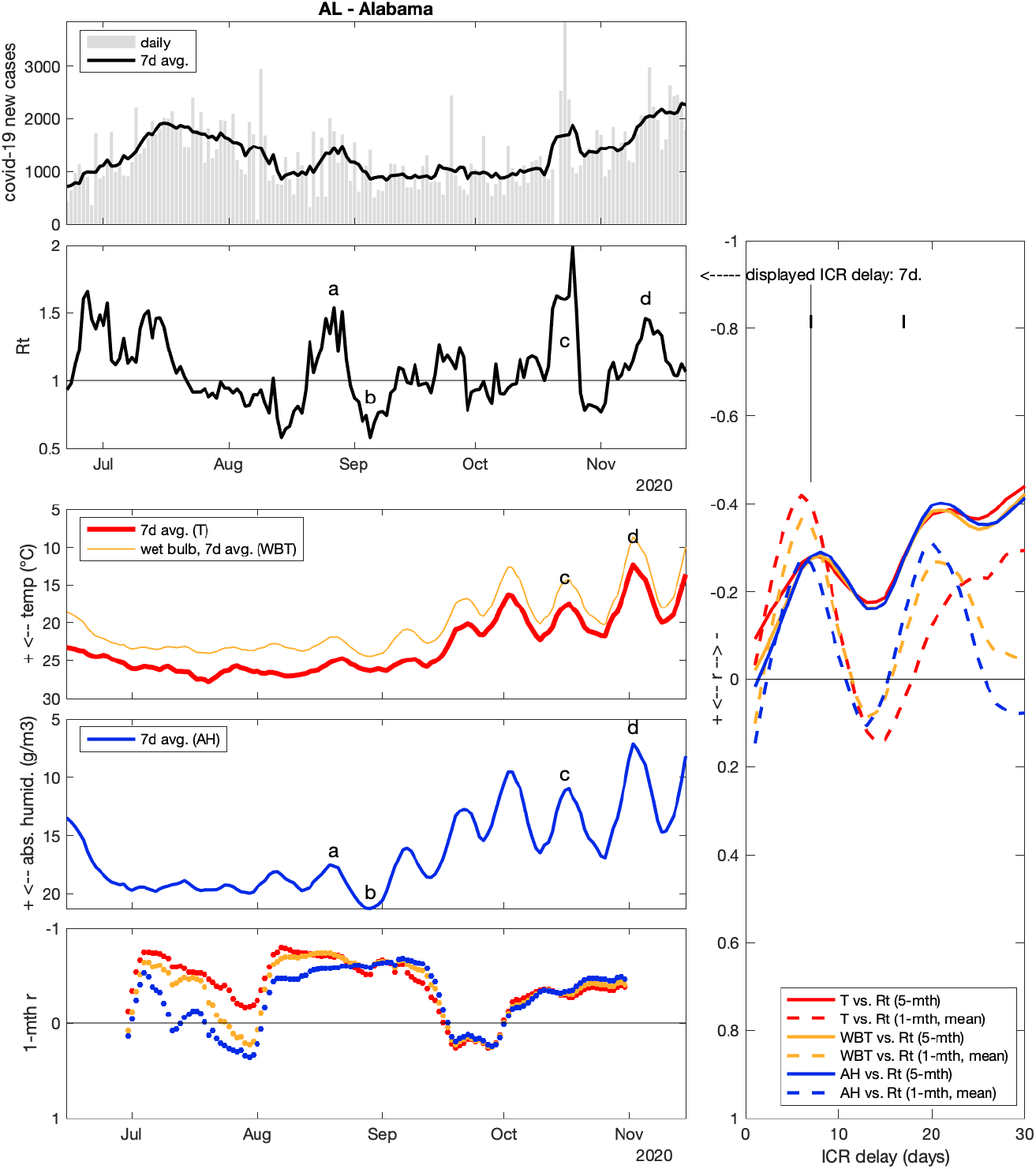

**Alabama (AL)** r > −0.4

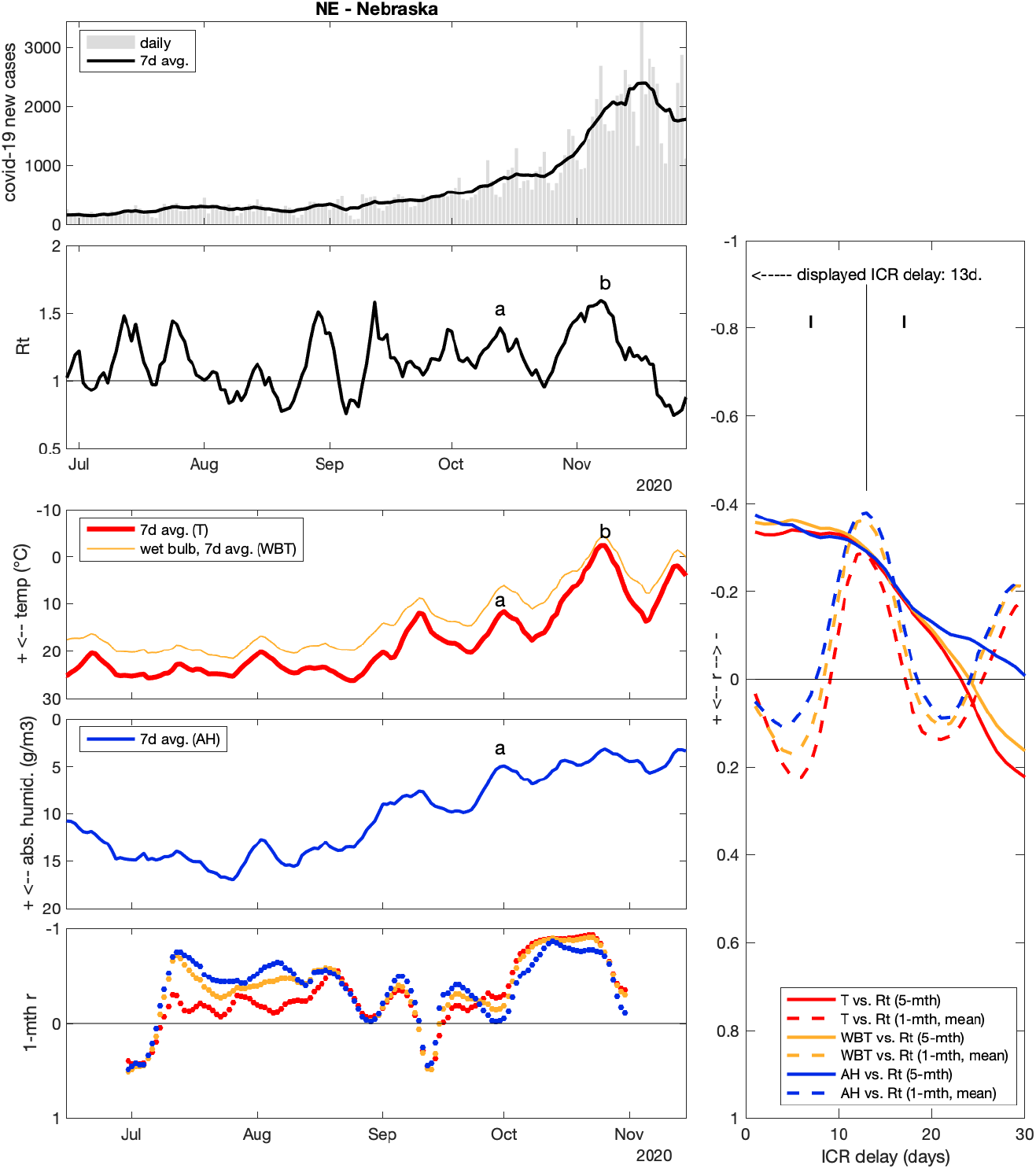

**Nebraska (NE)** r > −0.38

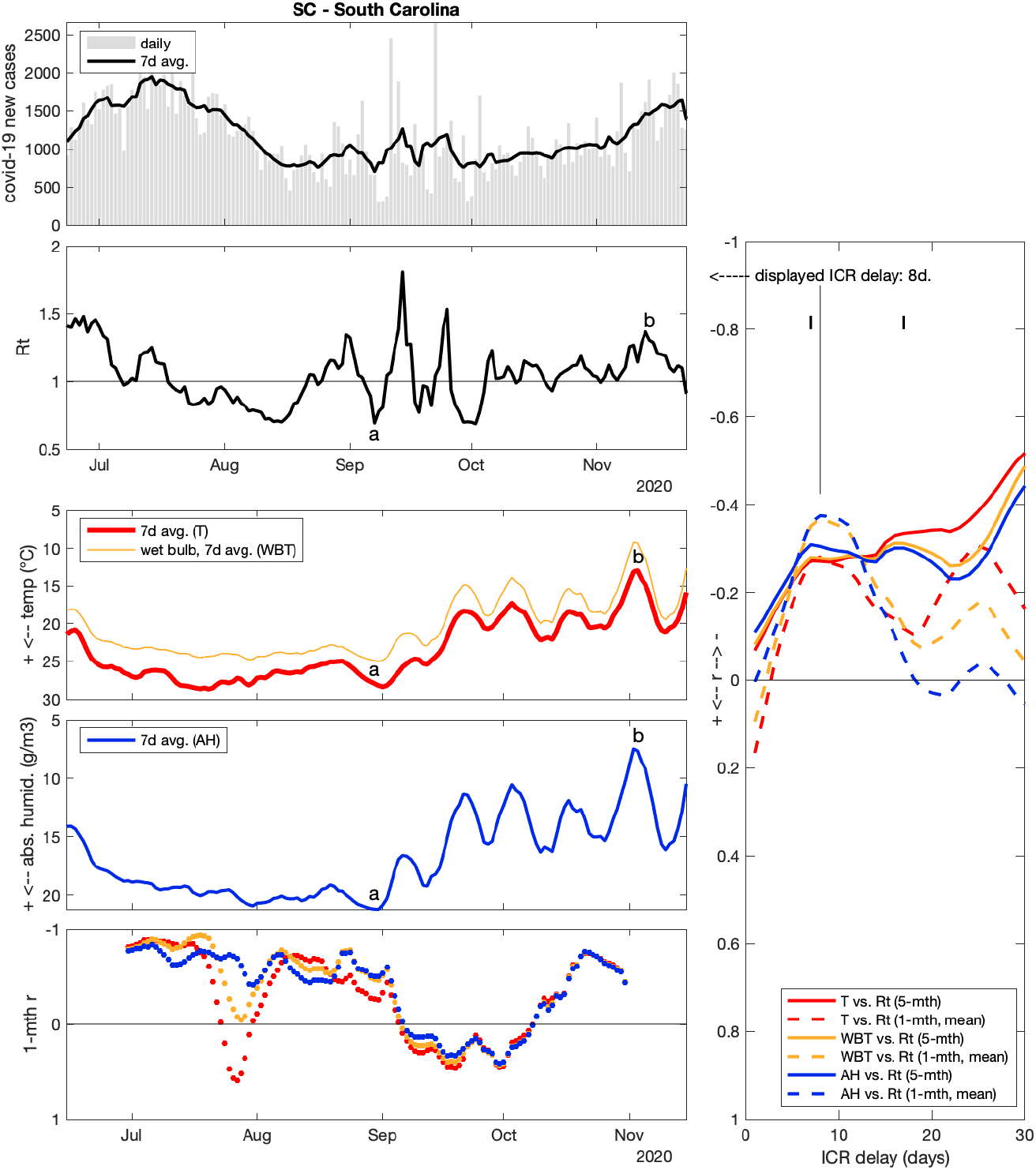

**South Carolina (SC)** r > −0.38

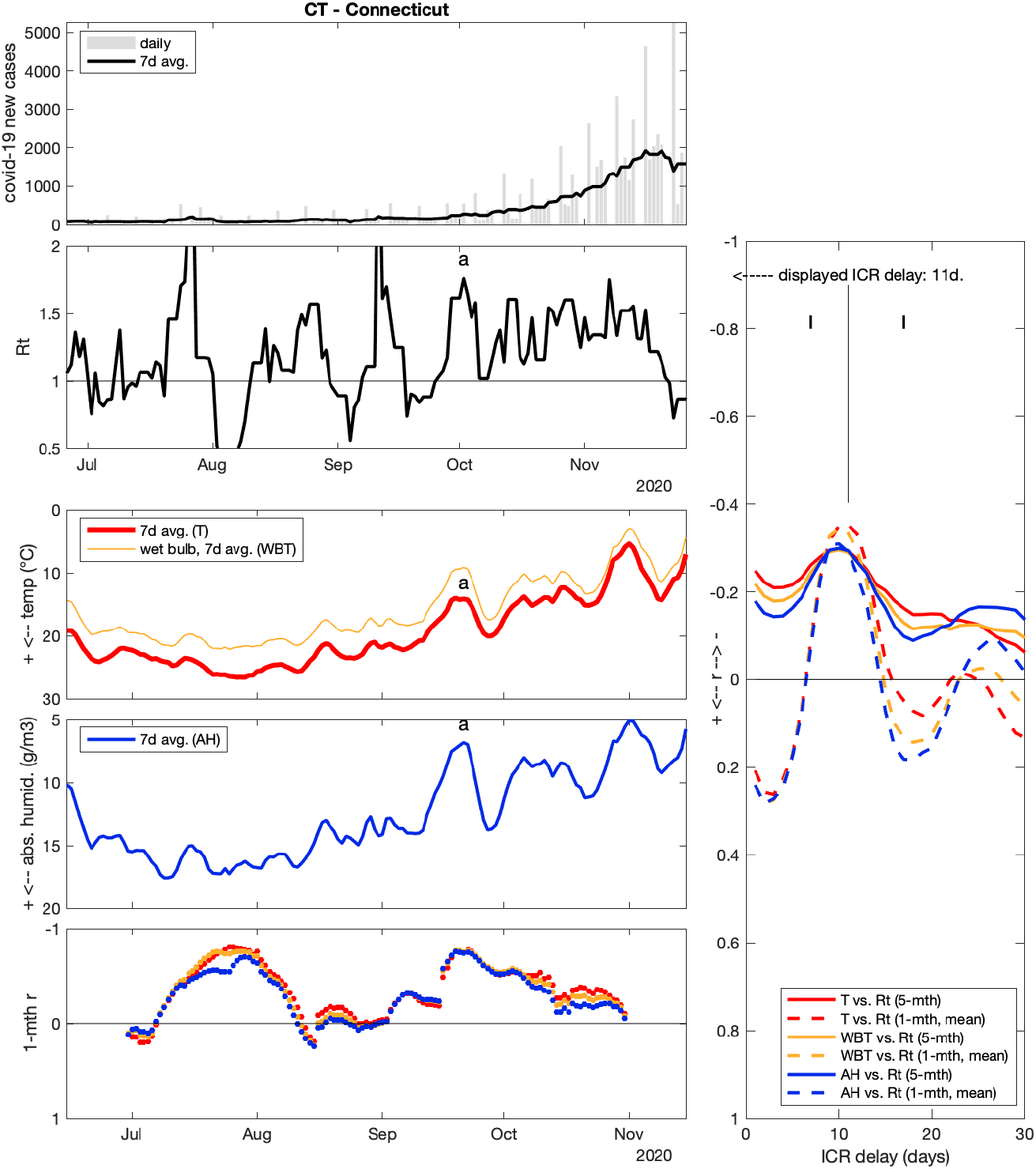

**Connecticut (CT)** r > −0.36

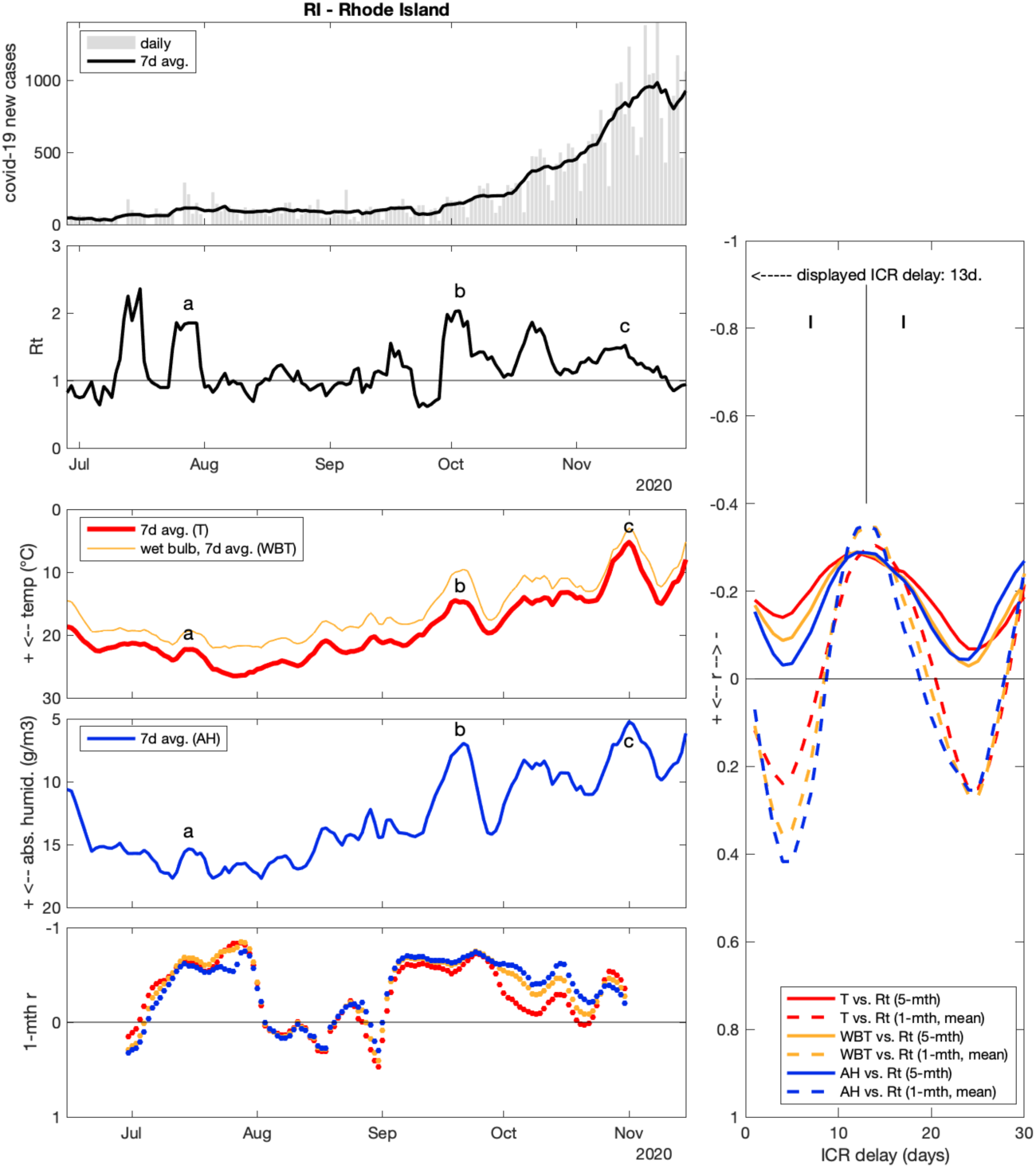

**Rhode Island (RI)** r > −0.36

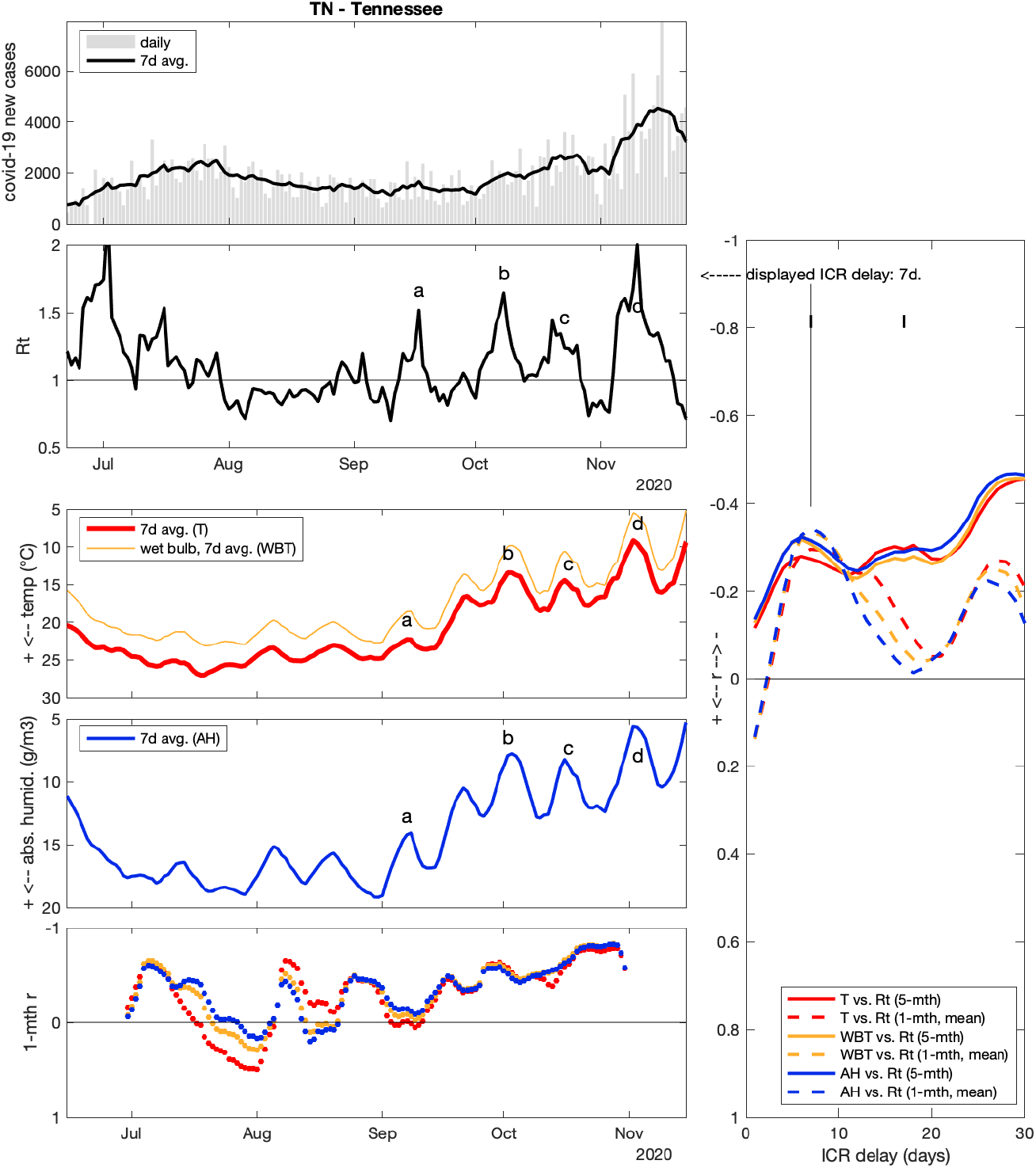

**Tennessee (TN)** r > −0.34

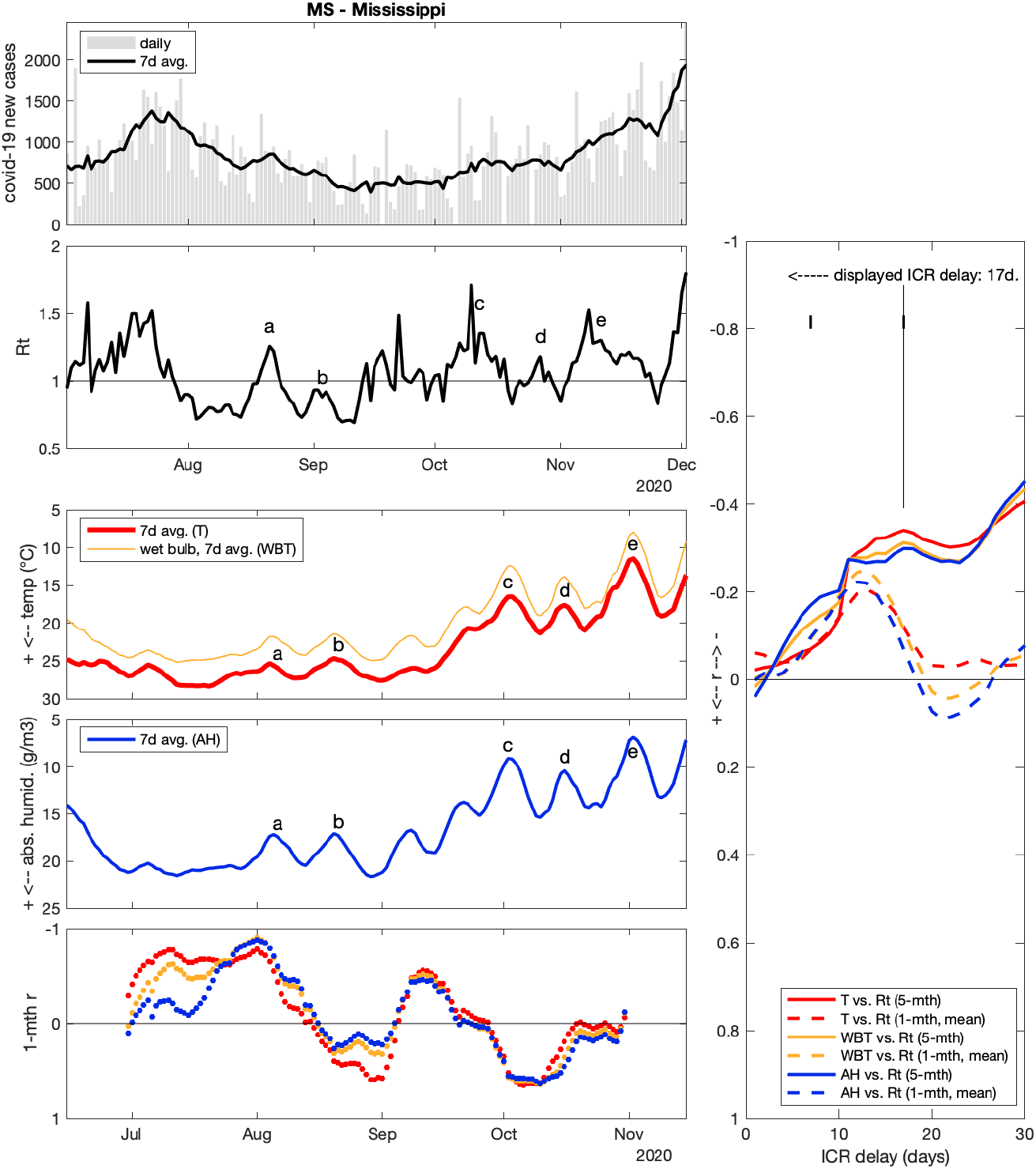

**Mississippi (MS)** r > −0.35

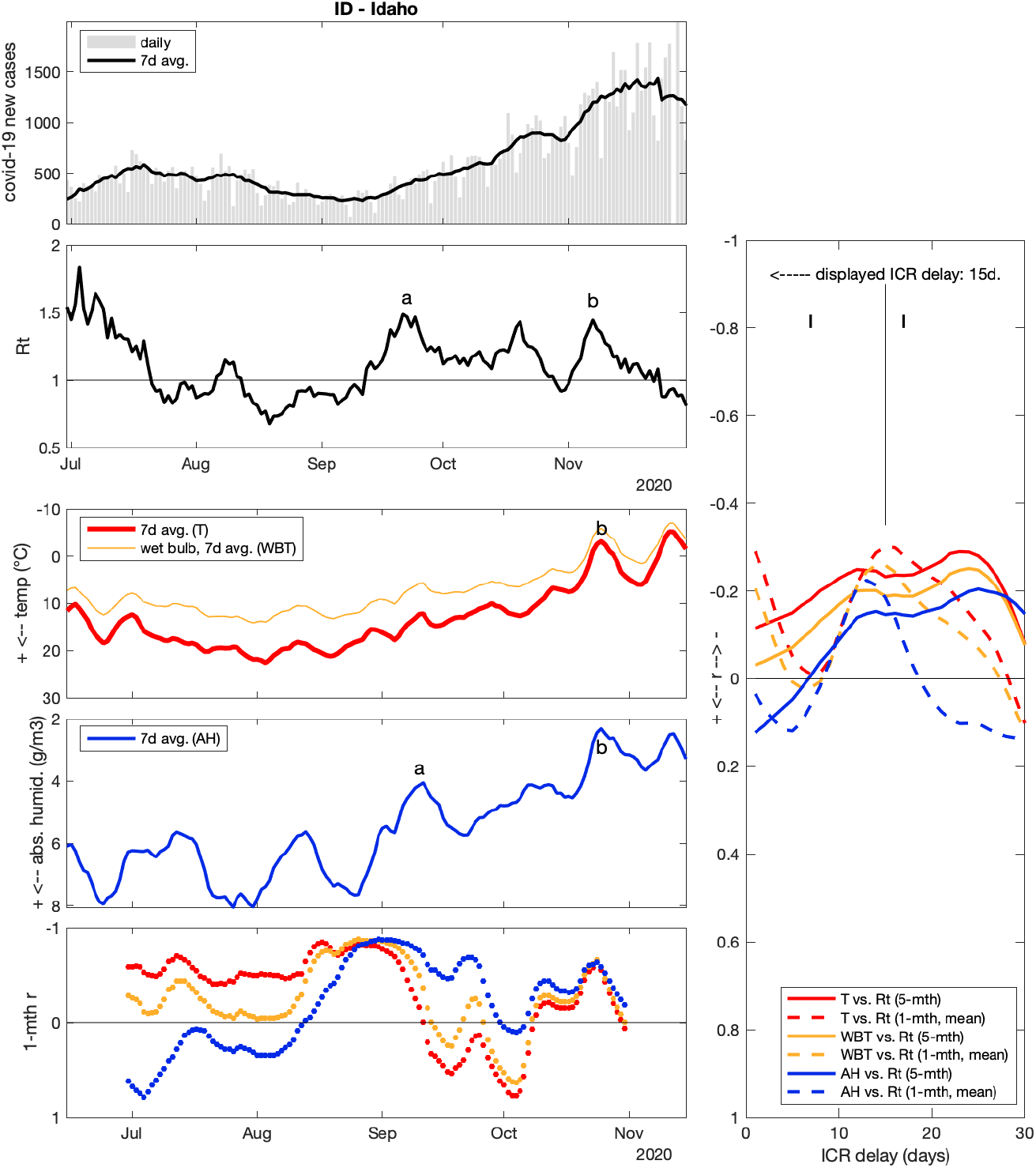

**Idaho (ID)** r > −0.31

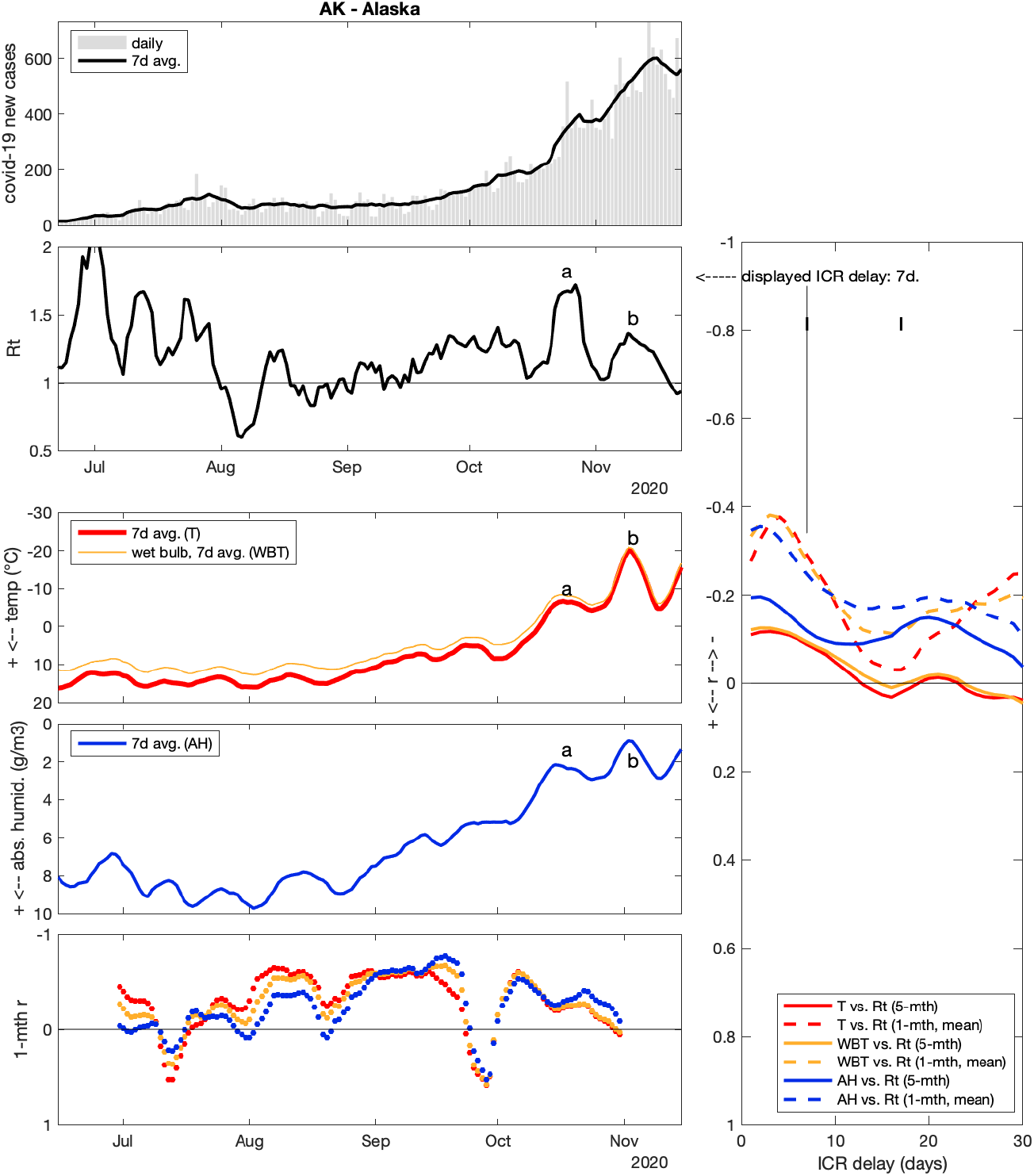

**Alaska (AK)** r > −0.30

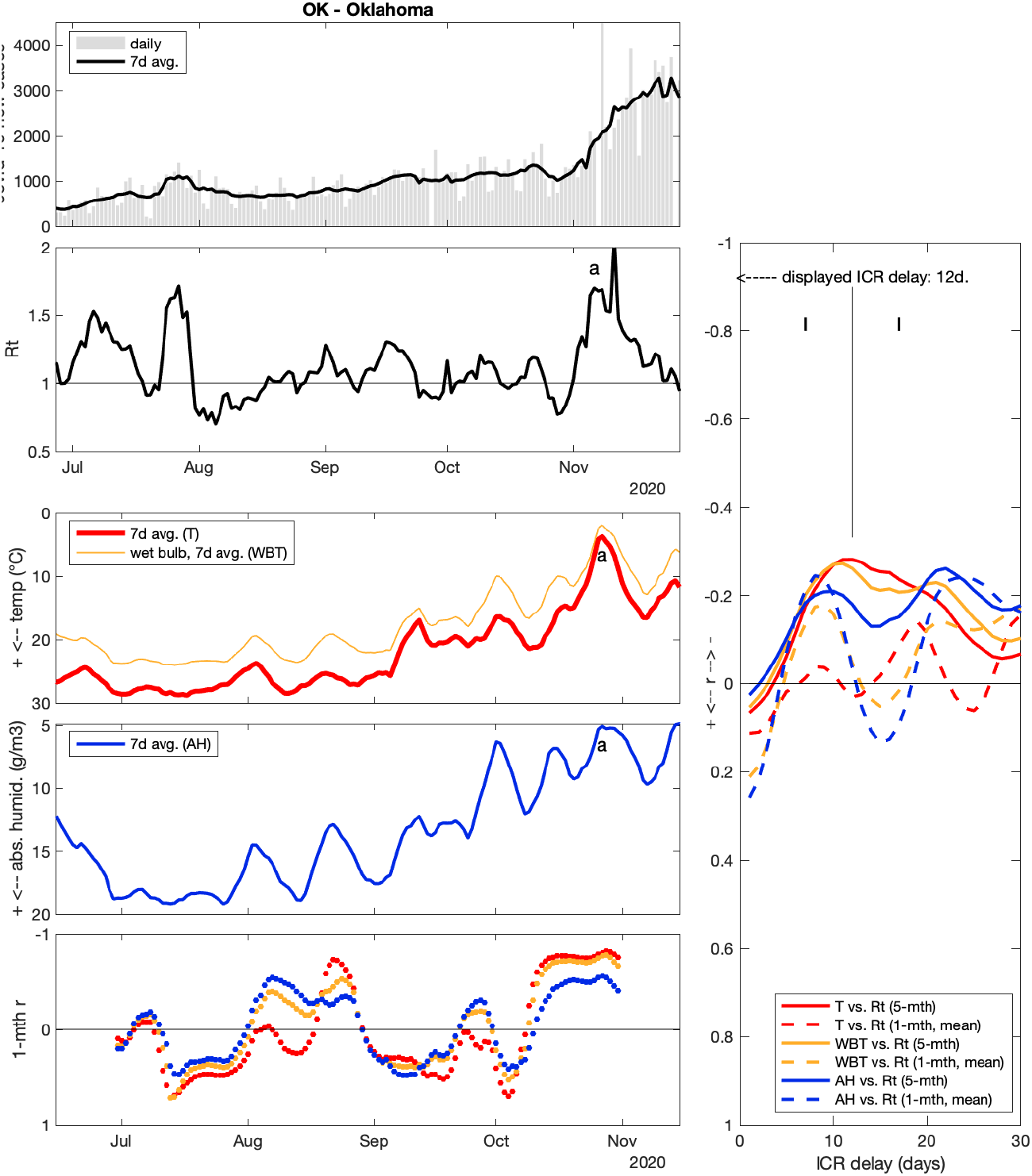

**Oklahoma (OK)** r > −0.28

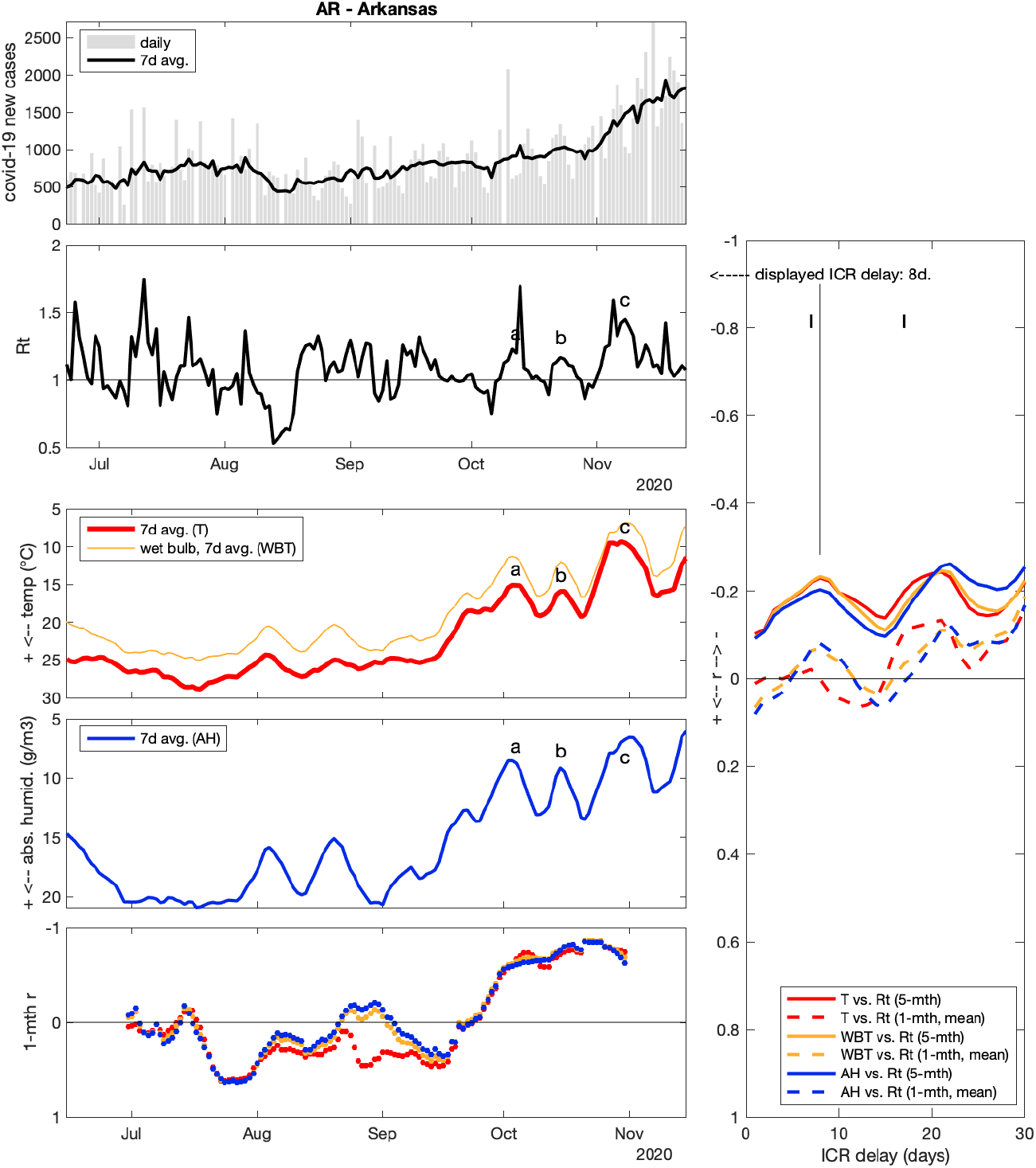

**Arkansas (AR)** r > −0.24

### 7 states with discrepant weather/Rt correlation

These remaining states have weak to no negative weather-Rt correlations (−0.2 **<** r **≤** 0), or show no negative r peak for realistic ICR delays, or even have positive weather-Rt correlation for some variables and/or delays.

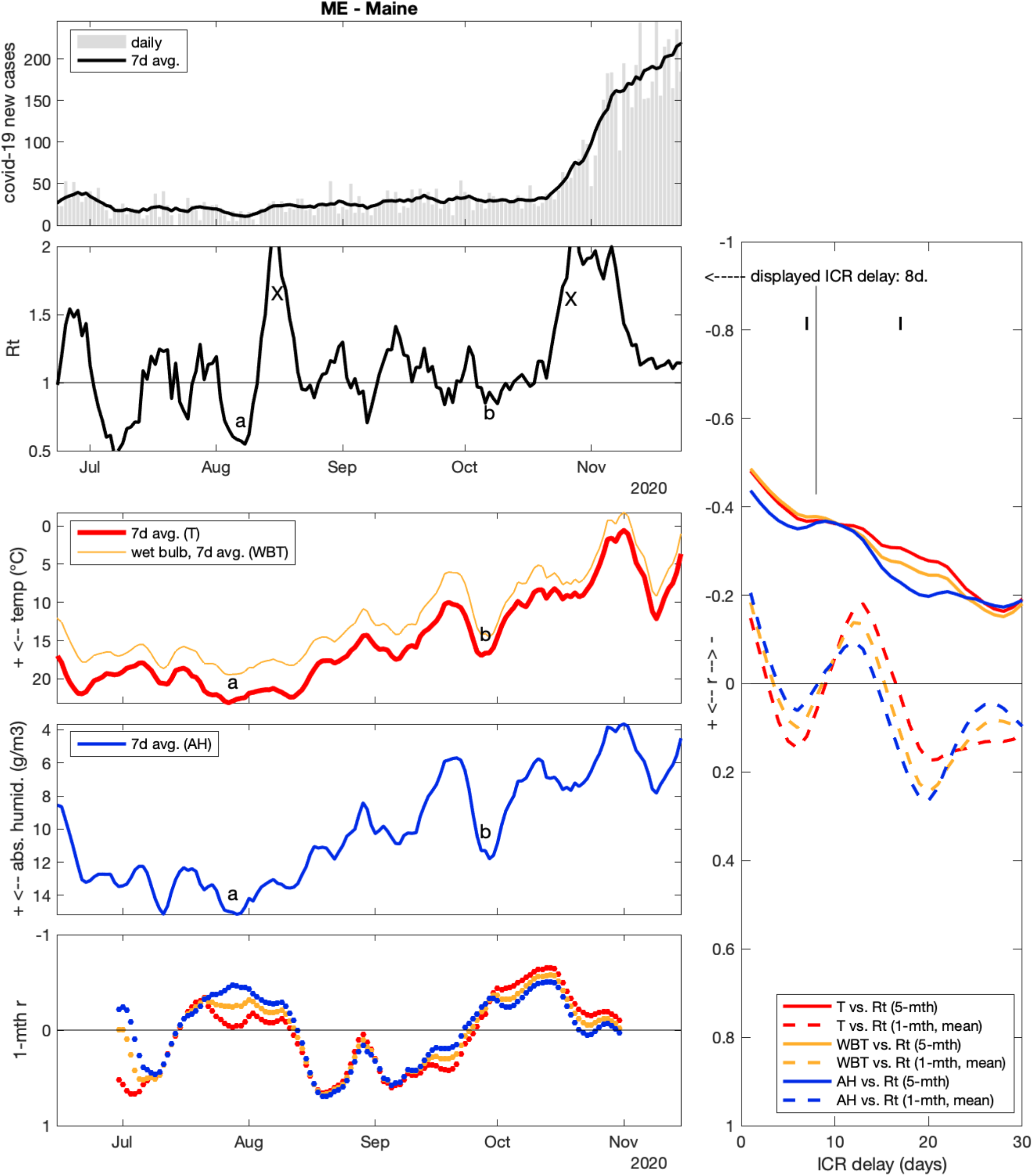

### Maine (ME)

in ME, 5-month correlation between weather and Rt was negative and moderate (r ∼ −0.4), reflecting a general trend towards higher Rt and colder, drier weather through the study period. However, 5-month r plots showed no clear peak around 12-day ICR. Moreover, mean 1-month r could be negative or positive (by similar amplitudes) depending on the assumed ICR. When looking at Rt and weather data, there are a few matching episodes, especially for low Rt values (a, b), but also strong discrepancies (X). Namely, large Rt peaks around Aug 15 and Oct 27 could not be related to any cold-dry episode within realistic IRC delay. Note that when new case counts are very low (e.g. <100, as is the case for ME before November), case data is potentially more sensible to new large covid-19 clusters, making Rt variations less deterministic and more stochastic.

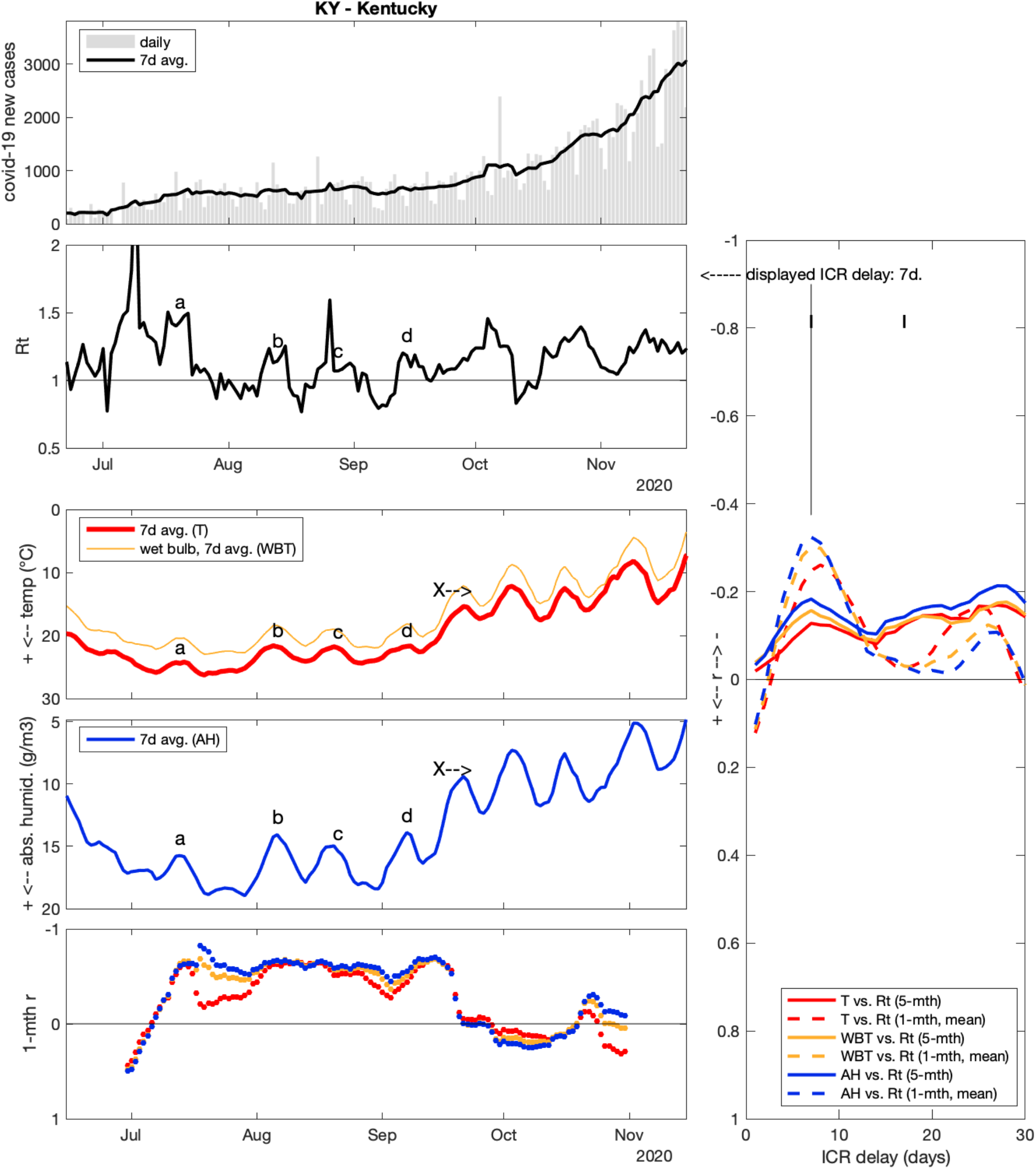

### Kentucky (KY)

5-month correlation between weather and Rt was weak in KY, with −0.2 < r < 0 for any realistic ICR. On the other hand, 1-month r showed a moderate peak for ICR = 7-8 days, and some peak matches (a-d) could be observed until mid-September. However, a clear weather change towards colder and drier air around Sep 16 (X) could not clearly be related to any strong Rt change in KY, and the 1-month r drops to ∼zero in the autumn (lower left panel).

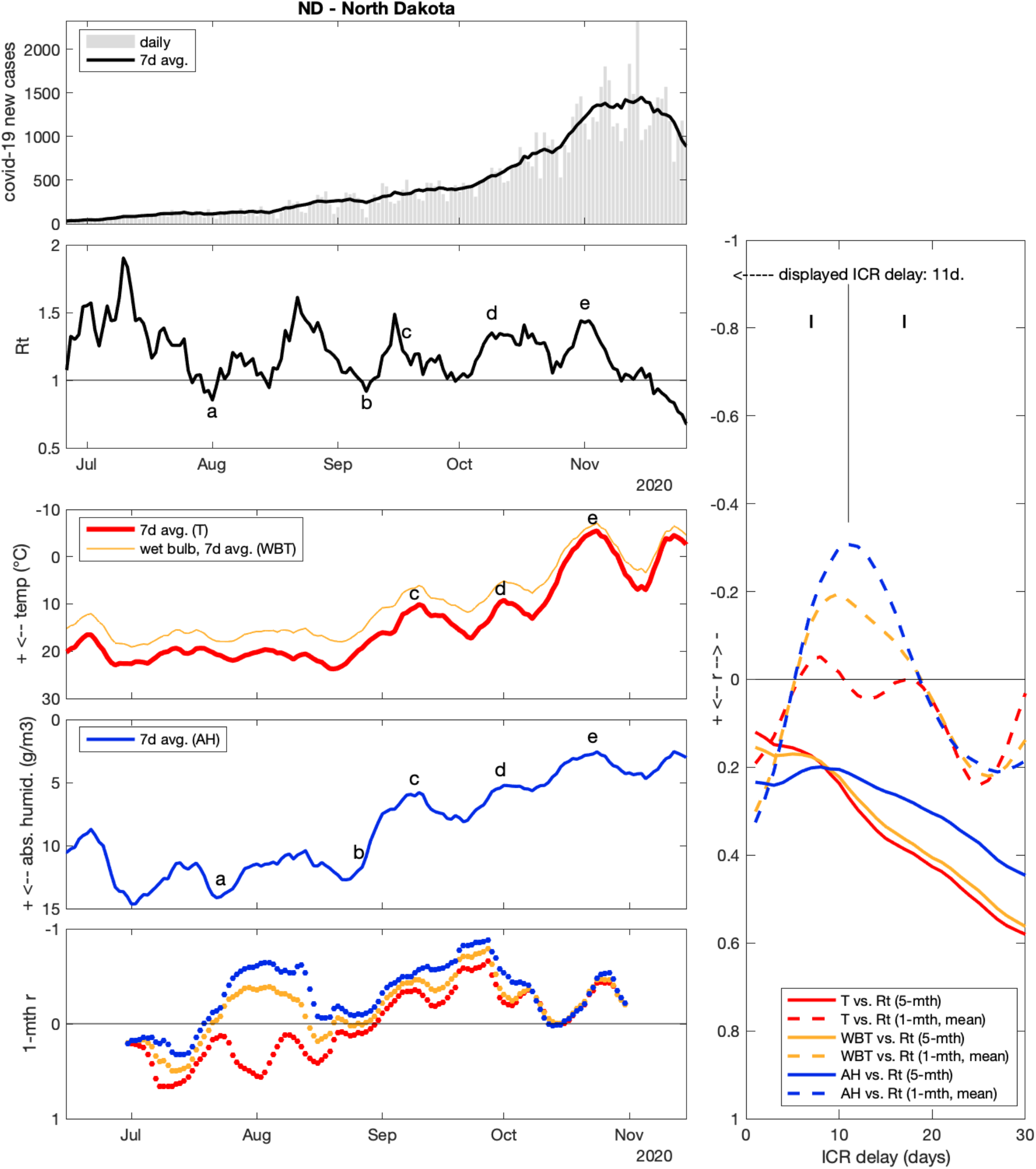

### North Dakota (ND)

Surprisingly, correlation between weather and Rt over the full study period was positive in ND (r ∼ 0.15 to 0.4 for realistic ICR delays). Indeed, while T and AH decreased from June to November (inverted axes), Rt also showed an overall tendency to decrease, contrary to most other states (note that as long as Rt remains > 1, case counts still increase through time). On the other hand, mean 1-month r show a more typical negative peak, especially for AH (r = − 0.31 for ICR = 11 days), and one can associate Rt peaks with weather episodes (a-e). In other words, Rt did increase after short-term cold-dry anomalies, but in ND this effect apparently came “on top” of an overall Rt decreasing tendency. Some elements might explain the Rt overall decrease: (1) very low case count (<100) early in the period might favor high Rt if large clusters occur, and (2) a possible onset of efficient social restrictions sooner in November than in other states.

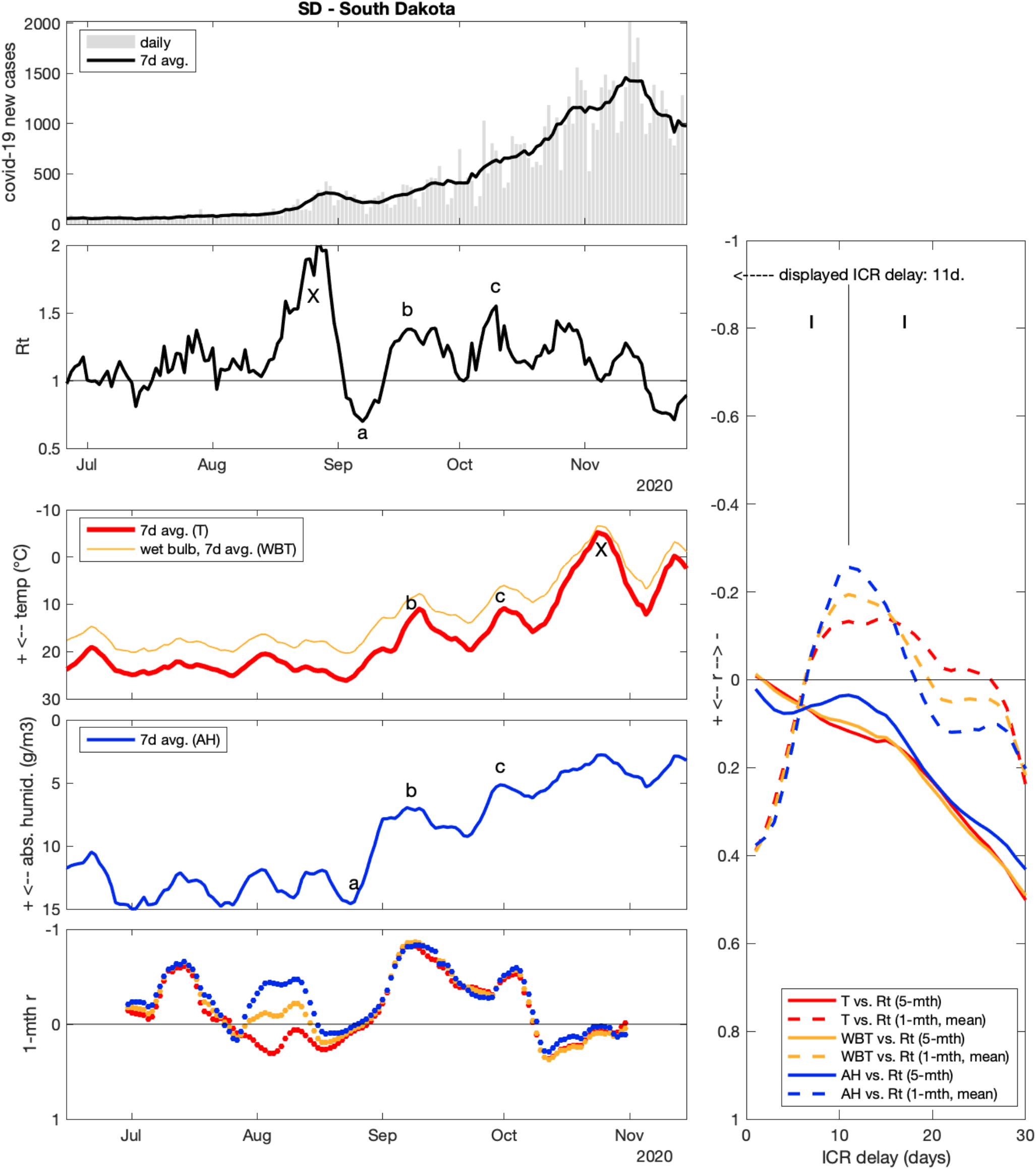

### South Dakota (SD)

Correlation results in SD were very similar to ND, i.e. overall positive correlation between Rt and weather variables, but negative short-term correlation (r = −0.26 for AH vs. Rt, for ICR = 11 days). Letters (a-c) on the figure show local similarities between T, AH and Rt evolution, but there were also notable discrepancies in SD: a late-August surge (Rt = 2) was not preceded by any strong weather change, and a very cold episode in late October (smoothed daily T = - 5°C) was not followed by a strong Rt increase within realistic ICR delay.

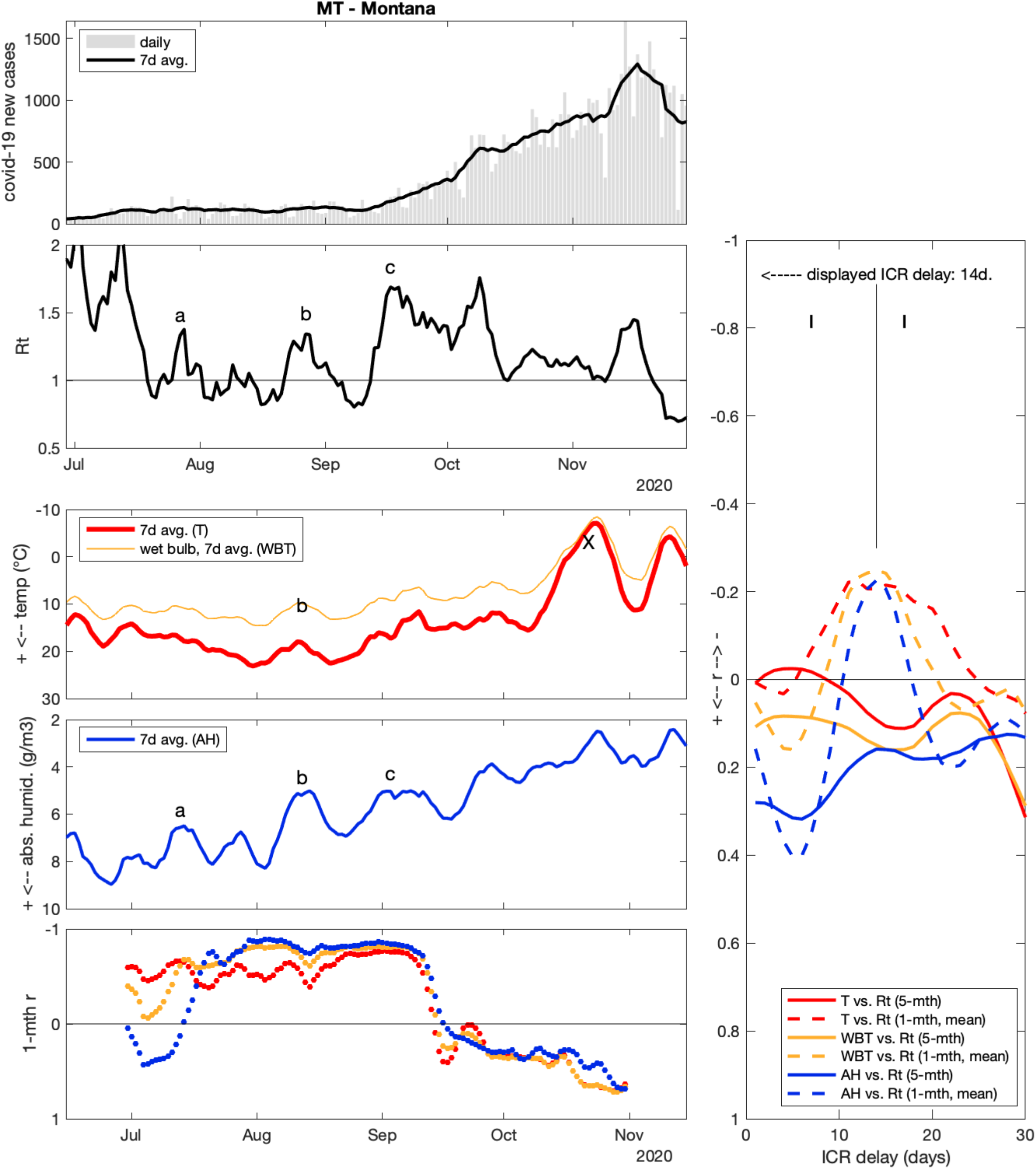

### Montana (MT)

As in ND and SD, 5-month r was positive in MT, but 1-month r showed a small negative peak (r = −0.25 for WBT, ICR = 14 days). In MT, short-term correlations were strongly negative until September, with several matching peaks (a, b, c), but later disappeared or turned positive (see lower-left panel), with a notable discrepancy (X) as a very cold episode (smoothed daily T = - 7°C on Oct 23) was not followed by a strong Rt increase within realistic ICR delay.

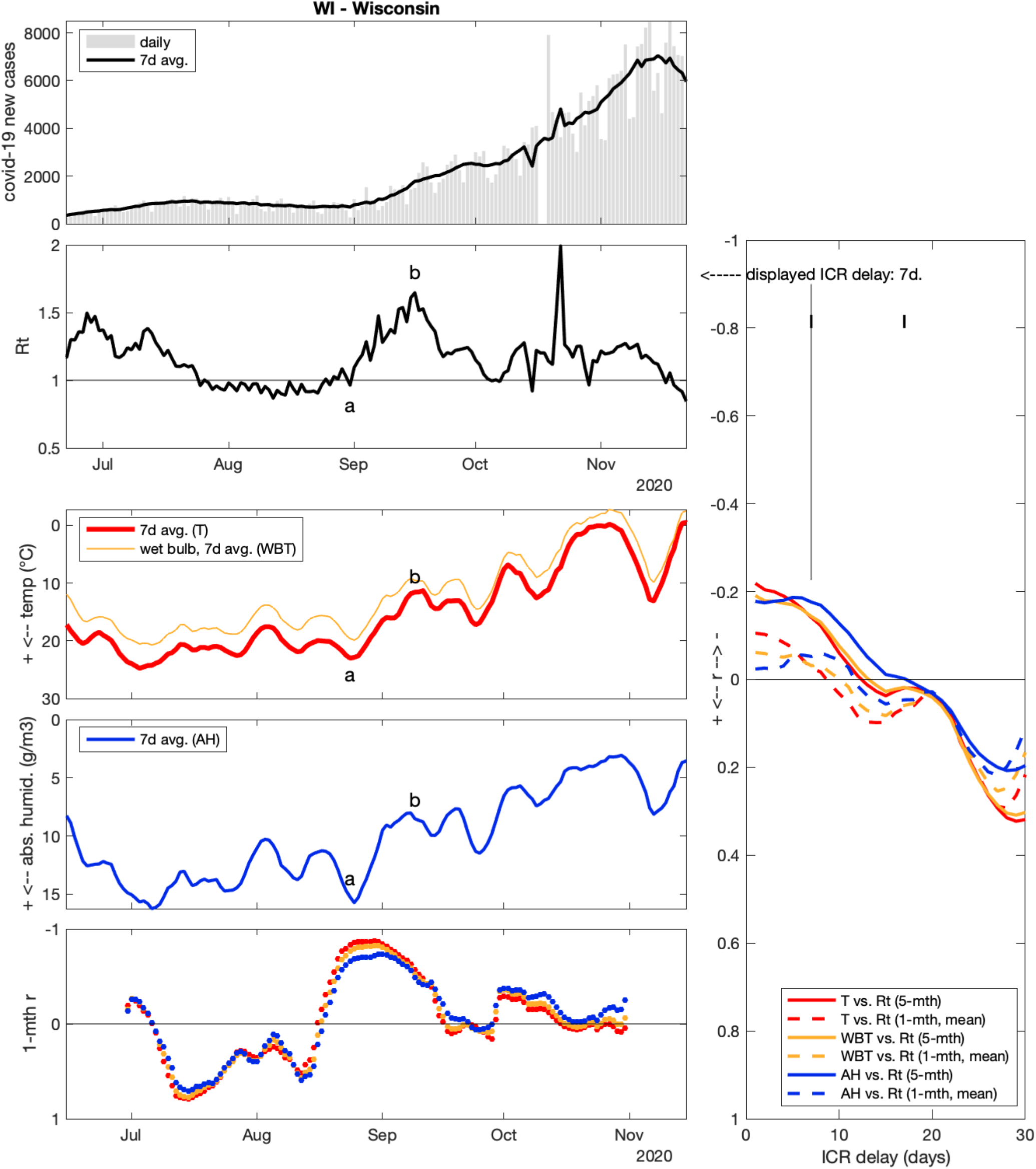

### Wisconsin (WI)

In WI, the September surge (Rt evolving from 1.0 on Aug 31 to 1.6 on Sep 16, a to b on figure) was preceded by a cold-dry evolution about one week earlier (T decreasing by 12° and AH by 8 g/m^3^ from Aug 25 to Sep 8). However, other episodic associations over the study period are lacking. As a result, 5-month r remained close to zero (−0.2 < r < 0), as did mean 1-month r (−0.1 < r < 0.1). WI data as a whole did not support the moderate to strong negative correlations observed for many other states.

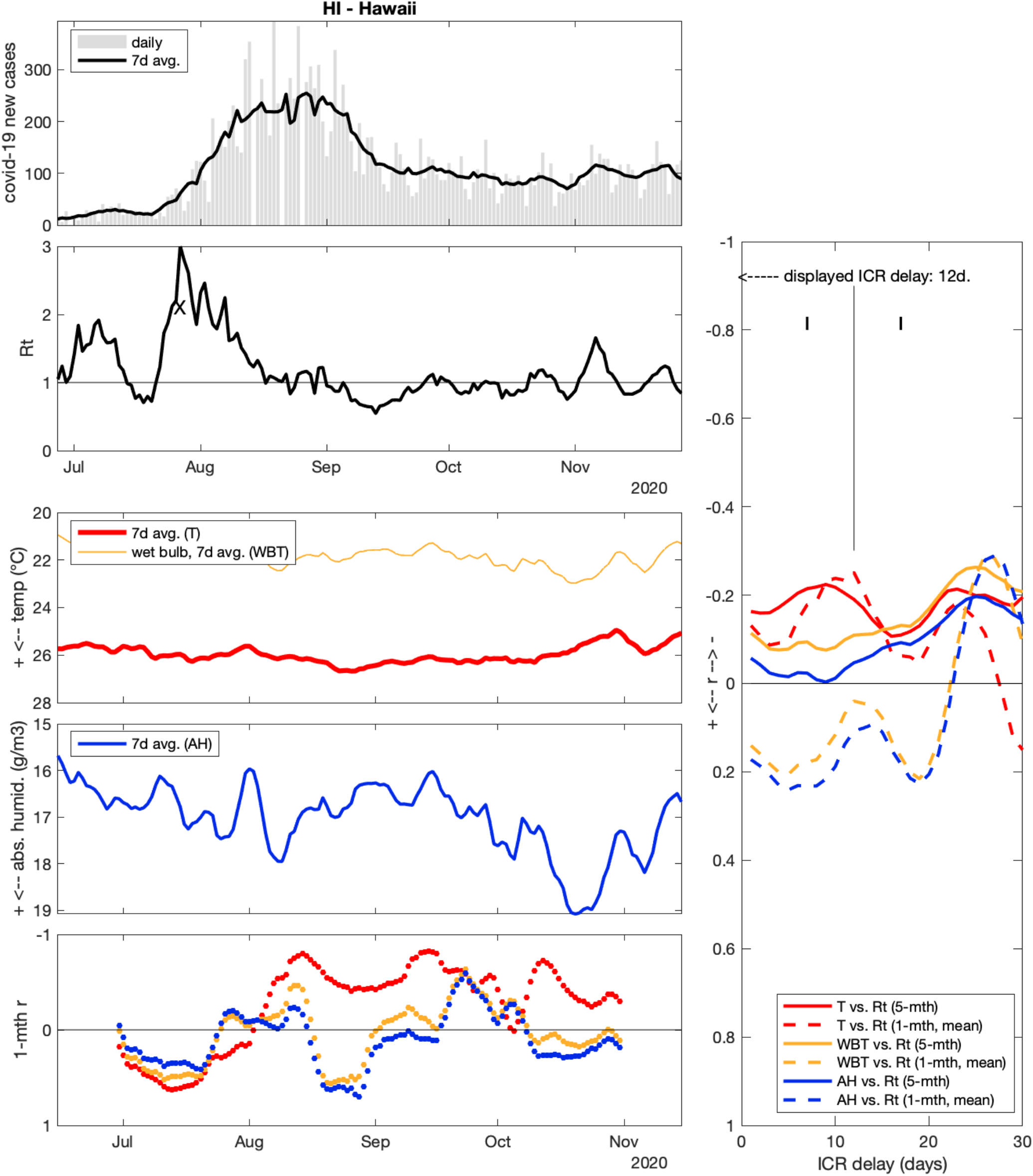

### Hawaii (HI)

In HI, T and AH were not surprisingly much more stable over the study period than in most other states. Still, there was a summer surge (X) with daily new cases increasing from ∼20 to ∼250, Rt attaining 3 on Jul 27. This surge could not be related to T or AH variations.

## Discussion

Associating short weather episodes with local Rt peaks in individual states is interesting, but certainly has its limits. The main result of the present study is rather based on the convergence of these local correlations, as initially presented in Fig. 1 (p.6): (1) Rt was most often negatively correlated to weather variables such as T, WBT and AH and (2) this correlation was stronger for time shifts of around 11 days, i.e. delays that are congruent with estimated time between infection and case report (ICR, see methods).

A spurious correlation between weather and Rt could be due to weather becoming naturally colder through autumn, and Rt coincidentally increasing, as U.S. approached a 3rd wave nationally. 5-month correlation remaining moderately negative for all tested ICR, from 1 to 30 days (Fig. 1, top) is the expression of such overall, mildly informative covariation between weather and Rt. However, the peak of 5-month correlation around 10-11 days of delay cannot be explained by such a confounding covariation. Even more striking is the result that mean 1-month r becomes moderately negative *only* for ICR around 11 days (Fig. 1, bottom).

Another spurious effect could be that Rt and weather coincidentally oscillate with similar time periods, and the peak in correlation for a given state would only express the phase shift between unrelated periodic signals. Under this hypothesis, the phase-shift value should be essentially random, and different from state to state. This not what was found here, as pooled national data shows a convergence of phase shifts across states (Fig. 1).

The detailed observation of weather vs. Rt in a given state sometimes shows striking parallelism between weather record and time-shifted Rt data (see e.g. New York state on p.8), but the parallelism is never perfect, and sometimes hardly visible. This is not surprising as (1) covid-19 and weather variables were collected from unrelated sources with different methods, and hence are expected to contain different noise/error patterns. Moreover, (2) even if there is a causal mechanism linking weather to covid-19 Rt, weather is not expected to have a direct, linear effect on transmission (see below). Last, (3) even though the study period was selected for comparatively low social restriction changes, it is probable that restrictions varied to some extent, and also that people’s behaviour adapted to the report of local surges, which could have a self-leveling effect on observed Rt.

### Hypothetical causal mechanisms

There is an apparent paradox in finding correlations between Rt and outdoor weather data, while it is now commonly admitted that a majority of covid-19 transmissions are airborne, and occur indoors [1]. Here I list 3 putative mechanisms (via T, WBT and AH, respectively) that might link outdoor weather with indoor disease transmission. These proposed mechanisms are admittedly speculative at this point, and not exclusive: 2 or 3 of them might be at play simultaneously, or alternatively according to the time period and temperature range. Maybe only one of these mechanisms operates, but correlations are observed for all three weather variables because they are physically related (see equations in Methods section). It is also possible that none of the following causal hypotheses hold, and that other mechanisms are at play.

i. Outdoor temperature (T) might have at least two effects on human behaviour that can play an important role in indoor airborne transmission. First, as weather becomes cold (e.g. progressively through autumn, or on an episodic “cold wave”), a significant proportion of population might actively avoid long periods of time outdoors, and thus spend a larger proportion of their time indoors. Collectively, this can concentrate viruses in indoor air, and also individually increases the probability to encounter contaminated air. Moreover, while staying indoors, people might be less prone to ventilate their home or office room frequently if the outdoor temperature is very cold, for thermal comfort reasons. Both of these possible behavioral effects are not expected to have a linear relationship with T: for example, avoidance of outdoor space and indoor ventilation appears likely when outdoor T drops from 10 to 0°C, but less so from 30 to 20°C. However, by looking back at state results, one can observe Rt peaks associated with colder episodes for a wide range of outdoor temperatures, from below 0°C (e.g. MN, ND, ID) to above 20°C (e.g. FL, VA, KY, MO).
ii. Outdoor wet-bulb temperature (WBT) is a compound variable integrating both T and RH [16], and can be understood as a proxy to upper respiratory tract temperature of people breathing outside (see Methods). A low tract temperature might in turn facilitate penetration of the virus through several cellular effects (e.g. [14,15], and other mechanisms cited in [2]). As such, outdoor WBT might be of importance to (rare) outdoor disease transmission, e.g. in crowded outdoor spaces (e.g. streets, queues, etc.). On the other hand, an effect of low outdoor WBT on indoor transmission is not straightforward if indoor spaces are heated for comfort. Still, an effect might exist due to the mobility from indoor to outdoor environment. If, after “catching viruses” in a contaminated room, people go outside in cold, dry air (e.g. for daily commuting), the viruses deposited in their upper respiratory tracts might beneficiate from a cooling mucosa, and see facilitated conditions for infection. Similarly, before entering a building filled with contaminated indoor air, if a person breathed cold, dry air outside, it is possible that her/his respiratory mucosa remains cold and “fragile” to virus penetration for a certain amount of time, before warming up.
iii. A less speculative and more general mechanism implies absolute humidity (AH). Note that like WBT, AH is calculated from T and RH, and as such is a compound of 2 primarily collected variables. However, unlike WBT, AH ultimately reflects a basic physical property of air: simply the amount of water per air volume, in g/m^3^.

In first approximation, indoor air initially comes from the outdoor air reservoir. During the cold season, indoor air is often heated for comfort, hence indoor and outdoor T become very different. This is not the case for AH: in the absence of secondary indoor water vapor sources, AH is conserved from outside to inside [20]. Thus, AH is the only outdoor variable of the 3 analyzed here that is conserved indoors.

Indoor heating is known to “dry air”, but this refers to RH, not AH. For example, outdoor air at 10°C and 80% RH contains about 7.5 g of water per m^3^ (the maximum it could contain, i.e. at 100% RH, would be 9.4 g/m^3^). If this outdoor air is simply drawn indoors and heated to 20°C (thermal expansion is neglected here), the amount of water does not change. AH remains ∼7.5 g/m^3^, but as 20°C air could potentially contain up to 17.3 g/m^3^ of water vapor, RH is now 43% only.

From the previous example, it appears that outdoor AH not only is conserved inside, but also determines indoor RH for a given indoor temperature. Low RH has been reported to increase sars-cov-2 and other viruses stability [21], so this could be a first causal mechanism linking outdoor AH and indoor infection probability: the virus survives better in dry indoor air.

Moreover, if the indoor temperature is held constant by heating, AH also directly determines indoor WBT. Building on the same reasoning as for the previous mechanism (ii), low AH coming from outside would lower indoor RH, which despite regulated indoor T would lower indoor WBT, resulting in a cooler, more fragile respiratory mucosa as individuals breathe dry indoor air. This could be a second possible causal link between outdoor AH and indoor infections.

Focusing back on outdoor weather, it is noteworthy that, as hot air can physically contain much more water vapor than cold air, a primary determinant of outdoor AH in the first place is outdoor T, and this translates into an important covariation between both environmental variables. The question is to know whether an effect of outdoor T on Rt operates through human behaviour (mechanism i) or through AH and its effects on indoor RH and WBT (mechanism iii/ii). In the vast majority of investigated states, AH and T correlations with Rt were of very similar strength. AH appeared as a better Rt predictor than T in a few states only (e.g. NM, CA, FL, WY), whereas T was a better predictor than AH in TX. Overall, it seems the present results are not sufficient to conclude on which of the proposed mechanisms is more likely, and to understand if T or AH is a better candidate as primary driver of indoor transmission.

### Limitations

The present results are highly dependent on the quality of raw data. New covid-19 case counts are manually collected from states’ public data [3], which can contain variable delays between states, but also even within a state, if the reporting policy changed through months in the pandemic. Large, prolonged surges can amplify issues, with increasing testing delays [6] or reporting backlogs, that add up to variable ICR delays, between and within states. The method used here is quite sensible to “spikes” in the data, as outliers have a heavy contribution to product-moment sums as computed in Pearson’s r. Here I chose to remove only the most disrupting spikes in 3 states (NC, MA, TX), otherwise sticking to the raw covid-19 new cases data (+ 7-day averaging). Other approaches with systematic, stronger smoothing are possible [see e.g. 10] and might allow higher correlation coefficients.

In states with small populations and/or very low case counts, Rt becomes more dependent on individual, stochastic cluster emergences, which can hide any potential weather determinism of transmission. For this reason, I do not expect that the present approach translates as well to infra geographic levels, e.g. counties.

State meteorological records from the *COVID-19 Open-Data* [11] were used as-is, and would also benefit from a careful re-examination. Ideally, hourly T and RH should be used to compute hourly AH and WBT before averaging to daily values. Also, the way individual weather station data are aggregated to yield state summary variables should be carefully considered by specialists, and maybe weighted by local county population. Such weather data quality-control might also contribute to revealing stronger correlations than what I report here, or higher homogeneity across states.

As each state most probably has its own epidemiological parameters (population density, mobility, compliance to social distancing policies, behavioral reaction to local surge reports, behavioral reaction to weather change, etc.), it should not be expected that the observed weather/Rt correlations superpose or line-up well between states (but this would deserve further investigation). There are a lot of unaccounted-for local factors that can shift or distort any effect of weather on disease transmission. Here we only tried to mitigate one of the most powerful factors, i.e. social distancing policy changes, by choosing the mid-June to mid-November period as the scope of the present study. Investigating how an effect of weather on Rt partially persists or disappears altogether during periods of stricter restrictions (e.g. current winter) would need further investigation, ideally including detailed data on the time course of social restriction measures by state.

For the same reasons, the present approach can be tried on other countries, but is expected to work better (1) in countries with limited and/or relatively stable restriction measures and (2) at aggregation levels large enough to yield significant (ideally >500) daily case counts. These two factors guided the choice of U.S. as a model nation for the present study.

Last, an hitherto overlooked confounding factor could be transmission from state to state. If the population has frequent inter-state mobility, a state with high infection levels could cause rise of infections in neighbor states. This phenomenon would presumably lower the level of local correlation between weather and Rt, hence contribute to deteriorate the present results. However, this dependency between neighbor states could also cause some level of autocorrelation in inter-state correlations. In other words, observing similar weather and similar Rt in 2 neighbor states should probably not be considered as 2 fully independent observations of a “weather effect”, if there was significant inter-state mobility.

### Applications

If a weather-Rt causal relationship gets confirmed by further investigations, there is a potential for predicting case count evolution about 10 days in advance, from the observed current daily weather data. In fact this would not be “forecasting”, only a way to catch up with the virus and “nowcasting” current transmission level (rather than retro-calculating Rt from case counts with a ∼2-week delay as is currently done). However, looking at the present results, and the confounding factors at play, such current transmission estimation from weather data might only be in terms of increased/decreased risk of infection, at best. This still could prove quite useful to try to outrun the continuing unpredictability of local pandemic surges.

Another prospective question is whether specific actions could be proposed to mitigate a possible increased infection risk during cold/dry weather episodes. Generic actions such as stricter social restriction measures during these weather episodes could be a first, debatable avenue. Concentrating strict measures (e.g. stay-at-home orders, business closure) on short high-risk periods could allow for a more normal social and economic life the rest of the time. More specific countermeasures (e.g. supplemental indoor water vapor sources, indoor RH monitoring) and adapted behaviors (e.g. avoidance of shared indoor spaces specifically during cold/dry weather episodes) can be imagined, but should be based on better understanding of the mechanism underlying the observed correlations.

## Data Availability

This study uses raw data from public projects. Processing code and processed data are available upon request.

## Acknowledgments

I thank the teams at the *Covid Tracking Project* and the *Covid-19 Open-Data* project for collecting, compiling and publicly providing data.

I also would like to thank the numerous online contributors that help make sense out of the pandemic: among others, biology and medicine scientists (A. Rasmussen, F. Kramer, B. Hanage, D. Costagliola, A. Kucharski, T. Bedford, A. Flahault, E. Topol, B. Watcher, R. Corsi and others), science journalists (E. Yong, K. Kupferschmidt, T. Louis and others), data visualization creators (J. Burn-Murdoch, G. Forestier, E. Orphelin, G. Saint-Quentin, G. Rozier, M. Pandaï), as well as participants to informal online discussions that stimulated the present work (J.-B. Mouret, M. Rahman, D. Marcombes, Fabien L., A. Modesto, R. Demol).

